# An agent-based model of COVID-19 in the food industry for assessing public health and economic impacts of infection control strategies

**DOI:** 10.1101/2024.06.18.24309041

**Authors:** Christopher Henry, Ece Bulut, Sarah I. Murphy, Claire Zoellner, Aaron Adalja, Diane Wetherington, Martin Wiedmann, Samuel Alcaine, Renata Ivanek

## Abstract

The COVID-19 pandemic exposed challenges of balancing public health and economic goals of infection control in essential industries like food production. To enhance decision-making during future outbreaks, we developed a customizable agent-based model (FInd CoV Control) that predicts and counterfactually compares COVID-19 transmission in a food production operation under various interventions. The model tracks the number of infections as well as economic outcomes (e.g., number of unavailable workers, direct expenses, production losses). The results revealed strong tradeoffs between public health and economic impacts of interventions. Temperature screening and virus testing protect public health but have substantial economic downsides. Vaccination, while inexpensive, is too slow as a reactive strategy. Intensive physical distancing and biosafety interventions prove cost-effective. The variability and bimodality in predicted impacts of counterfactual interventions, explained by the chance effects and early stochastic infection die-off, caution against relying on single-operation real-world data for decision-making. These findings underscore the need for a proactive infrastructure capable of rapidly developing integrated infection-economic mechanistic models for the essential industries to guide infection control, policy-making, and socially acceptable decisions.

## Introduction

The United States (US) food industry, known for its labor-intensive nature [1], was significantly affected by the Coronavirus disease 2019 (COVID-19) pandemic, alongside other essential industry sectors [2]. Early in the pandemic, labor shortages led to closures or reduced production in food facilities/operations [2, 3]. US livestock processing, including poultry, pig, and cattle slaughter, was reduced by up to 45%, resulting in job losses, financial impacts, retail shortages, and loss of animals [4, 5]. During 2020 alone, the combined value of production for beef, pork, broilers, turkeys, eggs, and milk was reduced by $12.8 billion, 9% below the pre-pandemic forecasts [6]. Dairy supply chain disruptions caused increased milk dumping, where 2.5% of all federally regulated milk was dumped compared to the usual 0.2-0.5% [7]. By September 2021, nearly 100,000 workers in US meatpacking facilities, food processing facilities, and farms were reported positive for COVID-19 [8]; importantly, these statistics are likely underestimated [9]. These outbreaks drove infection rates in rural communities [10], with the estimated mortality and morbidity costs of community infections exceeding US$11.2 billion by the end of 2020 [11]. These impacts affected the functioning of the national food supply chain.

Multiple strategies have been considered, encouraged, or enforced to control the spread of the severe acute respiratory syndrome coronavirus 2 (SARS-CoV-2) within the US food industry [12, 13]. These strategies include vaccination, practicing physical distancing, use of face coverings, screening for infection, practicing personal hygiene (e.g., hand washing), cleaning and disinfection of the working environments, ventilation improvements, and minimizing community spread. We use food ‘operation’ for brevity when referring to any individual fruit or vegetable (produce) farm operation or food processing facility. While having certain features in common, food operations vary greatly in size, physical characteristics, and organizational structure [14]. Their varying locations, policies, and workforce demographics have resulted in significant differences in worker histories with respect to vaccination, boosting, and past infection [15, 16]. The diversity of food operations and mitigations have led to strong interest from industry stakeholders in modeling tools tailored to the particular characteristics of their individual operations [17]. These tools would aid industry stakeholders in making predictions, such as regarding the expected outbreak dynamics and impacts (public health and economic) of possible interventions, and decisions, such as what level of investments to make in biosafety measures or when to start or stop an intervention (personal communication with the Industry Advisory Council for the study).

Mathematical models have been developed to evaluate and compare COVID-19 mitigation strategies and assess their effectiveness [18–30]. These models were primarily designed for national scale assessments [21, 25–27] but also include more localized communities, encompassing cities [24, 28, 30], closed societies with shared environments [19, 29], and even smaller communities in universities [22], companies [20, 23], and office spaces [18]. Evaluated interventions include physical distancing, mask use, vaccination, testing, contact tracing, quarantine, restrictions on travel, isolation, and school closures [18, 19, 26–28]. While most modeling studies concentrate on health outcomes in the general population, a handful have considered the health of individuals within workplace settings. These settings include a generalized company building [20], an oil and gas facility [23], a meatpacking plant [29], and a university building [22]. These studies have accounted for the complex disease transmission process between individuals using agent-based models (ABMs), which can simulate employees’ decisions based on their social and physical profiles. A few studies utilized ABMs to simulate the economic impacts of COVID-19 [24, 25, 30, 31]. Nevertheless, there remains a need for models that assess COVID-19 spread and the health and economic impacts of mitigation strategies in individual food operations to improve their resilience in future infection outbreaks.

Here, we provide **F**ood **Ind**ustry **CoV**id-19 **Contr**ol To**ol** (FInd CoV Control), a customizable tool based on an ABM developed to simulate COVID-19 transmission in a food operation and assess public health and economic impacts of interventions. Our objective was to develop a tool that helps policymakers and individual food operations navigate infection control in the essential food industry sector.

## Results

### Model setup

FInd CoV Control consists of three modules: Employee population, Work environment, and Disease transmission (**Figure 1A**; definitions and further details in **Supplementary Texts S1-S4** and **Tables S1-S15**). The modules define a single modeled food operation, which is either a produce (i.e., fruit or vegetable) farm or food processing facility (i.e., a meat or produce packing plant or an operation processing foods, such as beef, pork, poultry, milk, or fresh produce). The Employee population includes all employees (agents) in the modeled food operation, each of which is characterized by a set of attributes (**Table 1**). Attributes that represent past events and current state include age, (directly) immunity-related attributes, vaccination history, and the current state of infection, if any. The Work environment module defines the characteristics of the work environment in terms of a produce farm or processing facility setting (hereafter referred to as ‘farm’ and ‘facility’ for brevity), shift schedule (**Figure 1B.i**), and agent hierarchy and contact network (**Figures 1B.ii, 1B.iii, and Fig. S1**). The Disease transmission module tracks COVID-19 infection spread in the employee population, using an elaborated variant of a “Susceptible-Exposed-Infectious-Recovered-Susceptible” (SEIRS) model (**Figure 1B.iv**). FInd CoV Control is customized to the population and work environment of a particular food operation based on the user-set parameters (**Table 2**), which are also used to calculate the number of agents with various immunity trajectories and histories (**Table S1**). For simplicity, we assumed that all workers live in the same type of housing (i.e., either individual or employer-provided shared housing). To emphasize typical behaviors in these two housing settings, we also assumed that workers in individual housing do not socialize with each other outside of work but they are exposed to the community transmission of COVID-19 when not working, and that workers in shared housing socialize with each other outside of work but are not exposed to the community transmission. All contact between workers was assumed to occur either (a) while traveling to, at, or traveling home from work, or (b) in shared, employer-provided housing.

**Figure 1.**
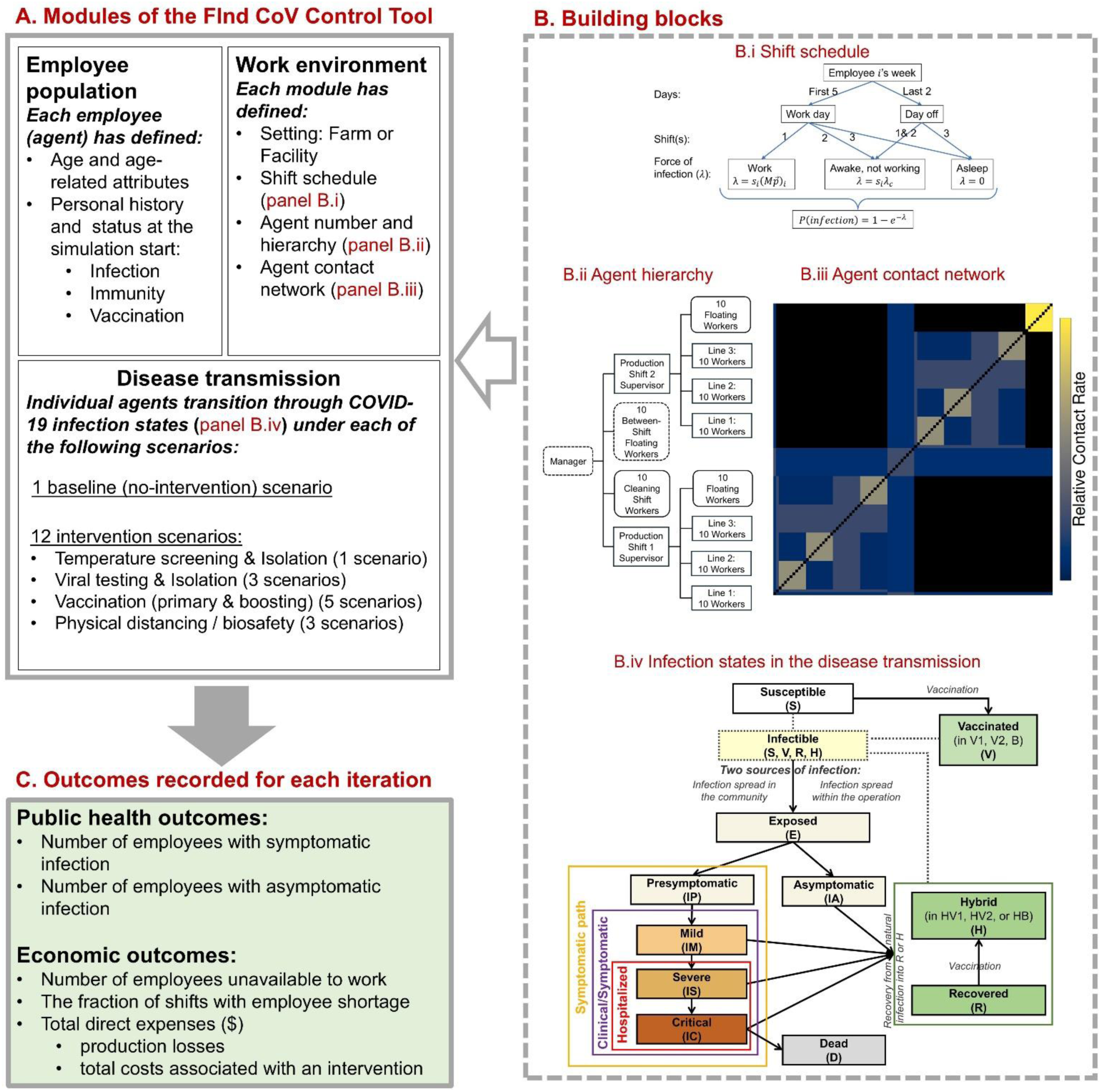
Overview of the FInd CoV Control tool. (A) Three modules of the tool: Employee population, Work environment and Disease transmission. (B) Module building blocks: B.i Example of a shift schedule (Note: shift (i.e., 8-hour time step) is the time unit of the model; in a week, there are 5 work days and 2 days off (weekend). On a work day, the example agent shown spends shift 1 awake at work, shift 2 awake and not working, and shift 3 asleep, while on a day off, the agent spends shifts 1 and 2 awake not working and shift 3 asleep), B.ii Agent hierarchy in a processing facility, B.iii Heatmap showing a contact network among agents in a processing facility and relative rates of contacts, and B.iv Infection states in the COVID-19 disease transmission module. (C) List of outcomes recorded for each iteration of a simulation with the FInd CoV Control Tool. In B.iii, the relative contact rates are calculated from user set parameters in **Tables 2** and **S11**. These relative contact rates are multiplied by a constant factor (i.e., scaled) to obtain the user set *R_0_*(s) when calculating the effective contact rates. Because of scaling, the values of relative contact rates for individual employee pairs in the contact matrix should only be interpreted relative to each other. For illustration, the contact matrix shown here is for a default facility model with individual housing; the relative contact rates range from 0 to 1.685824, where 0 is for a pair of employees who are not expected to make any contacts at work (if permanently assigned to different shifts), 1.685824 is for a pair of employees with the most frequent contacts for the given parameter set (which in the default facility model are cleaning shift employees), and 1 is the value for two workers on the same production line.

**Table 1.**
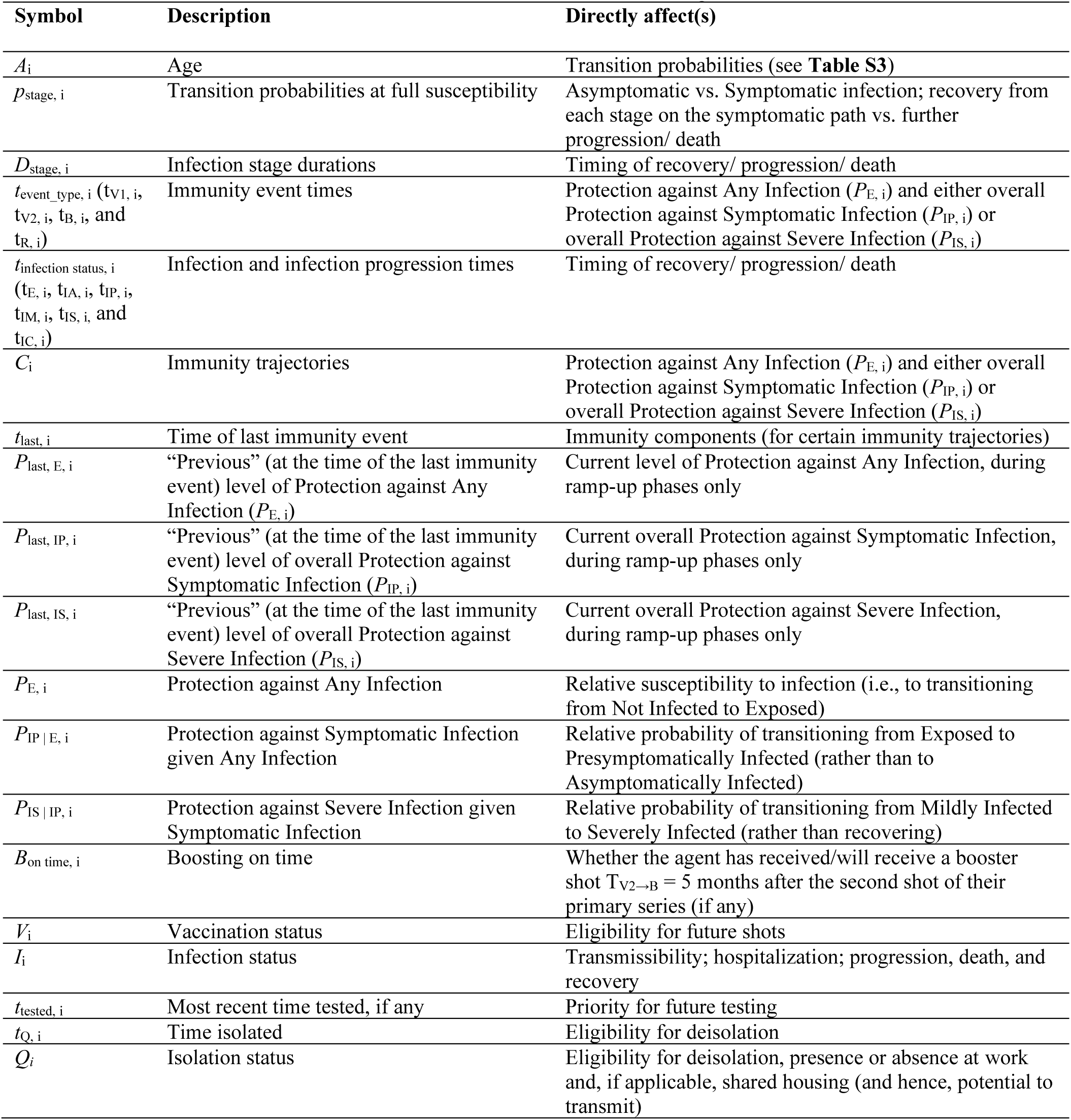
Summary of agent attributes. *‘Ramp-up’ refers to an initial increase in immunity following a vaccination event. Additional information for *P*_last, IP, i_ and *P*_last, IS, i_ is in **Table S4**. Distributions for infection stage durations are in **Table S2. Table S14** is a version of this table with additional notes and information on how attributes are set at simulation start and/or during the simulation.

**Table 2.** User-settable model parameters.

| Symbol | Definition (unit) | Possible values | Default value | References and Notes |
| --- | --- | --- | --- | --- |
| $N$ | Total number of agents (employee) | Farm: $\geq 4$<br>Facility: $\geq 7$ | 103 | From user input below using the approach in **Table S13** |
| $N_E$ | Initial number of agents who are in the Exposed state (E) (employee) | $1-N$ | 1 | Assumed |
| $N_{IM}$ | Initial number of agents with Mild COVID-19 symptoms (employee) | $0-(N - N_E(0))$ | 0 | Assumed |
| $f_R$ | Initial proportion of agents who have recovered from COVID-19 infection within the past $T_{RQ} = 1$ year (including those who were not symptomatic) (unitless) | 0%–100% | 69% | [32, 34, 35] (Derivation in **Text S11**) |
| $f_{V2}$ | Initial proportion of agents who are fully vaccinated as part of the primary series of a COVID-19 vaccine (e.g., 2 shots of Pfizer) (unitless) | 0%–100% | 71% | [32, 33] (Derivation in **Text S11**) |
| $f_{V2, \text{recent}}$ | Initial proportion of agents who have become fully vaccinated within the past $T_{V2 \rightarrow B} = 5$ months (unitless) | $0\% - f_{V2}\%$ | 9% | [32, 33] (Derivation in **Text S11**) |
| $f_B$ | Of agents who are eligible to receive a booster shot (i.e., have completed their primary series of COVID-19 vaccination at least $T_{V2 \rightarrow B}$ ago), initial proportion who have received a booster shot (unitless) | 0%–100% | 45% | [32, 33] (Derivation in **Text S11**) |
| $f_{B, \text{recent}}$ | Of agents who are eligible to receive a booster shot, fraction who have received a booster shot within the past $T_{V2 \rightarrow B}$ (unitless) | $0\% - f_B\%$ | 45% | [32, 33] (Derivation in **Text S11**) |
| $T$ | Number of days to simulate (day) | 30–150 | 90 | |
| $H$ | Employee housing type (categorical) | “Shared” or “Individual” | Farm: “Shared”<br>Facility: “Individual” | Farm or facility specific |
| $R_{0, \text{housing}}$ | Contribution of (expected) transmissions in shared housing to $R_0$ for a given level of physical distancing (High”, “Intermediate”, or “Low”) (unitless) | “High”: 1,<br>“Intermediate”: 2, or<br>“Low”: 4 | 2 | Assumed<br>When housing is “Shared” |
| $\lambda$ | Daily force of infection from sources outside of the workforce of the operation modeled for a given level of physical distancing (“High”, “Intermediate”, or “Low”) (employee<sup>-1</sup> day<sup>-1</sup>) | “High”: 0.02,<br>“Intermediate”: 0.002 or<br>“Low”: 0.00002 | 0.002 | Assumed<br>When housing is “Individual” |
| $R_{0, \text{work}}$ | Contribution of work transmissions to (homogeneous) $R_0$ for a given level of physical distancing (High”, “Intermediate”, or “Low”) (unitless) | “High”: 4,<br>“Intermediate”: 6, or<br>“Low”: 8 | 6 | Assumed based on physical distancing to/at work |
| $n_{w, c}$ | Number of field workers per crew (excluding foreman) (employee) | 1–100 | 10 | Farm specific |
| $n_{c, s}$ | Number of crews per supervisor (employee) | 1–100 | 3 | Farm specific |
| $n_s$ | Number of supervisors (employee) | 1–100 | 3 | Farm specific |
| $D_{\text{weekly}}$ | Total production value per week (default defined by employee number) (US \$) | 0–1,000,000 | 247,612 | Farm specific |
| $W$ | Average hourly wage of a worker (US \$) | 1–100 | 13.89 | Farm specific |
| $n_{w, l}$ | Number of workers per production line (employee) | 2–100 | 10 | Facility specific |
| $n_l$ | Number of production lines (lines) | 1–100 | 3 | Facility specific |
| $n_{sh}$ | Number of production shifts (employee) | 1 or 2 | 2 | Facility specific |
| $n_{cs}$ | Number of cleaning shift workers (employee) | 2–100 | 10 | Facility specific |
| $n_{f,sh}$ | Number of floating workers in a production shift (e.g., quality assurance technician, mechanic) (employee) | 1–100 | 10 | Facility specific |
| $n_{f,all}$ | Number of workers that may be present across shifts (including manager) (employee) | 1–100 | 11 | Facility specific |
| $D_{week}$ | Total production value per week (default defined by employee number) (US \$) | 1–10,000,000 | 784,346.67 | Facility specific |
| $H_w$ | Average hourly wage of a production line worker (US \$) | 1–100 | 16.57 | Facility specific |
| $F$ | Indoor facility size (for estimation of the cost of a HEPA air cleaner) (sq ft) | 1–100,000 | 1,000 | Facility specific |

We used FInd CoV Control to predict COVID-19 transmission dynamics following the arrival at work of an index case infected outside the workplace under a no-intervention “baseline” and various interventions. The evaluated interventions (**Figure 1A; Text S5**) included:

- “Temperature screening” (1 scenario) denotes temperature screening and isolation where all employees arriving for their shift are tested prior to admitting employees to work for that shift. The temperature threshold is set at 38°C.
- “Virus test” (3 scenarios) denotes low-, moderate-, and high-intensity of viral testing and isolation, respectively modeled with the probability *p* = 5, 30, or 100% of each employee being tested each shift upon arrival at the workplace. The isolated employee is unable to spread the infection to co-workers either at work or in shared housing (if applicable).
- “Vaccine” (5 scenarios) denotes primary “vaccination” of unvaccinated workers with 2 doses at a daily probability of *p* = 2 or 4%; “boosting” of boosting-eligible but unboosted workers at *p* =2 or 4% per day; or a combination of both primary and boosting vaccination interventions (“vax+boosting”) at *p* = 2% per day.
- “Physical distancing/Biosafety” (3 scenarios) where physical distancing and/or biosafety interventions are modeled as generating a 20, 40, or 80% reduction in the basic reproduction number (*R_0_*) at work (i.e., −20, −40 or −80% *R_0_*), such as through the application of masks, face shields, physical barriers, and/or ventilation. The exact approach necessary to achieve these desired effects on *R_0_* will vary across real-world work environments. However, to illustrate how higher-effectiveness interventions may be built by “stacking” multiple lower-effectiveness strategies and to be able to quantify their net cost in the subsequent economic analysis, in absence of required data we assumed: (i) For a low-effectiveness (−20% *R_0_*) intervention, the use of KN95 masks, one per employee per shift; (ii) For a moderate-effectiveness (−40% *R_0_*) intervention, a low-effectiveness intervention combined with the use of face shields, one per employee per 30 days; (iii) For a high-effectiveness (−80% *R_0_*) intervention, a moderate-effectiveness intervention combined with ventilation improvement using (a) portable high-efficiency particulate air (HEPA) fan/filtration system(s) with an average cost of $1,000 per 1,000 sq. ft. (where the sq. ft. of the operation area is set by the user, **Table 2**).

The baseline and interventions were simulated in a way that allows counterfactual comparisons. Interventions were evaluated using two groups of metrics: (i) Public health: the number of employees with symptomatic and asymptomatic infection (and total infected), and the initial effective reproductive number (*R_eff._*); and (ii) Economic: the number of employees unavailable to work, the fraction of shifts with employee shortage, and total direct expenses, production losses, and total costs associated with an intervention (expressed in US$) (**Figure 1C**). While economic effects are often interrelated and ripple over multiple dimensions, in FInd Cov Control, the economic analysis is limited to the costs directly borne by food operations (**Text S5**) and is meant to serve as a reference, together with the infection model, for employers’ decision-making.

### Model validation

FInd CoV Control was validated with publicly available data on outbreaks from early in the pandemic when few, if any, interventions would have been implemented. For produce farms, FInd CoV Control was validated using two outbreaks, one on a fruit orchard farm with shared (i.e., employer-provided dormitory style) housing and the other on a strawberry greenhouse farm with a mix of shared and individual housing. For processing facilities, FInd CoV Control was validated using outbreaks in three facilities with individual housing, one in each of the dairy, pork, and produce processing facilities. The results of the validation analysis indicated a reasonable fit between the reported data and model predictions (**Text S6**).

## Main results

We present a representative set of results over a 90-day simulation (Figures 2-4) for a processing facility with 103 employees, shared housing, and otherwise default parameters (**Table 2** and **Table S2**). Results pertaining to the number of symptomatic infections over time show little qualitative difference between the curves depicting the mean incidence (Figure 2A) and prevalence (Figure 2B), although there is a difference in scale and a slight difference in location (with incidence peaking slightly before prevalence) and noisiness (with more visual noise in the incidence vs prevalence curves). This result is expected, so we focus on (cumulative) incidence in the remaining panels to avoid redundancy. Insights from analyzing a large facility (**Figs. S2, S3** and **S4**) and the produce farm are similar (**Text S7**). Subject to model limitations, Figures 2-5 show that intensive physical distancing and biosafety interventions have the best combination of health and economic effects. Temperature screening and virus testing are optimal from the public health point of view but are costly, while vaccination is too slow as a reactive strategy though relatively cheap. These results are explained in the following paragraphs.

**Figure 2.**
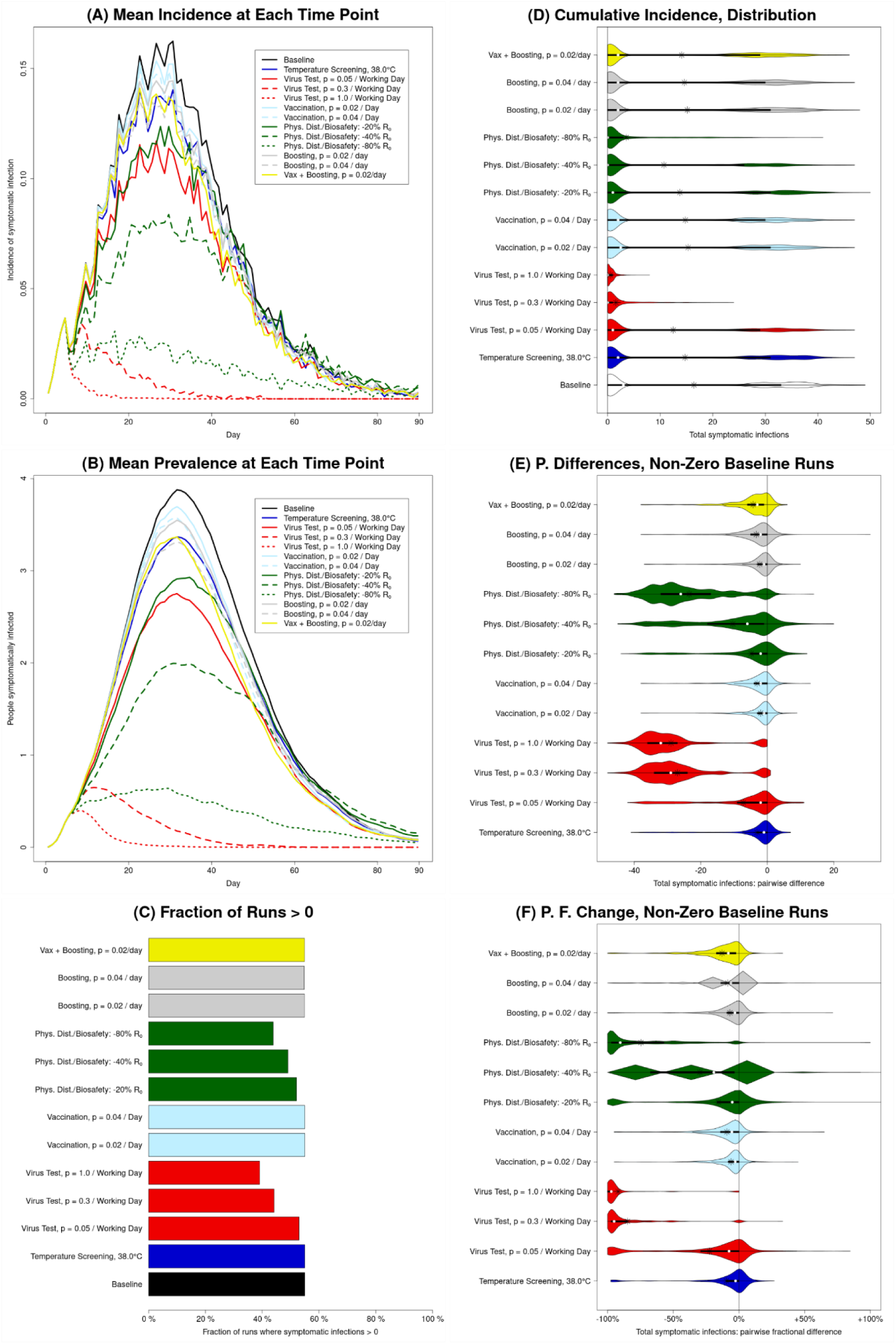
Illustration of public health outcomes for baseline (no intervention) and each of the interventions in absolute terms as well as relative to the baseline. Results for number of symptomatic infections in a processing facility with 103 employees over the 90 days of the simulation run are shown (results for total infections are similar, apart from scale). (A and B) The mean across all runs of the incidence (A) and prevalence (B) of symptomatic infection, at each time point; these illustrate the dynamics over time, but also conceal the high level of variation between runs. (C) The fraction of runs for which the total number of symptomatic infections is greater than zero. (D) Violin plots representing the distribution, between runs, of the *total* number of symptomatic infections; these violin plots illustrate the bimodal nature of most distributions. (E) Violin plots representing the distribution of *counterfactual effects* of the various interventions, i.e., the distribution of *pairwise differences* between *corresponding* runs with and without that intervention (the number at that intervention, *N*_*I*_, minus the number at baseline, *N*_*B*_; *N*_*I*_ − *N*_*B*_), for runs that *do* have one or more symptomatic infections at baseline. (F) Violin plots representing the distribution of pairwise *fractional* differences (i.e., (*N*_*I*_ − *N*_*B*_)/*N*_*B*_), for runs with a non-zero number of symptomatic infections at baseline. For three of the interventions, there is a single positive outlier (i.e., 1 iteration out of 1,000 runs per intervention) that is cut off by the axis limits to avoid excessively compressing the depiction of the other 11,997 points.

**Figure 3.**
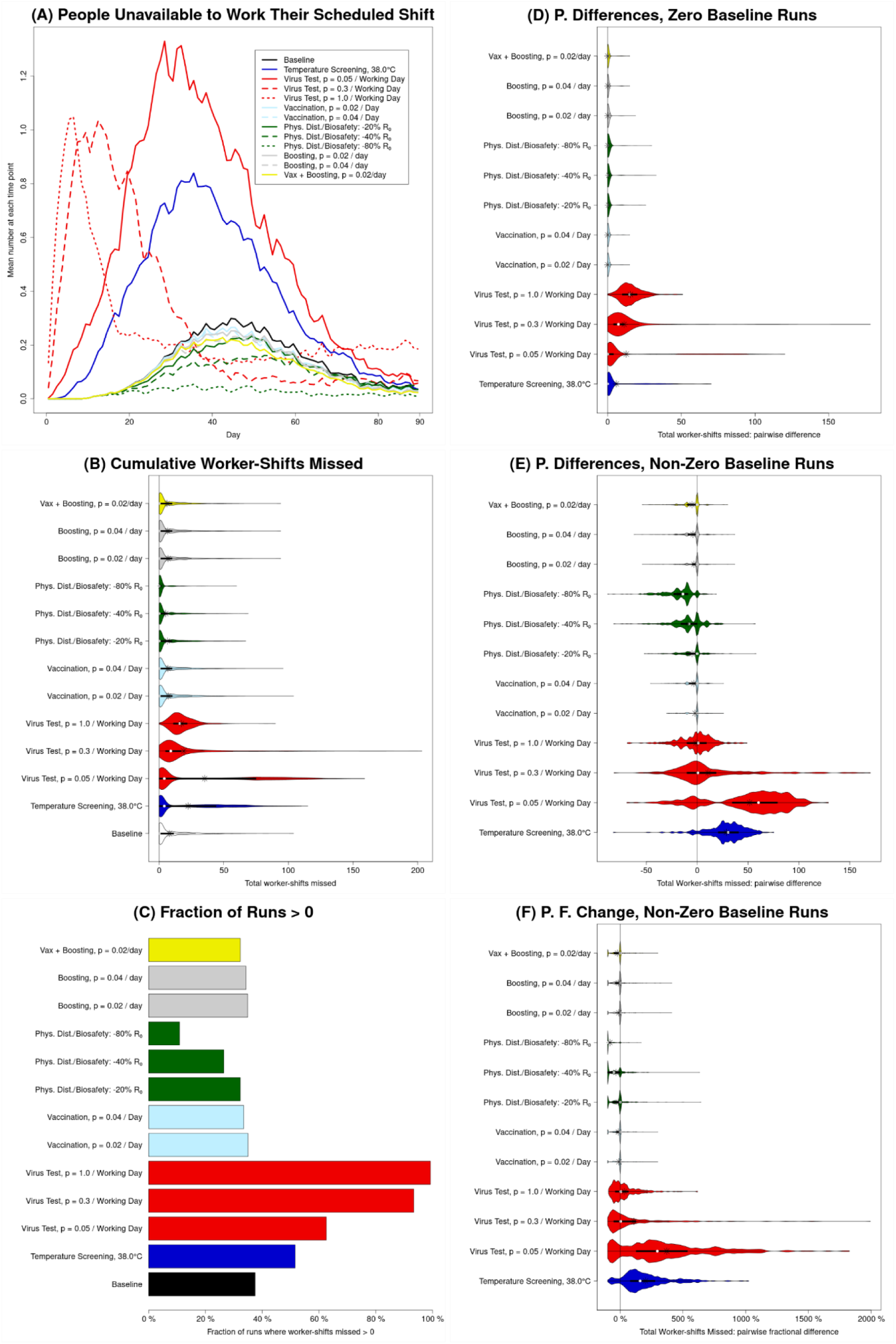
Illustration of unavailability for baseline (no intervention) and each of the interventions in absolute terms as well as relative to the baseline. All results are for a processing facility with 103 employees over 90-day long simulation runs. Unavailability (i.e., worker-shifts missed) depends not only on how many employees are infected, and how many of those are symptomatic, but also on how likely an infected employee (whether symptomatic or asymptomatic) is to be removed from the workforce (due to hospitalization, or detection and isolation). (A) The mean across all runs of the number of employees unavailable to work their scheduled production shift, for each day of the simulation; this illustrates the dynamics over time, but also conceals the substantial level of variation between runs. (B) Violin plots representing the distribution, between runs, of the sum of the number of workers unavailable to work their scheduled production shift, over all such shifts; this violin plot illustrates the varying shapes of these distributions. (C) The fraction of runs for which the total number of worker-shifts missed is greater than zero. (D and E) Violin plots representing the distribution of *counterfactual effects* of the various interventions, i.e., the distribution of *pairwise differences* between *corresponding* runs with and without that intervention (the number at that intervention, *N*_*I*_, minus the number at baseline, *N*_*B*_; *N*_*I*_ − *N*_*B*_), for runs with zero (panel D) and non-zero (panel E) worker-shifts missed at baseline. (F) Violin plots representing the distribution of pairwise *fractional* differences (i.e., (*N*_*I*_ − *N*_*B*_)/*N*_*B*_), for runs with non-zero worker-shifts missed at baseline.

**Figure 4.**
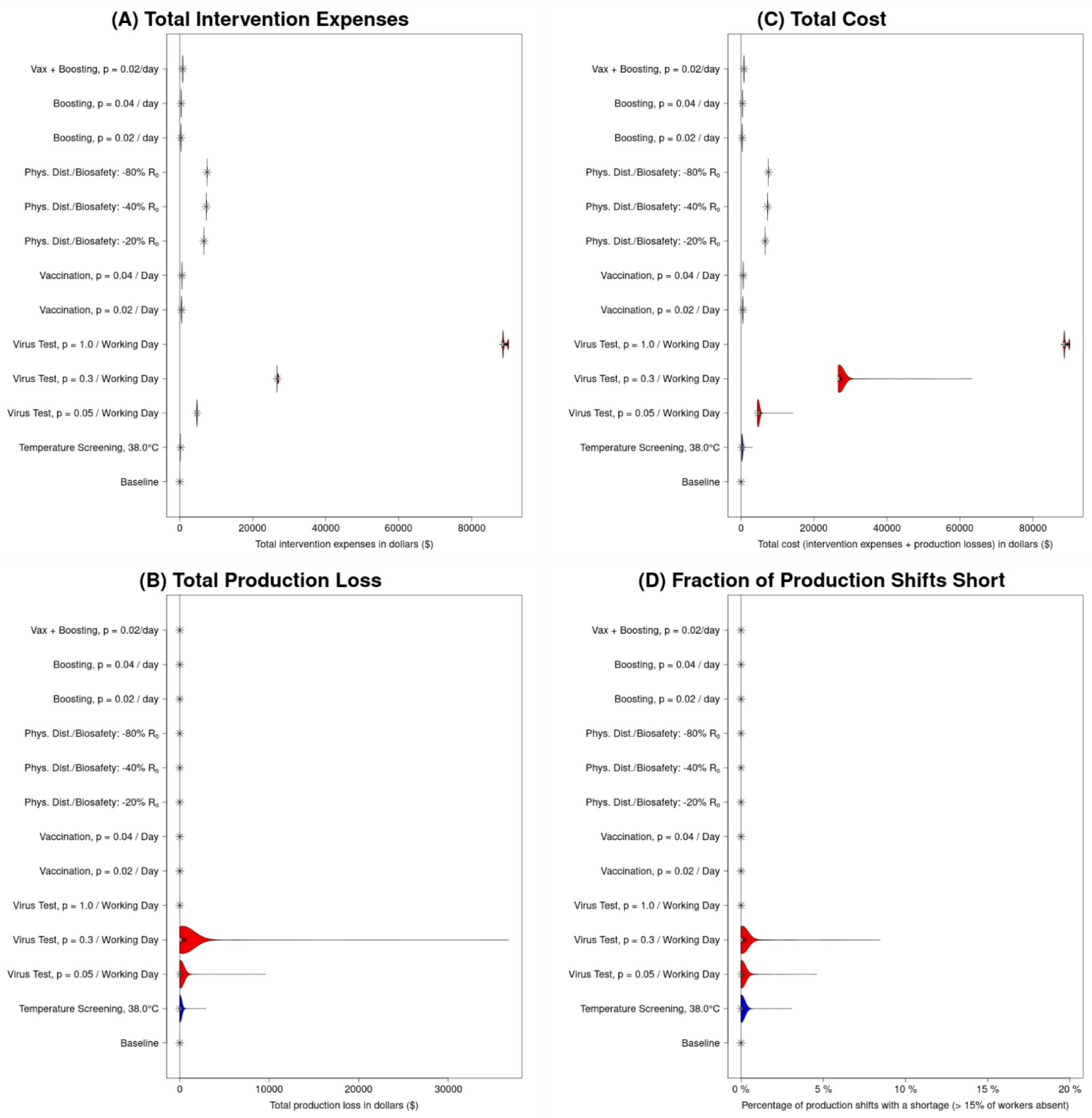
Illustration of costs for baseline (no intervention) and each of the interventions. All results are for a processing facility with 103 employees over 90-day long simulation runs. All panels consist of violin plots (although in some cases, these may be sufficiently horizontally compressed that this is not obvious) representing the distribution (across runs within an intervention) of an outcome. (A) Distribution of direct intervention expenses (supplies purchased and/or additional wages paid for tasks performed outside of an individual’s normal scheduled working hours); these are generally relatively constant for an intervention, and are always US$0 by definition for the baseline. (B) Distribution of production losses due to worker absences; as a result of unavailability (occurring only on days when >15% of workers miss their shift) this is almost always US$0 in the absence of a testing intervention. Here, we can see that low to moderate levels of routine viral testing may be insufficient to interrupt transmission, but sufficient to remove significant numbers of employees through detection and isolation, and thus causing significant production losses. (C) Distribution of total costs (in US$), which we define to be the sum of intervention expenses and production losses. (D) Fraction of production shifts (within a single run) that are “short”, i.e., more than 15% of workers absent (“0%” means that in a particular run none of the shifts were “short”).

**Figure 5.**
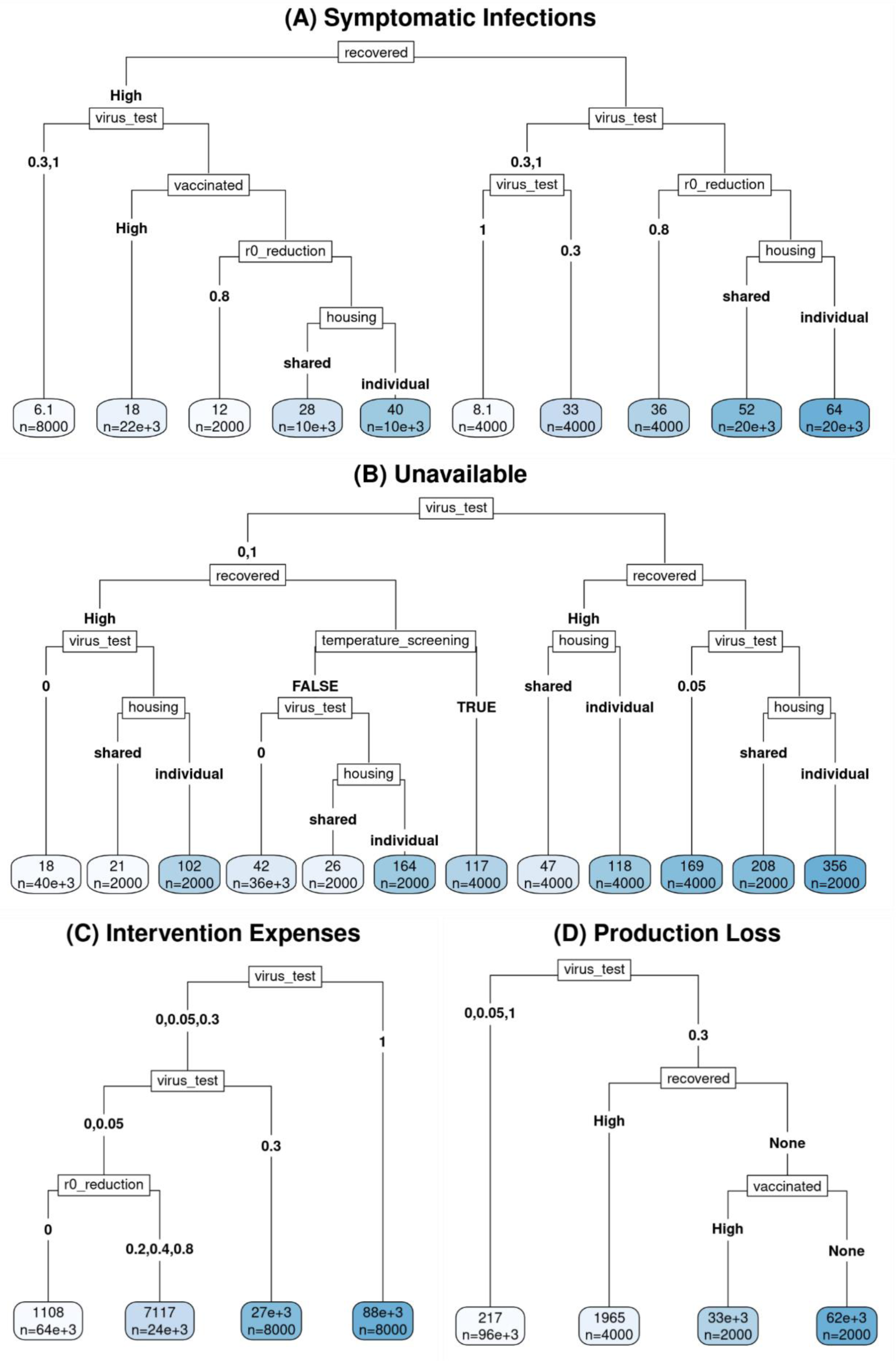
Regression Trees. For all panels, the labels on the branches descending from a node represent values of the parameter listed in the node itself. Where there are only two values for a parameter, the value is sometimes only listed explicitly on the left branch, to save space; the right branch simply has the value of that parameter that the left branch does not. In all cases,branches are ordered by making the left branch the one with the *lower* average value for the outcome represented in that panel. (This does not, however, result in *all* leaves being ordered from lowest to highest, because the branches are not allowed to cross.) The value at each leaf indicates the mean value of the outcome across relevant scenarios x interventions x runs over the 90 days of the simulation run, and *n* indicates the number of runs represented by the leaf (out of 104,000 runs). (A) Total number (Cumulative Incidence) of Symptomatic Infections, (B) Total number of Worker-Shifts Unavailable, (C) Cumulative intervention expenses (in US$), (D) Cumulative production losses (in US$).

### Bimodality and variability in symptomatic infections

For all interventions (including the no-intervention baseline), over 40% of all runs result in no symptomatic infections at all (Figure 2C), and as a result, the number of symptomatic infections for a given intervention is strongly bimodal at baseline and for all interventions except for moderate viral testing (*p* = 0.3, i.e., 30% of scheduled workers tested each shift, amounting to testing every worker 1.5 times per week), high-intensity viral testing (*p* = 1, i.e., every worker scheduled for testing each shift), and “high-effectiveness (−80% *R_0_*) Physical distancing/Biosafety” (Figure 2D). This reflects that, in the absence of repeated reintroduction from the broader community, a major source of variance is the possibility of early stochastic die-off, even at an initial *R_eff._* well above 1 (e.g., *R_eff._* = 2.52 at baseline for this scenario). The scholastic die-off effectively partitioned outcomes into two modal regions (groups) with respect to the predicted outbreak size: (i) small or non-existent outbreaks and (ii) large outbreaks (**Text S7**).

### Bimodality and variability in counterfactual effects of interventions

We can refine and expand observations in the previous section by counterfactual comparisons (see “Interventions” section); specifically, the *i*-th run of the model with any intervention corresponds in a meaningful way to the *i*-th run of the model at baseline, with the difference between the two being attributable solely the intervention. Consequently, it is meaningful to examine the pairwise differences, with respect to a particular outcome, between corresponding runs with and without a given intervention. The distributions of these pairwise differences for runs with at least one symptomatic infection at baseline are presented in Figure 2E. There, we can see that there is not only a great deal of variance in outcomes within an intervention (or within the baseline), but also a substantial variance in the counterfactual effects of an intervention. This is meaningful, considering that the individual ABM runs reflect the real-world variation in epidemiologic outcomes, and the model provides a view into the counterfactual impacts of interventions that cannot be observed in the real world. As an example, the “moderate-effectiveness (−40% *R_0_*) Physical distancing/Biosafety” intervention is modestly beneficial on average (mean reduction in number of symptomatic infections = 10.4), but can be extremely effective (maximum reduction of 45, close to the maximum across all interventions of 48), and entirely ineffective or even counterfactually *counterproductive* (68 runs with an *increase* of 1-20) (Figure 2E). This is due to the timing and chance effects (**Text S8**). We can also note that most of these distributions of pairwise differences are themselves bimodal, reflecting two ways that an intervention can counterfactually affect a run that produces a large outbreak in the no-intervention scenario. In these counterfactual comparisons, on the one hand, an intervention may prevent a large outbreak altogether, producing a data point in the high-effectiveness modal region in Figure 2E (and contributing to the difference in the number of large outbreaks between the intervention and no-intervention scenarios in Figure 2D). Alternatively, it may produce a smaller difference in the outbreak size (or none at all), producing a data point in the low-effectiveness modal region (and still contributing a large outbreak to both the intervention and no-intervention scenarios in Figure 2D).

We can further refine these observations by considering the change in number of symptomatic infections as a fraction of the baseline number of symptomatic infections (Figure 2F; **Fig. S2F** for a large facility). For those interventions with a significant fraction of runs in the high-effectiveness modal region, the primary way that they shift runs from having large outbreaks in the no-intervention scenario to not having large outbreaks in the presence of the intervention is by causing them to have no symptomatic infections at all; the apparently symmetrical lower modes seen in Figure 2E are primarily produced by variation in the number of infections (in a *large* outbreak) at baseline, not by variation in the number of infections (in a small or (effectively) non-existent outbreak) under the intervention.

### “All good things in moderation” may backfire in viral testing

The most effective reductions in symptomatic infections are seen for moderate- and high-intensity viral testing (Figure 2F). Figure 4 (**Fig. S4** for a large facility) shows that viral testing at a fairly high rate is also generally more costly. In the case of high-intensity viral testing, this cost is overwhelmingly due to direct intervention expenses (primarily the cost of test kits), which is unsurprising considering this strategy results in the lowest incidence and thus prevents production losses caused by employee unavailability (Figure 2A). This is also true in most runs for moderate-intensity viral testing, but in some runs, moderate-intensity viral testing can result in significant costs due to both direct intervention expenses and production losses. The latter reflects the ability of testing at a “moderate” rate (*p*=0.3/working day) to generate a “worst of both worlds” scenario. This scenario generates large numbers of employees who are isolated at the same time, resulting in large numbers of worker-shifts missed due to isolation, yet, infected employees are not identified and isolated fast enough to prevent a large outbreak from occurring (and consequent unavailability). On the other hand, testing frequently (*p* = 1/working day) quickly identifies and isolates cases, and while the isolation of identified cases contributes to unavailability, it also prevents the infection from spreading to other employees, thus preventing the unavailability of those prevented cases. A more frequent version of this can be seen for low-intensity viral testing (*p* = 0.05/work day), where mean and median increases in unavailability are both greatest (Figures 3E and **3F**; **Figs. S3E** and **S3F** for a large facility). This observed pattern reinforces and extends the result from prior research that existent but inadequate larger-scale (city-level) non-pharmaceutical interventions can result in what the authors describe as a “dual blow of increased deaths and unemployment,” which fall disproportionately on low-income workers [30].

### Health benefits of physical distancing/biosafety interventions at low cost

After the moderate- and high-intensity viral testing, the next-most effective intervention in preventing symptomatic infections is the “high-effectiveness (−80% *R_0_*) Physical distancing/Biosafety” intervention (Figure 2F), for illustration purposes represented by a combination of masking, face shield use, and ventilation improvements. We found this to be much more effective than the “moderate-effectiveness (−40% *R_0_*) Physical distancing/Biosafety” intervention (represented by masking and face shield use, without ventilation improvements), but only modestly more costly (and substantially less costly than the more effective viral testing interventions) (Figure 4).

### Scenario analysis

For our scenario analysis, we first defined several elements (factors) whose effects and interactions with intervention effects we wished to examine (Figure 5). These scenario elements were “setting” (“farm” vs. “facility”), “housing” (“individual” vs. “shared”), “vaccinated” (“high” (based on US national levels in early 2022 [32,33]) vs. “none”), and “recovered” (“high” US national levels [32, 34, 35] vs. “none”) (details in section **Text S9** and **Table S14**). We then conducted a full factorial analysis for all 16 combinations of these four factors (and for all 13 intervention scenarios) in the default facility size of 103 workers over 90-day-long simulation runs. Many combinations of these scenarios are intended to represent limiting cases, rather than realistic scenarios, e.g., a scenario with both “vaccinated” = “high” and “recovered” = “none” represents a limiting case of relatively high vaccination and no history of infection. Results of this analysis, evaluated using regression trees for each of the three primary outcomes (symptomatic infections, worker-shifts unavailable, and total cost) and the two separate contributors to total cost (production losses and intervention expenses), indicated that, for outcomes other than production losses, the effects of “setting” were relatively limited, and mostly pertained to which *other* effects were strong enough to be included in the pruned partition trees. Because transmission in the two settings is defined by the user-settable value of *R_0_* (**Table 2**), which was kept the same between the two settings, the observed differences between settings can be attributed to differences in the work environment (Figure 1B **and Fig. S1**). In general, the evaluated outcomes were slightly higher for the “facility” than for the “farm” setting. Because of this, and to omit explanations of the different equations used to set default production-per-week in the two different settings, we chose to describe results from the facility model here. Because total cost is simply a sum of intervention expenses and production losses, and because various factors affect each component differently, in the following paragraphs we will focus our discussion on each cost individually.

### Interactions between the worker infection history and intervention intensity drive outcomes

For almost all evaluated outcomes, the two biggest factors driving outcomes are the intensity of virus testing intervention and “recovered” (i.e., whether the employee population has a significant history of natural infection) (Figure 5); the only exception is intervention expenses (Figure 5C), for which intensity of virus testing was the strongest factor, but “recovered” did not produce a sufficient impact to appear in the regression tree. While “recovered” being “high” (rather than “none”) had a desirable impact (i.e., produced lower symptomatic infections, unavailability, and intervention expenses) on all outcomes for which it was relevant, the effects of virus testing were more variable. Symptomatic infections are minimized by viral testing at a rate high enough to reliably control an outbreak before it can get large (testing every worker every shift or roughly every 3 shifts (*p* = 1 or 0.3/ work day, respectively)) (Figure 5A). Unavailability (Figure 5B), on the other hand, is lowest when *either* testing is non-existent (and so no workers are isolated as a result of testing, but only as a result of hospitalization) *or* testing is extremely intensive (and so the outbreak(s) is/are rapidly contained; *p* = 1/work day); however, even such intense testing may be insufficient to achieve a reasonable level of control in the face of a population with insufficient natural and hybrid immunity (“recovered” = “none”) and constant reintroduction of infection via workers infected in the community (housing = “individual”). Conversely, unavailability is *highest* when the testing rate is intermediate (*p* = 0.05 or, even more so, *p* = 0.3/work day), resulting in enough asymptomatic and mildly symptomatic cases being detected to increase unavailability, but not enough to rapidly contain the outbreak(s). Similarly, production losses (which are driven by unavailability of ≥ 15% on a production shift) are highest when *p* = 0.3/work day, to the point that recursive partitioning with default parameters results in further division only of the node with *p* = 0.3; all scenario-intervention combinations with either a lower (*p* = 0, *p* = 0.05) or a higher (*p* = 1) rate of virus testing are combined in a single node (Figure 5D). Intervention expenses, move in an opposite pattern to symptomatic infections, and are highest when the rate of viral testing per work day is highest (*p* = 1), and lowest when it is low (*p* = 0.05) or non-existent (*p* = 0) (Figure 5C).

### Strongest outcome drivers are viral testing intensity and history of natural infection

While “recovered” is consistently more important than “vaccinated” (i.e., splits defined by it occur closer to the root of each tree, where either occurs at all), and both share a uniformly desirable effect (where they show any effect at all), the interaction of the two is more complex: For production losses, there is only a split defined by “vaccinated” in a branch in which “recovered” is set to “none,” but for symptomatic infections, the reverse is true – the only split defined by “vaccinated” occurs in a branch in which “recovered” is set to “high.” In more conceptual terms, this amounts to saying that, at the population level, immunity resulting from recovery from natural infection plays a stronger role in determining a wide range of simulation outcomes than immunity resulting from vaccination; whether the interaction of these two is sub-additive or super-additive depends on which outcome one is considering. In particular, with respect to symptomatic infections, there may be a synergistic effect of vaccination and recovery from natural infection, likely reflecting the strong protective effect of modeled hybrid immunity. For production losses, on the other hand, vaccination is less influential in the presence of moderate-to-high levels of natural recovery within the past year. This likely reflects the threshold effect in our model of production losses (occurring only if >15% of workers are unavailable) – in the presence of sufficient protection from natural recovery, the probability of suffering production losses at all may be low enough, even in the absence of vaccination, to reduce the importance of vaccination in predicting or determining that outcome.

The only panel from which the “recovered” factor is absent, or even not one of the two strongest factors, is Figure 5C Intervention Expenses; the “vaccinated” factor is absent from this panel as well (although the cost of giving workers time off for vaccination is accounted for). This is unsurprising, given that intervention expenses are driven far more by what interventions one decides to implement than by transmission dynamics; as a result, *no* scenario parameters appear in it. The only split that does, other than the virus testing splits, is a split by whether there is a Physical distancing/Biosafety (“*R_0_* reduction”) intervention, which raises costs (mean cost = US$7,171 vs. US$1,108; **Text S5**) over the alternative (a weighted average of no-intervention baseline, temperature screening, and vaccination and/or boosting interventions), in line with what we would expect. Temperature screening, being similar in certain respects to virus testing, but substantially cheaper and generally substantially less effective, appears only in the tree for unavailability (Figure 5B), where its use increases unavailability, more than a maximal rate (p = 1) of viral testing (117 vs. 85=(26+164)/2).

### Intervention effectiveness is highly sensitive to the degree of community transmission

Housing affects both unavailability (Figure 5B) and number of symptomatic infections (Figure 5A); both are higher when housing is “individual”. This is not to say that “shared” dormitory housing is a poorer environment for transmission than individual housing; rather, it reflects the role of community transmission in creating opportunities for reintroduction of infection from outside the employee population. This result is confirmed and elaborated by tests in **Text S9**, where we treat presence or absence of community transmission and presence or absence of dormitory transmission as separate factors. In line with this, additional analyses (**Text S9**) further indicate that our predictions about intervention effectiveness can be highly sensitive to the degree of community transmission.

### “R_0_ reduction” strategies are cost-effective

Physical distancing/Biosafety interventions (“*R_0_* reduction”) can reduce the number of symptomatic infections (when sufficiently effective) with minor increase in intervention expenses (Figures 5A and **5C**, respectively). This suggests that highly effective *R_0_* reduction strategies are cost-effective and, hence, should be prioritized for implementation.

### Sensitivity analysis

Figure 6 shows the sensitivity of 5 representative interventions to the 5 parameters with the greatest impact across all evaluated parameter-outcome combinations. Symptomatic infections were affected by the mean duration (*μ*_IM_) of mild symptomatic infection and relative per-contact probability of transmission (*β*_IM_) during this stage, followed by a parameter (*φ*) governing the protection from developing symptomatic disease provided by recovered and hybrid immunity (**Text S10B**). Worker-shifts missed were additionally affected by the relative frequency of severe infection given any symptomatic infection (*ψ*) and the mean duration of severe infection (*μ*_IS_), while the effects of these 5 parameters on *R_eff._* and total cost were generally smaller (**Text S10C**).

**Figure 6.**
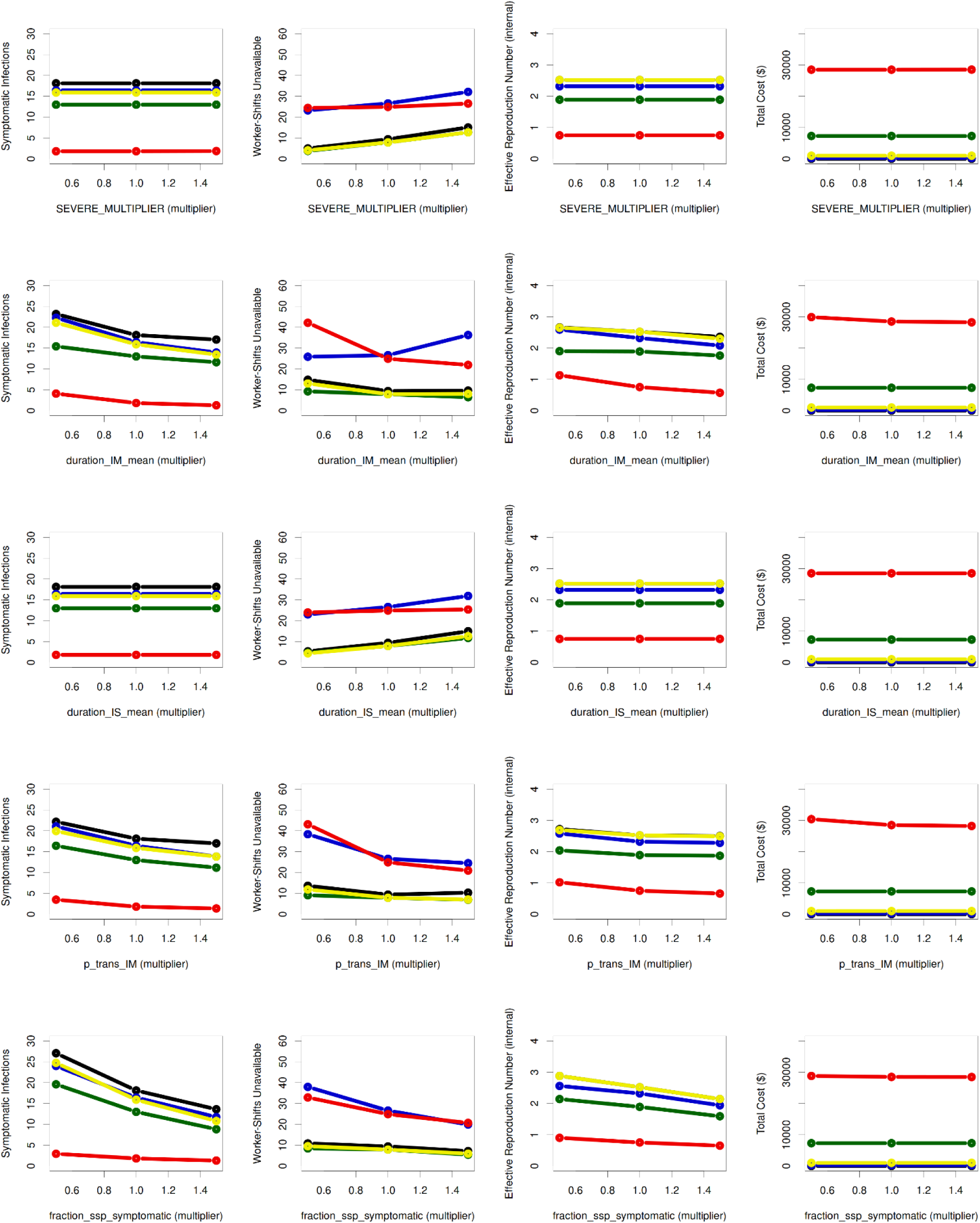
Sensitivity analysis. The plots show the dependence of the mean value of four major outcomes (columns), summed over the course of a 90-day simulation, on 5 selected non–user-settable parameters (rows), and on select interventions (colors). The outcomes, columns from left to right, are total number of symptomatic infections, total number of production worker-shifts missed, effective reproduction number, and total cost in US$. The parameters examined, rows top to bottom, are *ψ* (“SEVERE_MULTIPLIER” denoting a parameter scaling the fraction of symptomatic infections that become severe); *μ*_IM_, and *μ*_IS_ (mean duration of mild (“duration_IM_mean”) and severe (“duration_IS_mean”) infection, respectively); *β*_IM_ (“p_trans_IM” denoting relative probability of transmission, per potentially infectious contact, during mild infection); and *φ* (“fraction_ssp_symptomatic” denoting a parameter that pertains to how much protection from developing symptomatic disease Recovered and Hybrid immunity provide to infected individuals). The interventions examined were Baseline (black), Temperature Screening (dark blue), Virus Test at *p* = 0.3 / working day (red), Physical distancing/Biosafety at −40% R₀ (dark green), and Vax + Boosting at *p* = 0.02/day (yellow). These colors are the same as those in Figures 2**, 3,** and **4**, although the line type may vary. The x-axis of each plot represents the *multiplier* applied to the parameter in question (i.e., 0.5, 1, or 1.5 for all of the parameters depicted here).

Multivariable analysis of the interaction of these 5 most sensitive parameters (*μ*_IM_, *β*_IM_, *φ*, *ψ*, and *μ*_IS_) and the same intervention and setting parameters that were evaluated in scenario analysis revealed that the uncertainty in the relative per-contact probability of transmission (*β*_IM_) during mild symptomatic infection affects the predictions of production loss when viral testing is moderate (*p* = 0.3/work day) and both prior vaccination and prior recovery rates are low (**Figure S13**). This is expected since small *β*_IM_ means the probability of infection transmission during mild infection is reduced and hence, at a given *R_0_*, more transmissions occur from asymptomatic or presymptomatic infections. Hence, more transmissions occur relatively early in infection, when a moderate rate of viral testing is less likely to have detected the infected individual. To improve the ability of the algorithm we used to detect relatively small effects, we reran the same analysis with only sensitivity and intervention parameters (**Figure S14**). This resulted in detection of an additional effect on production losses, of the mean duration (*μ*_IM_) of mild symptomatic infection. The worst outcomes were seen when both *μ*_IM_ and *β*_IM_ were low. This is also expected; like a small *β*_IM_, a short *μ*_IM_ tends to shift transmission earlier in infection (both by increasing the fraction of transmissions that are from asymptomatic and presymptomatic infection, and by shifting transmissions that still come from (a shortened) mild infection earlier within that phase. This means that it is harder for moderate virus testing to catch infection in time to prevent secondary transmissions. To investigate this further, we searched for setting-intervention scenarios where the 5 most sensitive uncertain parameters had the most impact on predictions and identified the scenario for a facility with no vaccinated and recovered employees and with shared housing and moderate viral testing (**Figure S15**). Here, *β*_IM_ alone affected unavailability and production loss predictions, and it interacted with *μ*_IM_ when impacting the symptomatic infection predictions. This means, as expected, that symptomatic infections, unavailability, and production losses are all worse when there are more transmissions during asymptomatic or presymptomatic infections.

## Discussion

This study presents FInd CoV Control, an ABM for assessing COVID-19 transmission and control in the food industry. The model can be customized to produce farm or processing facility settings, and employees’ housing types, vaccination and infection histories and age characteristics. Additionally, the model assesses different interventions, evaluating them counterfactually with regard to public health and economic outcomes and interpreting predictions at the population and individual operation levels. The two strongest themes in our results are bimodality and tradeoffs. The model also provides insights into the effectiveness of different interventions and main knowledge gaps. The model is expected to facilitate the food industry’s resilience and responsiveness to COVID-19 and similar future outbreaks, as well as to help navigate tradeoffs between public health and economic impacts of infection control interventions in this essential sector.

### Bimodality and variability in outcomes and intervention effectiveness

The simulation runs of FInd CoV Control can be interpreted to represent a population of food operations with similar workforces and work environments. The simulation predicts how an outbreak would unfold in each operation (i.e., run) following infection introduction compared to counterfactual versions of the same operation that implemented different interventions. This allows us to interpret the predictions at the population level, answering questions such as: “What fraction of similar food operations would experience certain health and economic outcomes?” Not only are most outcome distributions bimodal, but the counterfactual effects of most interventions are bimodal as well.

Relatively ineffective (on average) interventions (e.g., temperature screening) not only sometimes appear to produce good outcomes, but also rarely produce strong positive effects in a counterfactual sense. This is a particular consequence of a broader phenomenon: Much of the positive effect (when there is one) of effective and ineffective interventions alike comes from their potential to control an outbreak at a very early stage, often before symptomatic infections emerge. This early window of opportunity presents a challenge for “reactive” interventions (i.e., those implemented after the detection of a first infected case); by the time food operation managers (or policy-makers) are aware of an outbreak, the best opportunity to control it has already passed. This further supports the value of proactive planning with tools like FInd CoV Control, perhaps quarterly or on a rolling 90-day basis, to improve preparedness against potential disease introduction.

Conversely, even fairly effective (on average) interventions can result in little or no effect, or even counterfactually worse outcomes (with low probability) in individual outbreaks. This can largely be attributed to the timing of infection, for individuals who are infected at some point in either case; this can have an impact both through chance occurrence of opportunities for secondary transmission at particular points in time, and through the increase in the probability of symptomatic infection that comes with increased time since last immunity event (i.e., last vaccination or last recovery from natural infection). Together, these possibilities, reflected in the bimodality in outcomes, call attention to the substantial real-world probability of misleading conclusions from anecdotal (sample size of one) observations in individual operations, reinforcing the importance of mechanistic, predictive models such as FInd Cov Control. Available data about COVID-19 spread in the food industry settings originate from limited observational studies [36, 37]. Our findings about the bimodality and variability in outbreak dynamics and intervention effectiveness emphasize the need for larger-scale data collection. However, given the cost, technical, and confidentiality-related obstacles to collecting such data, purely data-driven models are unlikely. Collecting confidential data on infection spread in a food operation for the operation’s private decision-making is, of course, encouraged. However, in isolation from equivalent data in other comparable operations, such data will have a limited value since the data will represent just one of many possible (chance-determined) ways an outbreak, with or without an intervention, has unfolded. This underscores the need to proactively develop infrastructure capable of rapidly building mechanistic or hybrid (e.g., combining ABM and machine learning [38]) models to guide infection control, policy-making, and socially acceptable decisions under urgency and sparse data conditions.

### Tradeoffs between health and economic impacts of interventions

One of the biggest tradeoffs highlighted in our results is cost vs. effectiveness, particularly regarding viral testing. Sufficiently intensive testing is highly effective at controlling transmission, but viral testing can be quite expensive, whether from the cost of test kits themselves, the cost of increased unavailability due to isolation of individuals who test positive, or both. Importantly, attempting to find a moderate level of testing that optimizes this tradeoff is not necessarily a productive approach—testing, but at an insufficient frequency to achieve reliable control can actually be more expensive than either more frequent testing or not testing at all. The level of testing at which this economic “worst of both worlds” occurs is one of the multiple aspects of intervention effects that is heavily dependent on the level of community transmission (modeled as individual housing that provided opportunity for acquiring infection within the community), further complicating the effort to select an optimal approach. It is helpful to realize the multiscale nature of infection control in work environments deemed essential to society, where the interest is to control the infection and its effects at the worker individual level (to protect the individual employee’s health) and at the worker population level (to reduce infection spread and cost of control, and increase labor availability). These scale-related tradeoffs spill over into the tradeoff between costs of control (borne primarily by the company) and effectiveness (borne by both the company and individuals), leading to inefficiencies commonly faced in the private provision of public goods [39]. Designing public health policies that align operation incentives with desired public health outcomes is, therefore, critical to ensure the optimal provision of infection control interventions by operations. This tradeoff spills over into a broader challenge around both protecting essential workers and supplying the country with food. Thus, there is a need for more discussion around essential categories of industry and appropriate metrics for evaluating “costs of control”.

### Cost-effectiveness of counterfactual interventions

At an individual food operation level, FInd CoV Control can be used preemptively to ask questions such as: “Given the characteristics of the workforce and work environment in my facility, if an infected worker enters my facility in the near future, how likely it is that we will experience an outbreak?” “If we have an outbreak, how likely (in terms of the measures of central tendency and variation) are health and economic impacts under different intervention scenarios?” FInd CoV Control evaluates cost-effectiveness of 12 intervention scenarios and a no-intervention scenario. As seen in Figures 2 and **3**, even intensifying vaccination in an already moderately vaccinated population can yield modest but meaningful benefits. For proactive control of COVID-19 in the food industry, maintaining a vaccinated and boosted workforce to be prepared for a new outbreak remains a cost-effective intervention (albeit not sufficient to make other interventions unnecessary). Vaccination uptake can be increased by removing convenience and confidence barriers and leveraging workers’ motivation to protect self, family, and community [40]. Our model findings resonate with perceptions of the food industry’s leadership. Certain companies, particularly those in the labor-intensive meatpacking sector, took proactive measures by mandating vaccinations for their workforce [41–43] to prevent infection among employees and potential plant closures that were prevalent at the beginning of the COVID-19 pandemic [44]. In situations where vaccine mandates were not in place, other strategies such as physical distancing requirements and quarantines were implemented; however, these interventions were reported to lead to high worker absenteeism and hindered the efficient operation of processing plants [1].

Strategies like screenings for the disease were valuable in controlling workplace transmission, but also had serious limitations regarding reliability of the results and posed challenges due to being labor-intensive and costly compared to simpler strategies like using face coverings and practicing personal hygiene [45]. These observations in the food industry match the findings in our model about the effectiveness of screening, testing, and physical distancing/biosafety strategies in preventing symptomatic cases, albeit with high expenses and production losses associated with testing strategies. Effectiveness can also trade off against “costs” that are not strictly monetary. For example, even very intensive physical distancing and biosafety measures may be cost-effective, but some aspects of such measures (e.g., masking and face shields) can be highly unpopular in the long run (and even limit productivity in harsh work environments, such as extreme cold, hot and wet/damp), especially given the previous observation of the limitations of *reactive* interventions [46]. A comparison between the effects of highly and moderately effective interventions in this category supports the idea that, if one is going to implement masking and face shields, the addition of ventilation improvements is likely to be cost-effective. Our model revealed that predictions about intervention effectiveness are highly sensitive to the degree of community transmission. This emphasizes the importance of interpreting the effectiveness of work-based interventions in the light of the disease epidemiology in the community. This also emphasizes the importance of mitigating disease spread outside of work; however, this is particularly challenging for agricultural workers that are not stationary and typically share housing and transportation, allowing for easy employment-related transmission of the virus [47, 48].

### Limitations and future directions

The findings in this study are subject to model limitations. Major simplifying assumptions were that all workers live in the same type of housing (either individual or shared), and that employees in individual housing do not to transmit infection to their fellow employees during non-working shifts, but they are exposed to community transmission, while employees in shared housing spread infection to each other during non-working shifts but do not get exposed to community transmission. These assumptions represented “limiting cases” of a continuous spectrum of relative level of off-work contact with coworkers vs. others. While these assumptions allowed us to emphasize typical behaviors in these two housing settings, they limited the model’s generalizability to other real-world situations. The empirical support for model parameters varies, raising concerns, especially for those highlighted in the Sensitivity analysis (Figure 6). In this regard, a critical knowledge gap is the magnitude of long-lasting immunity provided by boosting (e.g., parameter *φ*), given a combination of high sensitivity and changing virus strains. Another important knowledge gap is the per-contact probability of transmission (*β*_IM_) during mild symptomatic infection relative to the transmission during presymptomatic and asymptomatic infection, which is particularly influential under moderate viral testing regimes (**Figures S13, S14** and **S15**). Nevertheless, the relatively weak effects of sensitivity parameters, compared to scenario parameters and intervention effects can be considered a good sign for the usefulness of this model – the strongest effects are those of parameters that either are designed to be easily set by end users (scenario parameters), or are presented to end users as one of several possibilities for actions (intervention effects), with background assumptions (sensitivity parameters), while not altogether ignorable, tending to play a weaker role. We suggest three improvements to address the model’s structural limitations: (i) Replacement of the current discrete-staged model of infectiousness over time with continuous infectiousness curves, analogous to how we model the continuous immunity trajectories; (ii) Continued improvement of our model of immune effects, immune boosting, and immune waning, as well as accounting for the changes in vaccination guidelines; and (iii) Incorporating multiple simultaneous interventions (starting at different times) and incorporating mixed individual and shared employee housing with varying exposures to community transmission. More broadly, we hope to further increase the modularity, flexibility, and ease-of-use of the model, and standardize model descriptions, to facilitate easy modification to address other respiratory pathogens or other critical infrastructure sectors. Towards these goals, we created a user-friendly web interface for an early version of FInd CoV Control, which allows the user to customize it to the characteristics of their workforce and generate easily interpretable confidential predictions to support decision-making [49]. Finally, future research could extend this modeling framework to food industry settings not covered in this study, including livestock farms and hospitality operations, as well as to modeling food trade between locations.

## Materials and Methods

### Employee population module

We model a heterogeneous population of agents (employees) with a variety of attributes reflecting both their current state and certain aspects of their personal history. Agent attributes set at the simulation start are explained below and summarized in **Table 1**.

### Age

Agents are randomly assigned an age category, with probabilities derived from industry-wide data about the age of agricultural workers [50]. This age category is then used to determine their probabilities, in the absence of immunity, of experiencing symptoms or dying (**Table S3**).

### Immunity-related attributes

All of an agent’s attributes that are directly relevant to that agent’s immunity, and that are not a consequence of their age, represent acquired immunity (whether complete or partial) to SARS-CoV-2 infection and COVID-19 disease. These attributes are associated with specific past events that created or boosted that agent’s immunity; for ease of reference, we refer to these simply as *immunity events* (**Table S4)**. Thus, all agents had an immune status of fully Susceptible (S) and a vaccination status of Not Vaccinated (NV, also referred to as unvaccinated) at the start of the COVID-19 pandemic, but some may have different values for one or both of these attributes at the start of the *simulation*.

Immunity-related attributes include the time (*t*_last, i_) of the *most recent* immunity event, the time (*t*_R, i_) of the most recent recovery from natural infection (if any), an “immunity trajectory” (*C*_i_, representing a trajectory defined by a combination of vaccination status and whether the agent has previously recovered from natural infection), and what the agent’s level of immunity was immediately prior to their most recent immunity event (*P*_last, E, i_, *P*_last, IP, i_, and/or *P*_last, IS, i_) (**Table 1**). Together, these determine an agent’s Protection against Any Infection (*P*_E, i_), Protection against Symptomatic Infection given Any Infection (*P*_IP | E, i_), and Protection against Severe Infection given Symptomatic Infection (*P*_IS | IP, i_) at the present time (**Table 1** and **Text S3B**). Each immunity trajectory includes curves for *P*_E, i_, *P*_IP | E, i_, and *P*_IS | IP, i_. These may include an initial increase in immunity, referred to as “ramp-up”; an initial period of total immunity; and/or immune waning. During both ramp-up and immune waning, *t*_last, i_ is relevant; during complete immunity (for immunity trajectories R, HV1, HV2, and HB only), *t*_R, i_ is relevant (specifically, in determining that there *is* complete immunity); and during ramp-up, *P*_last, E, i_, *P*_last, IP, i_, and/or *P*_last, IS, i_ are also relevant. These attributes are discussed further in the section “Immune dynamics” and in **Text S3B** and **Tables S6-S9**.

### Vaccination History

We assume, for the sake of simplicity, that all vaccination and boosting uses the monovalent Pfizer vaccine. Therefore, vaccination history includes the number of doses of vaccine the agent has received (i.e., whether they are unvaccinated (NV), partially vaccinated (V1), fully (primarily) vaccinated (V2), or boosted (B)), and when they received each of their previous doses, if any (Figure 1B**.iv**). Generally, only the time of their most recent dose is relevant to the model dynamics (**Table S6**). The current version does not account for repeated boosting in the vaccination history.

### State of Infection

An agent’s current infection status can be: Not Infected (NI), infected, but not yet infectious (E, for “Exposed”), Asymptomatically Infectious (IA), Presymptomatically Infectious (IP), Mildly symptomatic (IM), Severely symptomatic (IS), Critically symptomatic (IC), or Dead (D) (Figure 1B**.iv**). Additionally, if that state is anything other than Not Infected, we record how long they have been in that state; together with their precalculated duration (see below) for that state, this determines how much longer they will remain in it before progressing, recovering, or dying.

### Initial setting of agent infection and vaccination history

Agents’ history of immunity events prior to the start of simulation is important for immunity (and immune ramp-up and waning), and their history of vaccination events specifically is important for their eligibility for future vaccination (both primary and boosting). Therefore, user input (**Table 2**), is used to determine what fraction of the population has and has not experienced various immunity events and when. We then randomly generate exact times in a simple fashion. This aspect of run initialization involves (i) current infection status initialization, (ii) infection history initialization, and (iii) vaccination history initialization. These aspects do not directly interact with each other, although infection history and vaccination history interact in their effects on immunity. The initial settings are described in **Text S2**.

### Disease transmission module

#### Course of infection

The main aspects of the course(s) of infection are summarized in Figure 1B**.iv**. Agents are categorized as Not Infected (NI); Exposed (E); Infectious (I); or Dead (D). Infectious agents are further divided by whether they currently have clinical disease (IA and IP vs. IM, IS, and IC), whether they will subsequently develop clinical disease (IA vs. IP), and/or how severe their symptoms are (IM vs. IS vs. IC). They are further categorized by their immunity trajectory – fully Susceptible (S); one of the Vaccinated trajectories (V1, V2, or B); Recovered (R); or one of the Hybrid (H) immunity trajectories (HV1, HV2, or HB) – and (for all immunity trajectories other than S) the times since their last immunity event (their entry or reentry into that immunity trajectory) and their last recovery, if any (see “Immune dynamics” section).

Infectible agents (i.e., agents whose infection status is NI, and whose susceptibility to infection (see “Transmission model” section) is greater than 0) can acquire infection with SARS-CoV-2 from a potentially infectious contact with an Infectious agent. The probability of such a contact resulting in an infection depends on the susceptibility to infection of the infectible agent (1 – *P_E, i_*) and the infectiousness of the infectious agent (*β_IA_*, *β_IP_*, or *β_IM_*). How contacts are made is described in the “Work environment module” section.

Upon infection, the formerly infectible agent enters the Exposed state (E), and after a period of time (*D*_E, i_), the Exposed agent becomes infectious (I). At this time, they enter one of two infectious states without clinical disease: either IA or IP, the latter of which is the first stage of the symptomatic path. The distinction between IP and IA reflects a substantial difference in their per-contact transmission rates [51].

All IA agents are assumed to recover following a period of time (*D*_IA, i_). Agents taking the symptomatic path progress through up to four stages: IP, IM, IS, and/or IC. All IP agents progress to the IM state after a period of time (*D*_IP, i_), but agents who are in IM, IS, or IC may, after the corresponding period of time (*D*_IM, i_, *D*_IS, i_, or *D*_IC, i_, respectively), either continue their disease progression to the next state on the symptomatic path or recover.

When an agent recovers, regardless of which infectious state they recovered from, their last immunity event time (*t*_last, i_) and Recovered immunity event time (*t*_R, i_) are both set equal to the current time, their infection status is set to NI, and their immune status is set to Recovered (R) if they have never been vaccinated (NV), or to the appropriate Hybrid state (HV1, HV2, or HB) otherwise. For agents in IC that do not recover, the next step is Death (D).

We do not explicitly model individual symptoms such as fever, cough, etc. However, for the temperature testing intervention, we do tacitly assume that fever is only present if the individual is symptomatically infected (IM, IS, or IC). We define “Severe” symptoms as those requiring hospitalization; consequently, we assume that only agents with an infection status of IA, IP or IM can transmit to their fellow employees, because agents with Severe or Critical symptoms are so sick that they require hospitalization. Relative transmissibility (per contact) is set based on the infection stage (**Table S5**). *Absolute* transmissibility has no effect in the model, as we set the average expected contact rate to achieve a specified basic reproduction number (*R*_0_) (**Text S3A**).

For agents in any of the infected states, disease progression is based on the duration of each state (**Table S2**), age-dependent baseline probabilities of entering each disease state during disease progression (**Table S3**), and immunity-dependent modification of those baseline probabilities.

#### Transmission model

Agents who are infectible (i.e., agents whose infection status is NI, and whose susceptibility to infection is greater than 0) can be infected by contacts with either infectious coworkers or infectious people outside of work, in the broader community (if housing is “individual”). Contact structures of the agents while at work were determined by the place of agents in the hierarchical structure of the farm or facility and their work schedule. To be more precise, for each shift type (e.g., weekday Production Shift 1, weekend Cleaning Shift, etc.), we have a matrix of expected contact rates between pairs of agents. For some combinations of a pair of agents and a shift type, a non-zero contact rate may represent contacts made at work; for others, it may represent contacts made in shared housing. Further details are in **Text S3A**.

#### Immune dynamics

We distinguish between 8 basic states (immunity trajectories) with respect to immunity: fully Susceptible (S), partially vaccinated (i.e., with one dose of a two-dose primary series) (V1), Fully Vaccinated (here defined as a 2-dose primary series; V2), Boosted (B), Recovered (R), and Hybrid immunity (H) with partial vaccination (HV1), with full vaccination (HV2), or with full vaccination and a Booster (HB). Non-hybrid vaccinated trajectories (V1, V2, and B) feature a smooth ramp-up from their individual’s previous level of immunity, that lasts for T_V1→V2_ = 21 days, T_ramp, V2_ = 14 days, or T_ramp, B, 1_ + T_ramp, B, 2_ = 14 days, respectively, counting from the time since the individual’s last immunity event (i.e., first vaccine dose, second vaccine dose, or booster shot, respectively). The non-hybrid Recovered trajectory (R) features an initial *T*_total, R_ = two months (61 days) period of total immunity, counting from the time of their (most recent) recovery. The Hybrid immunity trajectories (HV1, HV2, and HB) have characteristics of both vaccinated and recovered trajectories, and can be entered either by recovery following vaccination or by vaccination following recovery. Consequently, a particular individual’s experience of one of these trajectories may include either or both of total immunity and ramp-up. The transitions between these immunity trajectories are summarized in **Table S4**, and the equations for the protection they offer are given in **Table S9**. To model waning immunity, we assigned each agent three variables indicating the major factors influencing their level of susceptibility to both infection and progression: (i) the immune state that they entered at the time of their *last immunity event*, (ii) the time at which that event occurred (and hence, at any given subsequent point in time, how long it has been since that event), and (iii) the time of their *last recovery* from natural infection. The exceptions are agents whose immune state is fully Susceptible (S), whose time of last event is not defined, and agents whose immune state is either fully Susceptible (S) or one of the non-hybrid Vaccinated states (V1, V2, and B), whose time of last recovery is not defined. We then created functions (**Table S9**) giving, for any valid combination of state, time since entry, and time since recovery (and previous immunity, if they are currently in a ramp-up period), their level of relative protection from each of infection, symptomatic infection conditional on any infection, and severe infection conditional on symptomatic infection. We used exponential or exponential-mixture waning for long-term behavior of V2 and B (fitted from data in [52]), and logistic waning for long-term behavior of R, HV1, HV2, and HB (with parameters inferred from the tables in [53]), with some special case behavior at the start of states other than S (ramp-up and/or a period of complete immunity), to account for the delay in reaching full protection following vaccination, and to prevent unrealistic cycles of extremely rapid reinfection. This is further explained in **Text S3B**.

#### General model of vaccination

Agents’ vaccination status can be unvaccinated (NV), partially vaccinated (V1), fully vaccinated (V2), or boosted (B). This vaccination status directly corresponds to their immune status (with NV corresponding to fully Susceptible (S)) if they have never recovered from a natural infection; if they have, then their immune status is either Recovered (R), if they have never been vaccinated, or one of the Hybrid immunity trajectories (HV1, HV2, and HB).

#### Vaccination trajectories

Partially vaccinated agents become eligible to receive a second shot *T*_V1 → V2_ = 21 days after receiving their first one, and fully vaccinated agents become eligible to receive a booster shot *T*_V2→B_ = 5 months (treated as a deterministic 152 days) after their second shot [54]. We assume that all V1 agents enter V2 states as soon as they are eligible, but that only a fraction of V2 agents enters the B state as soon as they are eligible. We further assume that agents who are eligible to become V1/B at simulation start, but have not yet done so, will not become V1/B (respectively) during the simulation, in the absence of an intervention to promote primary vaccination/boosting, respectively.

#### Work environment module

In both farm and facility models, each week consists of 5 work days and 2 non-working days. Each calendar day is modeled as consisting of 3 eight-hour periods we call “shifts”, and, thus, shift (i.e., 8-hour time step) is the time unit of the model. Each agent spends two shifts awake, and one asleep. For simplicity, we tie the agent’s sleep cycle to their work cycle, so that each agent spends their first shift awake at work if it’s a work day. This structure is illustrated for a sample agent (one scheduled to work on the first shift of the calendar day) in Figure 1B**.i** and shown in greater detail in **Table S10**.

In the facility model, each working day consists of Production Shift 1, Production Shift 2 (which may actually be a non-working shift, if the facility in question only has one production shift per day), and a Cleaning Shift. In the farm model, all agents are scheduled to work on the same shift, which, by analogy with the facility model, we refer to as Production Shift 1. (Hence, by the same analogy, all farm agents are awake, but not working, during Production Shift 2, and asleep during the shift that would be the “Cleaning Shift” in the facility model.)

We distinguish between available and unavailable agents regarding SARS-CoV-2 transmission. Available agents are available to work their scheduled shifts – meaning that they can do work (relevant in the economic analyses), and can also potentially infect, or be infected by, other agents. Agents are available by default, but may become unavailable to work (and, subsequently, may become available again).

In the baseline (no intervention) model, unavailable agents are limited to those who are either hospitalized (i.e., those who have an infection status of IS or IC) or dead (those who have an infection state of D). Hospitalized agents become available again upon recovery. Under certain interventions, not hospitalized agents may become diagnosed and isolated; these remain unavailable until they are deisolated (**Text S3C**).

We model a facility or farm with a hierarchical organization, although the details of this hierarchy differ somewhat between the facility model and the farm model. This hierarchy is assumed to be fixed over the time horizon of the model, as is the associated work schedule. The expected number of contacts between pairs of workers who are both “available” (i.e., not isolated, hospitalized, or dead) is likewise held constant (i.e., we do not reassign workers between work crews or production lines based on other workers’ absences). To make the interface more manageable, we assume that the structure of the operation is “regular” in the sense that if there are two or more of the same sub-structure (e.g., two production shifts, two or more teams of work crews, two or more work crews within a team, or two or more production lines), then each of those sub-structures has the same structural characteristics (e.g., if one work crew consists of 10 workers and a foreman, than all work crews consist of 10 workers and a foreman). Features specific to the farm and facility models are described in **Text S4A** and **S4B**, and assumptions common to both models are in **Text S4C**.

#### Interventions

Apart from the baseline (non-) intervention, we modeled 12 interventions (introduced in **Results** and described in detail in **Text S3C)** and performed counterfactual comparisons of the no-intervention baseline and 12 interventions (Figure 1A). To make these comparisons more precise, on a run-by-run basis, we (a) reseed the pseudorandom number generator (pRNG) with the same value before processing each intervention and (b) make use of the pRNG in such a way as to ensure that two runs that start with the same pRNG state and that differ only in the factors that we allow to vary between possible interventions under a given scenario, will have the same pRNG state at all analogous points thereafter. The ABM model and all analyses were implemented in R software (version 4.0.4) [55]. Model running is described in **Text S13**.

### Model outcomes

Our outcomes of interest can be broadly grouped into those which pertain primarily to public health, and those which pertain primarily to business disruptions and economic impact. We compare these outcomes both across interventions within a setting scenario and across different scenarios.

#### Public health outcomes

Our primary public health outcome of interest is the total number of symptomatic SARS-CoV-2 infections that occur throughout a run. We define this as the number of occasions of an agent transitioning from the IP to IM state between the start of simulation (*t* = 0) and the end of simulation (*t* = *T*). This includes transitions into IM of agents who were in the E state at simulation start, if they transition from E to IP (and, subsequently, to IM) rather than to IA, but does not include the transitions into IM *before t* = 0 for agents that were in IM at simulation start. It also does not include the transitions into IM *after t* = *T* of any agents who are in E or IP at simulation end. If the simulation length and transmission dynamics are such as to result in some agents transitioning into IM more than once during the simulation, then we count each such transition separately; hence, it may be possible, depending on user-set parameters, to have a number of symptomatic infections that exceeds the number of agents. An additional outcome of interest is the number of total SARS-CoV-2 infections (i.e., including asymptomatic infections) that occur throughout a run. We present public health outcomes, for each intervention, as a curve of the mean over time, as well as violin plots summarizing the cumulative distribution over a defined planning horizon, to allow both comparison of expectations and understanding of the variance.

#### Economics outcomes

The main dimensions considered for economic analysis are: (i) direct costs of performing an intervention, and (ii) the productivity costs or benefits of the intervention. To assess (i), we add certain details to our hypothetical interventions, based on how interventions are implemented in real life.

Relative to a “no-intervention” baseline of doing nothing, the direct costs of interventions can be expected to always be greater than or equal to zero. Each intervention is compared to the baseline to estimate direct costs of performing each intervention. Calculations of the costs of interventions are described in **Text S5**. To answer (ii), we estimate the productivity loss based on the worker absences from the infection model with a Cobb-Douglass production model. The Cobb-Douglass production function takes the general form

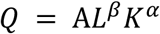

where operations use labor *L* and capital *K* to produce output *Q*, and *A* is some constant. The output elasticities of labor and physical capital are *β* and *α*, respectively. In choosing output level *Q*, operations face the following cost function (abstracting from fixed costs):

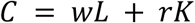

where *w* is the wage rate and *r* is the rental rate for capital. Profit-maximizing operations operate at the efficient production frontier in equilibrium.

Based on a previous survey of producers (targeting fresh produce growers and processors, and dairy, beef, pork processors) [17], we assume that the facility can maintain full production (*Q*_*f*_) with up to 15% of absenteeism without incurring appreciably higher production costs. If an operation can reduce its “full output” labor input allocation *L_f_* to a short-staffed model where *L*_*s*_ = 0.85*L*_*f*_ without reducing full output (*Q*_*s*_ = *Q*_*f*_) in a costless way, the operation must be able to substitute enough additional capital above the current “full output” allocation in the short run, such that the cost of increased capital is completely offset by reduced labor costs. To capture this short-run flexibility, we therefore assume that the current equilibrium is not unique, but instead one in a set of cost-minimizing equilibria where 0.85*L*_*f*_ ≤ *L* ≤ *L*_*f*_.

If labor absenteeism exceeds 15%, however, we assume that the capital allocation now remains fixed at the (higher) 15% absenteeism point, and the production paradigm takes the previously described Cobb-Douglass form. Under that framework, since operations cannot substitute capital for labor, they will have to reduce output based on these labor shortages. In an empirical work application of productivity analysis in the food and agricultural sector in the US, Ahmad [56] estimates a Bayesian stochastic Cobb-Douglas production function using US state-level agricultural data from 1960-2004 (n=2,160). He estimates *α* = 0.316 [0.271, 0.362] and *β* = 0.437 [0.392, 0.483]. Thus, production is

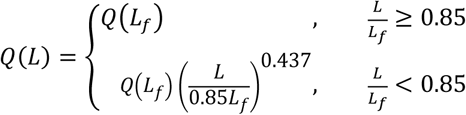

Full production quantity is provided by the users, and estimated production loss is simply

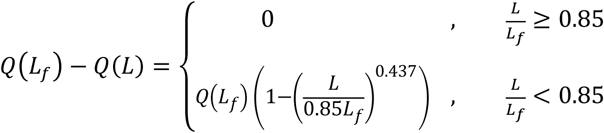

The results of the above analyses are summarized by the following economic outcomes: the mean number of employees unavailable to work production shifts over time; violin plots summarizing the distribution of the cumulative number of production worker-shifts missed; fraction of shifts short; and total direct expenses, production losses, and total costs associated with an intervention (US$).

#### Scenario analysis

We tested scenarios corresponding to factors: “setting” (“farm” vs. “facility”), “housing” (“individual” vs. “shared”), “vaccinated” (“high” vs. “none”), and “recovered” (“high” vs. “nothing”). In a full factorial analysis approach, we tested all 16 combinations of these four factors and for each scenario we ran the no-intervention baseline and all 12 interventions. We then constructed regression trees, using the R package rpart (version 4.1.19; using default values for all control parameters) for outcomes (symptomatic infections, worker-shifts unavailable, production losses and intervention expenses) vs. the four scenario parameters and the 5 intervention parameters defining our 12 interventions (**Table S15**). We present a subset of results for which setting is “facility,” for reasons discussed in the “Scenario analysis” in the Results section.

#### Sensitivity analysis

We evaluated the parameter sensitivity for four outcomes using a One Factor at a Time (OFAT) approach. Three outcomes were the total symptomatic infections, total number of worker-shifts missed and total cost over the simulation length. The fourth outcome was the effective reproduction number (*R_eff._*) at the start of simulation. Because of the large number of parameters to be examined, under (initially) a variety of scenarios, we averaged results over batches of 100 runs each, rather than the 1,000 runs that we use in most other contexts. In general, for a given parameter whose value in our model was *x*, we examined results when that parameter was set to 1/2 *x*, *x*, and *2x* (**Table S16**). For 2 parameters (*T*_V2→B_ and *T*_1st V2 -> 0_), one of these values was impossible, given a requirement that it be possible for some individuals to be eligible for boosting at simulation start. For these parameters, we set the relevant value to the most extreme possible value in the same direction (i.e., 212.5 days for (increased) *T*_V2→B_ and 305 days for (reduced) *T*_1st V2 -> 0_).

To estimate *R_eff._* at the start of simulation, we modified the model so that, when an employee was infected, instead of their infection status being changed from Not Infected to Exposed, their immunity was set to Recovered, and their *t*_last, i_ was reset to the current time. Conceptually, this amounts to their “skipping over” any infected states, and going directly from infection to recovery. This ensured that all employee-to-employee transmissions would necessarily be from employees who were infected at the start of simulation. We then ran the model starting with a single Exposed employee (as is the default), and observed the number of employee-to-employee transmissions that occurred. The average of that number across multiple runs was then used as an empirical estimate of *R_eff_*. In scenarios with “individual” housing, the community-acquired infection was allowed to occur (likewise transferring infected employees directly into a Recovered state), but was not included in the count of employee-to-employee transmissions measured by *R_eff_*.

We initially examined the results of sensitivity analysis using default user-settable parameters, except as noted, across the same range of settings as we initially considered for scenario analysis. We found that, for most parameters, sensitivity was higher in “high” settings (i.e., with a history of both vaccination and recovery from natural infection) than in historical or pseudo-historical settings, higher in facility settings than in farm settings, and higher in settings with shared housing than in settings with individual housing. For this reason, we focused our further examination on the “facility” setting, with all parameters default except for housing, which was “shared” instead of “individual” (with physical distancing in shared housing at the default level). Looking back at scenario analysis, this is equivalent to the scenario in which both “vaccination” and “recovered” were “high.”

For each parameter-intervention-outcome combination, we obtained three mean outcome values, as described above. We divided the largest of those three values by the smallest to obtain a simple measure *ω* of the sensitivity that could address both monotonic and non-monotonic effects. For each parameter, we then chose the largest such measure (*ω_max_*) across all intervention-outcome combinations. We then selected for depiction and discussion in the Results section those parameters for which *ω_max_* ≥ 2 (but all results are shown in **Figs. S5-S8**). As the only exception to this process, we omitted the baseline no-intervention scenario in the Total Cost outcome, as the sensitivity measure for this outcome-intervention was generally undefined or (rarely, and seemingly at random) infinite, due to the extreme rarity of non-zero Total Cost at baseline.

Finally, the most sensitive uncertain parameters identified in OFAT analysis were evaluated in multivariable analysis to determine how the uncertainty in these parameters interacts with settings and intervention parameters when affecting predictions. To this end, we represented the most sensitive parameters with Uniform probability distributions with ranges identical to those evaluated in OFAT analysis for the respective parameter. For each of the eight combinations of scenario parameters analyzed in scenario analysis (i.e. combinations of “housing” (“individual” vs. “shared”), “vaccinated” (“high” vs. “none”), and “recovered” (“high” vs. “nothing”)), we randomly selected 50 sets of sensitivity parameters using Monte Carlo simulation allowing the sensitivity parameters to vary simultaneously within their Uniform distributions. For each of the scenario x sensitivity parameters combinations, we ran 100 runs of the model for each of the 13 interventions, resulting in a simulation with 8 x 50 x 13 x 100 = 520,000 iterations in total. The simulation results were evaluated using regression trees as in scenario analysis with the goal of determining how the uncertainty of sensitive parameters interacts with (i) setting parameters and (ii) intervention parameters, independently and combined, or in other words, how robust are model predictions about the impact of settings and interventions in the face of parameter uncertainty. Additional data used in model development are in the Supplementary information file (and references therein [57–69]).

## Data Availability

All data are available in the main text or the supplementary information. R code is available under GPL-2.0 license at https://github.com/IvanekLab/covid-journal-code

## Acknowledgments

This study was funded by the Agriculture and Food Research Initiative from the USDA National Institute of Food and Agriculture (NIFA), Competitive Grant no. 2020-68006-32875 (RI, AA, SA, MW). Additional support was provided by the Cornell Institute for Digital Agriculture (CIDA) (RI, MW) and the Artificial Intelligence Institute for Next Generation Food Systems from the USDA NIFA, grant 2020-67021-32855 (RI, MW). The funders had no role in the study design, data collection, and analysis, decision to publish, or preparation of the manuscript. The authors thank the Industry Advisory Council, comprised of executive-level leaders in the major US produce farms and food processing companies and an official in the Centers for Disease Control and Prevention, which was assembled to guide modeling work presented here, for valuable feedback on the structure of FInd CoV Control. The authors also thank Ms. Yu Tang (Dyson School of Applied Economics and Management, Cornell SC Johnson College of Business, Cornell University, Ithaca, NY, USA) for her contribution to the methodology, investigation and writing of the original draft for the evaluation of economic impacts of interventions.

## Author contributions

Conceptualization: CH, RI, AA

Methodology: CH, RI, AA, CZ, DW

Investigation: CH, RI, EB, SIM

Visualization: CH, RI

Supervision: RI, AA

Writing—original draft: CH, RI

Writing—review & editing: CH, EB, SIM, CZ, AA, DW, MW, SA, RI

## Supplementary Information

Supplementary Information is enclosed.

