## Supplementary Information for "An agent-based model of COVID-19 in the food industry for assessing public health and economic impacts of infection control strategies"

[2]iFoodDecisionSciences, Seattle, WA, USA

[3]Nolan School of Hotel Administration, Cornell SC Johnson College of Business, Cornell University, Ithaca, NY, USA

[4]iDecisionSciences, Seattle, WA, USA

[5]Department of Food Science, College of Agriculture and Life Sciences, Cornell University, Ithaca, NY, USA

***** Renata Ivanek

### Contents

An agent-based model of COVID-19 in the food industry for assessing public health and economic impacts of infection control strategies ... 1

Supplementary text ... 4
- Text S1: Definitions, with abbreviations in parentheses ... 4
- Text S2: Employee population ... 7
- Text S3: Disease transmission module ... 8
- Text S4: Work environment module ... 14
- Text S5: Interventions cost ... 17
- Text S6: Model validation ... 18
- Text S7: Insights about farm, large facility and stochastic die-off ... 19
- Text S8: Additional results from counterfactual comparison of interventions ... 20
- Text S9: Scenario analysis ... 21
- Text S10: Sensitivity analysis ... 22
- Text S11: Derivations of defaults for initial fractions ... 23
- Text S12: Derivation of coefficients for logistic decay formulas ... 24
- Text S13: Model running ... 24

Supplementary figures ... 25
- Fig. S1. Schematic representation of the agent number and hierarchy (left) and agent contact network and relative contact rates (right) in the work environment module for a produce farm. ... 25
- Fig. S2. Illustration of public health outcomes for a large facility (1,003 employees) for baseline (no intervention), for each of the interventions in absolute terms as well as relative to the baseline ... 27
- Fig. S3. Illustration of unavailability for a large facility (1,003 employees) for baseline (no intervention), for each of the interventions in absolute terms as well as relative to the baseline ... 29
- Fig. S4. Illustration of costs for a large facility (1,003 employees) for baseline (no intervention) and each of the interventions ... 30
- Fig. S5. Plot of sensitivities of the number of symptomatic infections with respect to all sensitivity parameters ... 32
- Fig. S6. Plot of sensitivities of the number of worker-shifts unavailable with respect to all sensitivity parameters ... 33
- Fig. S7. Plot of sensitivities of the total cost with respect to all sensitivity parameters ... 34
- Fig. S8. Plot of sensitivities of the effective reproduction number with respect to all sensitivity parameters ... 35
- Fig. S9. Regression Trees for a variation of the model in which the presence (“True”) or absence (“False”) of community transmission and dormitory transmission are allowed to vary independently ... 37
- Fig. S10. Illustration of public health outcomes for baseline (no intervention) and each of the interventions in absolute terms as well as relative to the baseline, for a scenario with both community and dormitory transmission ... 39
- Fig. S11. Illustration of unavailability for baseline (no intervention) and each of the interventions in absolute terms as well as relative to the baseline, for a scenario with both community and dormitory transmission ... 41
- Fig. S12. Illustration of costs for baseline (no intervention) and each of the interventions, for a scenario with both community and dormitory transmission ... 42
- Fig. S13. Regression Trees for multivariable analysis of setting and intervention parameters and the five most sensitive uncertain parameters (from Figure 6) ... 44
- Fig. S14. Regression Trees for multivariable analysis of intervention parameters and the five most sensitive uncertain parameters (from Figure 6). ... 46
- Fig. S15. Regression Trees for multivariable analysis of the five most sensitive uncertain parameters (from Figure 6) in a particular setting-intervention scenario for a facility with no vaccinated and recovered employees and with shared housing and moderate viral testing ($p$ = 0.3/work day). ... 48

Supplementary tables ... 50
- Table S1. Numbers of agents with various immune states and histories derived from user-set parameters ... 50
- Table S2. Model parameters ... 52
- Table S3. Age categories and associated probabilities for stages of infection ... 53
- Table S4. Immunity events and their effects ... 54
- Table S5. Transmissibility by infection stage ... 55
- Table S6. Immunity curve symbols, definitions ... 56
- Table S7. Immunity curve symbols, additional notation ... 57
- Table S8. Explanation of “directly-parameterized” immune trajectory components ... 58
- Table S9. Calculation of relevant immune trajectory components ... 59

### Supplementary text

#### Text S1: Definitions, with abbreviations in parentheses

**Asymptomatic/Asymptomatically infected (IA)**: The infection status of a person who is infectious but shows no symptoms and will not develop any symptoms later. An individual in IA can transmit the infection to an infectible person, but with a lower probability of transmission per contact than an individual whose infection status is either Presymptomatic or Mildly symptomatic.

**Critical/Critically symptomatic (IC)**: The infection status of a person who requires hospitalization and whose next stage of disease progression, if they do not recover, is Death. The Critical infection status is distinguished from merely Severely symptomatic infection (which also requires hospitalization) by this imminent possibility (whether realized or not) of death. Although an individual in IC is infectious, they cannot transmit the infection *to their coworkers* because they are removed from the workforce.

**Community transmission**: Transmission from individuals who are not employees of the modeled operation. SARS-CoV-2 transmission in a community can be High, Intermediate or Low.

**Death (D)**: If an individual whose infection status is Critically symptomatic (IC) does not recover, the next step of disease progression is death (D).

**Exposed (E)**: The infection status of an individual who is infected, but is not yet infectious (i.e., is currently unable to spread the infection).

**Hospitalized**: Describes individuals whose infection status is Severe or Critical. We assume that hospitalized individuals cannot practically spread the disease to their coworkers, as they are removed from the workforce.

**Immunity Trajectory**: A property of an individual that is defined by a combination of their vaccination status and whether they have previously recovered from natural infection, and which, in combination with their time since last immunity event and (if applicable) time since last recovery, determines their level of complete or partial immunity to infection and/or progression to more severe phases of infection. We will sometimes refer, for convenience, to an individual who has an infection status of, for example, Recovered (R), as "being" Recovered or "being in" or "being on" R, and analogously for other immunity trajectories.

**Infected**: Describes individuals whose infection status is anything other than Not Infected or Dead (i.e., any of Exposed, Asymptomatic, Presymptomatic, Mild, Severe, or Critical).

**Infectible**: A person who is susceptible to infection and thus capable of becoming infected. This includes all individuals whose infection status is Not Infected and either:

- Their immunity trajectory is Susceptible (S), Partially vaccinated (V1), Fully vaccinated (V2), or Boosted (B); or
- Their immunity trajectory is Recovered (R) or one of the Hybrid trajectories (HV1, HV2, or HB), and the time since their last recovery is greater that $T_{total,\ R}$.

**Infection Status**: An individual's state of being uninfected (NI), infected and in a certain phase of infection (E, IA, IP, IM, IS, or IC), or dead (D). We will sometimes refer, for convenience, to an individual who has an infection status of, for example, Critical (IC), as "being" Critical or "being in" IC, and analogously for other infection statuses.

**Infectious:** Describes individuals who are Infected and whose infection status is not Exposed (i.e., individuals with an infection status of Asymptomatic, Presymptomatic, Mild, Severe or Critical). All such individuals can transmit the virus, and so are formally classified as infectious, but only those who are Asymptomatic, Presymptomatic, or Mild can transmit *to their coworkers*, as those who are Severe or Critical are assumed to be hospitalized (and therefore removed from the workforce).

**Infectious contact**: A contact between an infectible and an infectious individual that results in transmission of the disease to the infectible, which causes the infectible individual's infection status to become Exposed.

**Mild/Mildly symptomatic (IM)**: The infection status of an infected individual who has developed symptoms, but is not sick enough to require hospitalization. Follows the Presymptomatic stage. The Mildly symptomatic stage is the first stage where individuals start to show symptoms. Mildly symptomatic individuals can transmit the disease to infectible individuals, with a probability per contact that is higher than for Asymptomatic individuals, but lower than for Presymptomatic individuals.

**Presymptomatic/Presymptomatically Infected (IP)**: The infection status of a person who, though infectious, does not have any symptoms yet, but who will eventually develop symptoms later. An individual who is Presymptomatic can transmit the virus to an infectible person, with a higher probability per contact than an individual who is either Asymptomatic or Mildly symptomatic.

**Individual housing**: A model of contact patterns outside of work and transportation to and from work that reflects an operation having individually-housed employees. Unlike employees in shared housing, workers in individual housing are assumed not to transmit to their fellow employees during non-working shifts, but they are exposed to community transmission.

**Recovered (R)**: The immunity trajectory of a never-vaccinated person who has been infected with SARS-CoV-2, but who is not currently infected. Recovered individuals are not at risk of being reinfected for the first $T_{total, R}$ days after recovery, but are at risk thereafter, and that risk increases with the time since their last recovery from natural infection.

**Hybrid immunity (HV1, HV2, HB)**: A collective label for the immunity trajectories of any individuals who have recovered from natural infection and then received vaccine, or who have been vaccinated and subsequently got infected and then recovered from infection. This is divided up into three more specific immunity trajectories, HV1, HV2, and HB, which apply to individuals with Hybrid immunity whose vaccination status is V1, V2, or B, respectively. Individuals with any of these immune trajectories are at risk of being reinfected if the time since their last recovery is greater than or equal to $T_{total, R}$, and that risk increases (after a possible relatively brief ramp-up) with the time since their most recent immunity event.

**$R_0$**: Symbol for the **basic reproduction number**, a mathematical term that indicates how "spreadable" an infectious disease is in a given population, in the absence of any immunity or (non-negligible) prevalent infection. For example, if a disease has an $R_0$ of 3, a person who is infected would be expected to transmit it to an average of 3 other people if everybody in the population had an immunity trajectory of fully Susceptible (S), and everyone except for that one person had an infection status of Not Infected (NI). In the deterministic limit of a sufficiently large population:

- If $R_0$ is less than 1, each infected individual will, on average, causes less than one new infection during their infection. In this case, the disease will decline and eventually die out.
- If $R_0$ equals 1, each existing infected individual will cause, on average, one new infection during their infection. The disease is present, but there will not be an outbreak of disease that infects a non-negligible fraction of the population.
- If $R_0$ is more than 1, each existing infected individual on average causes more than one new infection. The disease will be transmitted between people, and there will be a non-negligible outbreak of disease.

In a population of finite size, and with stochastic transmission dynamics, these outcomes are not necessarily *guaranteed*, but generally provide a reasonable idea of what is most likely to happen.

**$R_{eff.}$:** Symbol for the **effective reproduction number**, a mathematical term that indicates how "spreadable" an infectious disease is in a given population, under current conditions, potentially including non-negligible fractions of the population that are or have previously been infected.

**Run**: A single iteration of the model. Multiple runs (1,000 by default) are performed for each intervention, and the results summarized.

**SEIRS model**: A model for infection, disease progression, and recovery. In a simple SEIRS model, Susceptible (S) individuals become Exposed (E), then become Infectious (I), and finally become Recovered (R), after which they cannot become infected until they once again (due to immune waning, antigenic drift, or other factors) become Susceptible. In our model, this is elaborated slightly (as shown in **Figure 1C**):

- We have multiple age brackets, with different probabilities of infection and progression at baseline.
- We have multiple Infectious states (infection statuses of IA, IP, IM, IS, and IC), with various trajectories an individual can take through them.
- Infectible individuals are distinguished from one another by an immune trajectory (essentially a combination of vaccination status and history of natural infection), a time since their last immunity event (either their last vaccine dose or their last recovery), and a time since their last recovery.
  - The combination of immunity trajectory, time since last immunity event, and time since last recovery determines the degree of protection from both infection and progression.
  - As in a simple SEIRS model, individuals with immunity from a sufficiently recent natural infection (R), or with hybrid immunity from vaccination and a sufficiently recent natural infection (HV1, HV2, and HB), are completely immune, but this complete immunity is not permanent. In addition, we model continuous waning of immunity for both these individuals and those who have immunity only from vaccination (i.e., from full vaccination (V1 and then V2), or booster (B)), rather than an instantaneous transition back to complete Susceptibility.
  - Similarly, immunity trajectory and time since last immunity event affect the probability of transition from Exposed (E) to Presymptomatic (IP) (as opposed to Asymptomatic (IA)) and the probability of transition from Mild (IM) to Severe (IS) (as opposed to recovery into Not Infected, with an immunity trajectory of either Recovered (R) or one of the hybrid trajectories (HV1, HV2, or HB), as appropriate).
- Thus, instead of just Susceptible individuals (S), we have eight different immunity trajectories that infectible individuals can be on (fully Susceptible (S), Partially vaccinated (V1), Fully vaccinated (V2), Boosted (B), and, after enough time has passed, Recovered (R) and Hybrid (HV1, HV2 and HB)), and these infectible individuals differ in how likely they are to become infected (even within the same age bracket) following a potentially infectious contact based on both which of these trajectories they are on, and how long they have been on it.
- Similarly, Exposed individuals can be on any of these eight trajectories, and can have different probabilities of becoming symptomatic (even within the same age bracket), depending on which of these trajectories they are on and how long they have been on it. Likewise, these trajectories can affect the probability of individuals who are Mildly symptomatic becoming Severely symptomatic instead of recovering.

This model can still be considered a form of an SEIRS model, because the basic infectible-Exposed-Infectious-Recovered (and temporarily immune)-infectible again trajectory is the same, even if there are additional details.

**Severe/Severely symptomatic (IS)**: The infection status of a person who requires hospitalization, but is not (currently) at imminent risk of death. Although they are infectious, they cannot practically transmit *to their coworkers*, because they are removed from the workforce (by being hospitalized). They can either continue their disease progression, to an infection status of Critical, or recover (and enter the Not Infected infection status and either the Recovered immunity trajectories or one of the Hybrid immunity trajectories, according to their vaccination status) after a period of time.

**Shared housing**: A model of contact patterns outside of work and transportation to and from work that reflects an operation in which employees reside in employer-provided housing, where their interactions during non work-shifts and sleeping arrangements may allow them to transmit to other employees.

**Physical distancing/biosafety**: Any strategy of physical distancing or other biosafety measures used during traveling and at work, in order to reduce the probability of transmission from an infectious employee to an infectible employee (as distinct from both interventions designed to encourage vaccination and boosting, and interventions designed to detect and isolate infectious employees), such as spacing workers ≥6 ft, installed physical barriers, staggered break times, improved ventilation, etc.

**Symptomatic**: Describes individuals who are Infected and are showing symptoms. This encompasses three more specific states (infection statuses), characterized by (current) severity: Mild, Severe, or Critical. (All Presymptomatic individuals will (by definition) eventually enter the Mildly symptomatic stage, but not all will progress to the Severely or Critically symptomatic states.)

**Unavailable (to work)**: Describes an individual who would normally be scheduled to work a given shift, but who is unable to do so, due to being hospitalized or being in isolation as a result of a (true or false) positive result on a viral test or temperature screen. This means that they cannot infect or be infected by other employees, and it may also affect the output of production shifts.

### Text S2: Employee population

A. Initial infection status

We model a heterogeneous population of agents (employees) with attributes that represent past events and current state include age, immunity-related attributes, vaccination history, and the current state of infection, if any. In addition, certain random consequences of possible future events (e.g., how long a currently-uninfected agent will remain asymptomatically infected if they become infected and do not develop symptoms) are pre-calculated for convenience and consistency, and are recorded as agent attributes, even though they may not directly represent any current facts about the agent.

We select at random $N_E(0)$ agents to begin the simulation with an infection status of Exposed (E). For each, we set their time of entering that state ($t_{E,\,i}$) to be uniformly distributed on the interval ($-D_{E,\,i}$, 0), i.e., we make it equally likely that they are at any point from the beginning to the end of the precalculated duration of their Exposed stage ($D_{E,\,i}$) at the time of simulation start. We then select a random $N_{IM}(0)$ agents to begin the simulation with an infection status of Mildly symptomatic (IM). As with the initially Exposed agents, we set each initially Mildly symptomatic agent's time of entering the Mildly symptomatic state ($t_{IM,\,i}$) to be uniformly distributed on the interval ($-D_{IM,\,i}$, 0). For consistency, we then set $t_{IP,\,i} = t_{IM,\,i} - D_{IP,\,i}$, and $t_{E,\,i} = t_{IP,\,i} - D_{E,\,i}$; in practice, these earlier times do not affect anything in the current version of the model. In all of the results presented in this paper, we set $N_E(0) = 1$ and $N_{IM}(0) = 0$.

B. Initial infection history

We calculate the number of agents who have recovered from natural infection in the past year as the product of the total number of agents ($N$) and the fraction of agents who have recovered from natural infection in the past year ($f_{R,365}$), and randomly choose that many agents to have a finite $t_{R,\, i,}$ i.e., to have recovered in the past. (For computational convenience, events that have never happened yet are generally recorded as “having happened” infinitely far into the future.) For each of those agents, we set $t_{R,\, i}$ to be uniformly distributed on the interval (-365, 0). We do not explicitly set times for their state transitions earlier in that same infection ($t_{E,\, i}$ and one or more of $t_{IA,\, i}$, $t_{IP,\, i}$, $t_{IM,\, i}$, $t_{IS,\, i}$, and/or $t_{IC,\, i}$), as these are not necessary in the current version of the model. For each agent, we combine this (past) immunity event and any (past) vaccination immunity events (below) to calculate their immunity trajectory, their last immunity event time, and their previous immunity, in the same way that we would if they experienced those events in that order and at those intervals during the simulation.

C. Initial vaccination history

As shown in **Table S1**, for a given parameter set, we calculate the number of agents who start the simulation with each of the several different categorical vaccination histories: those who have not received a complete primary course of vaccination (who are, for the sake of simplicity, assumed to be completely unvaccinated); those who have received a primary course of vaccination, but too recently to be eligible for a booster dose; those who are eligible for a booster dose, but have not received one; those who have received a booster dose within the past $T_{V2 \to B}$ = 5 months; and those who have received a booster dose more than $T_{V2 \to B}$ ago. We then randomly assign more detailed histories of vaccination events for each agent, based on the category they are in, an assumption that all currently-boosted agents received their booster dose exactly $T_{V2 \to B}$ after completing their primary course of vaccination, and uniform distributions for the sake of simplicity (details in **Table S1**). These more detailed histories are then combined with those same agents' histories (if any) of recovery from natural infection to determine their immune characteristics at simulation start.

D. Interaction of infection and vaccination history

Infection history and vaccination history are randomly determined (as discussed above), independently of each other, and the series of immunity events that these histories define are then applied to the individual, in chronological order, with immunity calculated at each step of the process, in the same fashion as it is during simulation.

### Text S3: Disease transmission module

A. Infection transmission algorithm

Based on the current understanding of the importance of fomites in the epidemiology of COVID-19, we do not explicitly model environmental contamination or fomite transmission, although some “contacts” may, in fact, represent this sort of indirect transmission.

If both members of a pair are “available” (i.e., scheduled for that shift (either definitely or potentially, in the case of all-shift floaters) and not isolated, hospitalized, or dead) during a given shift, we assume that the actual number of effective contacts is Poisson-distributed, with a mean of the expected contact rate for that pair and that shift type. This is not meant to imply that agents enter close proximity (for example) a discrete number of times per shift, and that for any pair with a non-zero expected contact rate, this number of times may be anywhere from 0 to arbitrarily large; rather, the number of contacts is meant to represent the *effective* number of “typical” contacts (in terms of probability of transmission) between two agents. Two agents may, in this sense, make zero contacts even if they do in fact come into physical proximity, if biosafety interventions, airflow patterns, reduced shedding during a particular span of time, or any number of other factors result in a probability of transmission that is effectively zero. Conversely, two agents who are packed tightly together, with poor air circulation, for hours on end, may make an extremely high number of effective contacts, even if they only come together and then separate (hours later) a single time.

We further assume an independent action model, at this “effective contact” level, for disease transmission. Similarly, we assume that partial protection works by reducing the probability of transmission *per effective contact* by some fixed ratio (limited testing of an alternative assumption of reducing the probability of transmission *per shift* by a fixed ratio produced comparable results). Finally, we assume that the contact matrix is symmetric, i.e., that in the contact matrix $M$, $M_{ij} = M_{ji}$ for all $i$, $j$. This assumption makes sense in most contexts, but could be violated if, for example, a facility's ventilation system produces highly directional air flow that carries potentially infectious droplets from one agent to another at a much higher rate than the other way around. Thus, given an infectible worker $i$, with susceptibility to infection (relative to a fully susceptible agent) $s_i$ (= 1 - $P_{E,i}$), and an infectious coworker $j$, who has a probability $p_j$ per effective contact of infecting a fully susceptible agent, the probability that one effective contact between $i$ and $j$ results in infection of $i$ is

$$s_i p_j$$

and (by the assumption of independent action), the probability that $k$ effective contacts results in infection (assuming that $i$ is not infected by someone else first) is

$$1 - (1 - s_i p_j)^k$$

and so (by the assumption of a Poisson-distributed number of effective contacts), if the expected number of contacts during a given shift (taken into account availability, by setting all entries for which either of the agents is unavailable equal to 0) is $M_{ij}$, then the probability that $j$ infects $i$ during that shift (again, assuming that no one else does, and that $i$ and $j$ are both available (see Work module Section) is

$$\begin{aligned}\sum_{k=0}^{\infty}\left(\frac{\left(M_{ij}\right)^k e^{-M_{ij}}}{k!}\right)\left(1-\left(1-\mathrm{s_i p_j}\right)^{\mathrm{k}}\right) &= 1-\sum_{\mathrm{k}=0}^{\infty}\frac{\left(\mathrm{M_{ij}}\left(1-\mathrm{s_i p_j}\right)\right)^{\mathrm{k}}\mathrm{e}^{-\mathrm{M_{ij}}}}{\mathrm{k}!}\\ &= 1-e^{-M_{ij}s_i p_j}\sum_{k=0}^{\infty}\frac{\left(M_{ij}\left(1-s_i p_j\right)\right)^k e^{-M_{ij}\left(1-s_i p_j\right)}}{k!}\\ &= 1-e^{-M_{ij}s_i p_j}\end{aligned}$$

Hence (again, by the assumption of independent action), the probability that *anyone* infects $i$ during that shift is

$$\begin{aligned}1-\prod_{j=1}^{N}\left(1-(1-e^{-M_{ij}s_i p_j})\right) &= 1-\prod_{j=1}^{N}e^{-M_{ij}s_i p_j}\\ &= 1-e^{-\sum_{j=1}^{N}M_{ij}s_i p_j}\\ &= 1-e^{-s_i(M\vec{p})_i}\end{aligned}$$

This naturally leads to the concept of a (negative log-scale) "force of infection" equal to $(M\vec{p})_i$ operating on $i$ over the course of the shift. We take advantage of this to model infection "in the community" more simplistically, as a per-"shift" force of infection, representing the negative log of the probability of not becoming infected by an (unspecified) infected individual who is not a fellow employee (and is therefore not represented by an agent in the model). This probability can broadly be thought of as reflecting the force of (non-employment) COVID-19 infection in the broader community, which can be inferred from public health reports. This transmission route occurs only for shifts in which the agent is expected to have contact with individuals outside the modeled company, such as time spent at home (i.e., during a non-work shift), if not housed in employer-provided dormitories.

B. Modeling immune dynamics

We do not explicitly model differences in either innate or cross-immunity, except for those that result from age. We also do not explicitly model either innate or acquired immunodeficiencies.

We distinguish between 8 basic states (immunity trajectories) with respect to immunity: fully Susceptible (S), partially vaccinated (V1), Fully Vaccinated (V2), Boosted (B), Recovered (R), and Hybrid immunity (H) with partial vaccination (HV1), with full vaccination (HV2), or with full vaccination and a Booster (HB). Non-hybrid vaccinated trajectories (V1, V2, and B) feature a smooth ramp-up from their individual's previous level of immunity, that lasts for $T_{V1\rightarrow V2}$ = 21 days, $T_{ramp, V2}$ = 14 days, or $T_{ramp, B, 1}$ + $T_{ramp, B, 2}$ = 14 days, respectively, counting from the time since the individual's last immunity event (i.e., receiving their first vaccine dose, receiving their second vaccine dose, or receiving their booster shot, respectively). The non-hybrid Recovered trajectory (R) features an initial $T_{total, R}$ = two months (61 days) period of total immunity, counting from the time of their (most recent) recovery. The Hybrid immunity trajectory (HV1, HV2, and HB), as they have characteristics of both vaccinated and recovered trajectories, and can be entered either by recovery following vaccination or vaccination following recovery, have both of these features, but a particular individual's experience of them may include either or both:

- Like an individual in the Recovered state, an individual in a Hybrid state experiences total immunity for so long until it has been less than $T_{total, R}$ since the time of their most recent recovery ($t_{R, i}$).
    - For an individual who enters a Hybrid state by recovering from natural infection (having previously been vaccinated), this complete immunity will last for the full $T_{total, R}$ from the time of that entry (which will be both the time of their last recovery and the time of their last immunity event (unless and until they experience another)).
    - For individuals who enter a Hybrid state by being vaccinated (having previously recovered from natural infection), some (or even all) of that time has already been spent prior to that entry (which will be the time of their last immunity event, and greater than the time of their last recovery), and so the amount of time that they experience complete immunity after that entry will be less (or even non-existent).
- Like an individual in an exclusively Vaccinated state (V1, V2, or B), an individual in a Hybrid state will experience a ramp-up from their previous level of immunity, which lasts $T_{ramp, RH}$ = one-month (30.5 days, measured from the time of their most recent immunity event ($t_{last, i}$)) for all Hybrid states, with the caveat that this is overriden by the complete immunity that lasts for $T_{total, R}$ from the time of their most recent recovery, whenever the latter applies. Thus:
    - An individual who enters a Hybrid state by recovering from natural infection will not experience this ramp-up at all, with the normal values of $T_{total, R}$ and $T_{ramp, RH}$, nor with

any of the alternative values we tested in sensitivity testing, since these all result in $T_{\text{total, R}}$ being greater than or equal to $T_{\text{ramp, RH}}$.

- An individual who enters a Hybrid state by being vaccinated, with their last recovery having been at least $T_{\text{total, R}}$ prior to that vaccination, will experience a ramp-up for the full period of $T_{\text{ramp, RH}}$ after that vaccination.
- An individual who enters a Hybrid state by being vaccinated, with their last recovery having been less than $T_{\text{total, R}}$ prior to that vaccination, will experience first total immunity, then a shortened period of "ramp-up" (that is actually a ramp *down* from the state of total immunity). If the values of $T_{\text{total, R}}$ and $T_{\text{ramp, RH}}$ are such that $T_{\text{total, R}} < T_{\text{ramp, RH}}$ (which is not true in any of the scenarios or sensitivity tests that we have examined in this paper), then this will also be the experience of individuals who enter a hybrid state by recovering from natural infection.

Once $T_{\text{total, R}}$ has elapsed, *measured from the time of their last immunity event* (not from the time of last recovery, which may be earlier), all of these cases will produce identical waning dynamics. The transitions between these immunity trajectories are summarized in **Table S4**, and the equations for the protection that they offer are given in **Table S9**.

To parameterize curves for the V2 and B states, we used data from British vaccine effectiveness surveillance [*52*]. For simplicity of modelling, we consistently used data for Pfizer vaccine specifically, which made up 59% of vaccine doses administered in the US, as of Oct 09, 2022 [*34*]. Initially, we fitted a model with two exponentially-decaying components for both V2 and B. The fit for V2 generated two components with negligibly different decay rates, so we simplified this to a single exponential. The same was not true of B, so we left that as a two-component mixture. For Recovered and Hybrid immunity, we used logistic decay curves based on those presented in [*53*]; although that paper did not present explicit coefficients, it presented values for the protection at various times after state entry from which it was easy to calculate coefficients. Broadly in line with the approach taken in that paper, we assumed that no reinfection is possible (i.e., protection from infection is complete, $P_{E,i} = 1$) for 2 months after an individual's most recent recovery ($T_{\text{total, R}} = 61$ days), and used the logistic decay curves after that. We also assumed a one-month ramp-up to full immunity for individuals with hybrid immunity who have been vaccinated more recently than they have recovered, applicable only to time during which they do not have complete protection (if relevant).

Evidence of the extent of long-term protection from partial vaccination is limited. Because our model, in its current version, does not allow agents to remain partially vaccinated for long periods of time, we simply allowed partially vaccinated agents' protection to ramp up over the course of three weeks (i.e., until they are expected to receive their second dose, and then to remain constant (with this constant phase only being relevant if they are delayed in receiving their second dose by illness). For analogous reasons, and because [*53*] did not present results for hybrid immunity resulting from partial primary vaccination (HV1), we simply used the same coefficients for HV1 as for HV2.

The data obtained from [*53*] and [*52*] were not quite *directly* comparable, and did not present all the details our model required. [*53*] presented data for protection from any infection ($P_{E,i}$) and for overall protection from severe disease ($P_{IS,i}$), from which it is possible to mathematically calculate protection from severe disease conditional on any infection:

$$P_{IS|E,i} = 1 - \frac{1 - P_{IS,i}}{1 - P_{E,i}}$$

In order to obtain protection from symptomatic disease conditional on any infection ($P_{IP|E,i}$) and protection from severe disease conditional on symptomatic infection ($P_{IS|IP,i}$), we observed that

$$P_{IS|E,i} = 1 - \left(1 - P_{IS|IP,i}\right)\left(1 - P_{IP|E,i}\right)$$

and parameterized the relative contribution of each to $P_{IS\,|\,E,\,i}$ with a parameter φ (with a default value of 0.5, i.e., equal contributions from $P_{IP\,|\,E}$ and $P_{IS\,|\,IP}$), as follows:

$$P_{IP|E} = 1 - \left(1 - P_{IS|E}\right)^{\varphi}$$
$$P_{IS|IP} = 1 - \left(1 - P_{IS|E}\right)^{1-\varphi}$$

[*52*] presented data for overall protection from symptomatic disease ($P_{IP,\,i}$); for simplicity, we assumed that for these immunity trajectories, $P_{E,\,i}$, $P_{IP\,|\,E}$, and $P_{IS\,|\,IP}$ were all numerically equal, i.e.:

$$P_{E,i} = P_{IP|E,i} = P_{IS|IP,i} = 1 - \left(1 - P_{IP,i}\right)^{0.5}$$

The practical meaning of the changes to $\varphi$ that we examined in our sensitivity analysis, and that resulted in our finding high sensitivity of the number of symptomatic infections to this parameter, can be illustrated with an example: Consider an individual *i* who last recovered from natural infection 6 months ago, and who has never been vaccinated (meaning that their immunity trajectory is Recovered, not one of the Hybrid trajectories). With default values for $\varphi$ (=0.5) and other relevant parameters (specifically, $a_{R,\,IS}$, $b_{R,\,IS}$, $a_{R,\,E}$, and $b_{R,\,E}$ (**Table S16**)), this trajectory grants them 51.2% Protection against Any Infection ($P_{E,\,i}$), and 80.1% *overall* Protection against Severe Infection ($P_{IS,\,i}$). As noted above, this overall protection against developing severe disease ($P_{IS,\,i}$) takes into account both the 51.2% protection against any infection, and a further protection against developing severe disease, given that an individual *does* become infected ($P_{IS\,|\,E,\,i}$). Mathematically, this implies that

$$\begin{aligned} P_{IS|E,i} &= 1 - \frac{1 - P_{IS,i}}{1 - P_{E,i}} \\ &= 1 - \frac{1 - 0.801}{1 - 0.512} \\ &= 0.592 \end{aligned}$$

In our model, this 59.2% protection, against developing severe disease conditional on any infection ($P_{IS\,|\,E,\,i}$), can be further broken down into protection against developing symptomatic disease, conditional on any infection ($P_{IP\,|\,E,\,i}$); and protection against developing severe disease, conditional on developing symptomatic disease ($P_{IS\,|\,IP,\,i}$). We parametrize this breakdown by $\varphi$ as discussed above. At the default value of $\varphi = 0.5$, $P_{IP\,|\,E,\,i} = P_{IS\,|\,IP,\,i} = 36.1\%$. In sensitivity analysis, values of $\varphi = 0.25$ and 0.75 were tested. When $\varphi = 0.25$, $P_{IP\,|\,E,\,i} = 20.1\%$ and $P_{IS\,|\,IP,\,i} = 49.0\%$; conversely, when $\varphi = 0.75$, $P_{IP\,|\,E,\,i} = 49.0\%$ and $P_{IS\,|\,IP,\,i} = 20.1\%$.

Lacking good data on the amount of further protection that various forms of immunity offered from critical infection conditional on severe infection ($P_{IC\,|\,IS,\,i}$), or from death conditional on critical infection ($P_{D\,|\,IC,\,i}$), we did not include effects for these in our model. For further details on all of this, see **Tables S6-S9**.

Note: For modeling of the very early phases of the pandemic, for the purpose of model validation, we use a somewhat simpler approach: Because no one has yet been vaccinated, and we assume that no agents have already recovered from infection, and because the time duration of the outbreaks we examine is short compared to estimates of the rate of waning of immunity against even Omicron, let alone 2020 strains, we simplify matters by treating immune protection as non-waning.

C. Modeling interventions

*Temperature screening.* In our temperature screening intervention, all otherwise available agents are temperature-screened with a non-contact thermometer, using a fever-threshold of 38.0 degrees Celsius (in line with CDC recommendations), immediately before each shift that they are scheduled to work. Based on Swiss Data [*59*], we model this screening as having a specificity of 100%, but a sensitivity of only 5.2%, and even that, only for symptomatically (i.e., Mildly, since Severely and Critically infected agents are already unavailable, due to being hospitalized) infected agents.

*Virus testing.* We model virus testing as occurring with some (average) probability per agent per shift that that agent is scheduled to work, and as occurring immediately before that shift. More precisely, we assume that this probability, combined with the number of agents scheduled, defines a number of tests that are available to test agents for that shift. Agents who have been less recently tested are prioritized over those who have been more recently tested, and ties are broken randomly. When it is not possible to assign the full calculated number of tests to agents, either because this calculated number is not an integer or because sufficiently many agents are scheduled for that shift but not available (due to isolation (if combining multiple interventions) or hospitalization), then any complete or fractional “surplus” tests are added to the number of tests for the next shift. We model this testing as being able to correctly detect agents with an infection status of IA, IP, and IM (but not E), and as having a sensitivity of 90% (we rounded down the reported estimate of 91.84% in [*60*]) and a specificity of 99.95% [*60*].

*Primary Vaccination and/or boosting-promoting interventions.* We model primary vaccination and/or boosting-promoting interventions as resulting in a specified fraction of agents, who are eligible for the indicated type of vaccination (i.e., for the first shot of their primary series and/or their booster dose, as the case may be) and who would not otherwise receive it, receiving it each day, during a shift in which they are awake and not working. We assume that all agents who receive a first dose of their primary series as a result of such an intervention go on to receive their second dose 21 days later.

*Direct $R_0$-reduction (biosafety and/or physical distancing) interventions.* We model interventions that work by (relatively) directly reducing $R_0$ (such as most physical distancing and/or biosafety interventions) as doing precisely that, resulting in a lower average contact rate being set by the code that sets that contact rate in order to achieve a certain (homogeneous) $R_0$. We do not differentiate such interventions from each other in our ABM model (as opposed to subsequent economic analysis), except by how large of a proportional reduction they produce.

*Isolation and Deisolation.* Under those interventions that involve testing agents to determine whether they are infected (temperature screening and virus testing), the conditions under which an agent can be unavailable for a scheduled work shift (section “Work module”) include not only hospitalization or death, but also isolation. An agent enters a state of isolation whenever they test or screen positive; the question of when they should *de*isolate is somewhat more complex.

The CDC has recommended the use of the following set of criteria [*61*] for discontinuing isolation following a positive temperature (and other symptomatic) screening result (or self-identified COVID-19 symptoms, although we do not include workers self-isolating on their own initiative in response to symptoms in the current version of the model) and/or following a positive virus test:

- For symptomatics, "fever [has] end[ed] for 24 hours (without the use of fever-reducing medication) and other symptoms are improving,"
- For everyone, a certain number of days have passed:
  - 5 days, with masking when around other people for 5 days more:
    - For those who never develop symptoms, 5 days since their first positive test;
    - For those who develop mild symptoms, 5 days since symptom onset (even if they had a positive test prior to that point)
  - 10 days from symptom onset, for "[p]eople who have moderate COVID-19 illness,"
  - "at least 10 days and up to 20 days after symptom onset [...] may be warranted," for "[p]eople who are severely ill (i.e., requiring hospitalization, intensive care, or ventilation support)."
  - "20 or more days" for "[p]eople who are moderately or severely immunocompromised"

A number of these points are irrelevant and/or difficult to implement in our model:

- We do not explicitly model improvement of symptoms, as distinct from recovery from the infection that produced those symptoms. Therefore, we do not implement the requirements that it be at least 24 hours since fever (if any) has resolved and that symptoms must be improving; instead, we simply require that anyone who has been isolated and has had symptoms must be recovered before being deisolated and returning to work.
- We do not currently implement masking for 5 days after deisolation for asymptomatic or mildly symptomatic cases, because of difficulties in implementing this in an internally consistent fashion across both scenarios with and without masking of the workforce as a whole. Moreover, given the above point (and our handling of reinfection following recovery), this would only be of practical significance with respect to asymptomatic cases, as mildly symptomatic cases are required to no longer be infected (and hence, to longer be infectious) when they return to work, and are not at risk of being infected themselves so soon after their previous infection.
- We do not explicitly distinguish between mild and moderate cases as this distinction may not always be clear to employers and employees; consequently, we cannot selectively implement 10-day isolation of moderately symptomatic cases. In any event, as noted above, we require symptomatically infected agents who are isolated to be recovered from infection before returning to work. In addition to this rendering the presence at work between days 5 and 10 of anyone whom we allow to return to work irrelevant in terms of transmission dynamics, it also means that the vast majority (approximately 95%) of "mildly" (or moderately) symptomatically infected agents will in fact be isolated for longer than 5 days, and a substantial fraction (approximately 16%) for longer than 10 days, given our use of a $Gamma(shape = 16, scale = 0.5)$ distribution for length of mildly symptomatic infection.
- We do not require "up to 20 days" to have elapsed since symptom onset for deisolation of severely or critically symptomatic cases. However, this is of little practical significance, given our requirement of recovery from infection, as most (approximately 3/4) of severe and effectively all (>99.9%) of critical cases will have an interval > 20 days between symptom onset and recovery (if any), given the distributions we use for the length of mild, severe, and critical symptoms. We do not explicitly simulate, and hence cannot explicitly account for a longer period of isolation of immunocompromised individuals.

Thus, our actual deisolation criteria are: (i) If the agent has had symptoms (of whatever severity), they are recovered from infection, and (ii) At least 5 days have elapsed since whichever is more recent of symptom onset or a positive viral test or temperature screening.

### Text S4: Work environment module

#### A. Farm model

We model a farm as consisting of one manager, under whom there are some number $n_s$ of supervisors. Under each supervisor, there is some number $n_{c,s}$ of crews, each of which consists of one foreman and some number $n_{w,c}$ of workers. Thus, the total workforce consists of one manager, $n_s$ supervisors, $n_s * n_{c,s}$ foremen, and $n_s * n_{c,s} * n_{w,c}$ workers, for a total of $1 + n_s * (1 + n_{c,s} * (1 + n_{w,c}))$ agents.

We refer to a supervisor and the crews under their supervision collectively as a team. All agents are assumed to be present for the same hours, namely the first shift of each of the first five days of each week. Consequently, all entries in the contact matrix $M$ for those shifts are non-zero, but their magnitude varies appreciably. The expected number of contacts for a pair of agents depends both on the role of each agent (manager, supervisor, foreman, or worker) and on the proximity of the two agents in the hierarchy (same crew (only possible if both agents are workers, or if one is a worker and the other is a foreman), same team (only possible if both agents are foremen or workers, or if one is a foreman or worker and the other is a supervisor), or "other," (meaning the two are on different teams, or one of them is manager)). The *relative* contact rates for each combination of these factors are given in **Table S11**. The absolute contact rates for work shifts are then calculated as described in section “Absolute contact rates” (**Text S4D**).

As previously noted, the second shift of each work day, and the first and second shifts of each non-working day, are considered to be "awake, non-work" shifts. Transmissions that are not attributable to work, namely transmissions "from the community" if housing is individual, or transmissions within shared, company-provided housing if housing is shared, occur during these shifts.

B. Facility model

In contrast to the relatively simple schedule of shift types in the farm model, the handling of shifts and worker presence in the facility model is somewhat more complex. In the facility model, for each work day, the first shift is designated as Production Shift 1, the second shift is designated as Production Shift 2, and the third shift is designated as the Cleaning Shift. The user can designate a facility as having either 1 or 2 production shifts ($n_{sh}$) per day; for a facility with only 1 production shift per day, workers will be present during Production Shift 1, and not during Production Shift 2.

For each Production Shift that has workers present, our model includes 1 shift supervisor, under whom there are some number $n_l$ of production lines, each of which has $n_{w,l}$ workers working on it, for a total of $n_l$ * $n_{w,l}$ production line workers. As in the farm model, workers who are closer in the hierarchy (i.e., workers who are on the same production line, as opposed to those who are on different production lines) have a higher contact rate than those who are more distant. In addition, unlike in the farm model, we incorporate within each production shift $n_{f,sh}$ within-shift "floating workers" who are assumed to have a more diffuse contact pattern; these may represent a variety of specialized workers who are not assigned to a single production line for the duration of their shift. For the cleaning shift, we have a simpler approach: there is homogeneous mixing within the pool of cleaning shift workers (of which there are $n_{cs}$).

Finally, the model also includes a number $n_{f,all}$ of agents, including but not limited to the overall manager, who are *between*-shift "floating workers," meaning that their contact patterns are not only diffuse within a shift, but are spread across all staffed shifts (i.e., across both Production Shift 1 and the Cleaning Shift, or across all three shifts, depending on whether the facility has one or 2 productions shifts per day). This is not intended to indicate that they are present 24 hours a day, five days a week, but rather, that their schedule may not be consistent from day to day. Accordingly, they are treated as being "one-half present" or "one-third present" on each staffed shift, for the purpose of calculations for which this is practical (transmission and vaccination assignments). For testing, this is not practical; consequently, each between-shift floating workers is independently randomly assigned a single shift to which they will be treated as being assigned for testing purposes only. These simplifications do not affect model predictions. Further details of the calculation of contact rates are provided in section “Absolute contact rates” (**Text S4D**).

As noted and illustrated in **Figure 1B.i**, for a worker who is assigned to work a specific shift (first, second, or third), the shift after that (i.e., second, third, or first, respectively) of each work day, and both that shift and the shift that they otherwise work of each non-working day (e.g., the first and second shift of a non-working day, for someone who works the first shift of each work day) are considered to be "awake, non-work" shifts. Transmissions that are not attributable to work, namely transmissions "from the community" if housing is individual, or transmissions within shared, company-provided housing if housing is shared, occur during these shifts.

Between-shift "floating" workers' awake non-work shifts are distributed according to the same basic pattern, taking into account their "one-half" or "one-third" presence on each work shift.

Work and sleep schedules are illustrated in **Table S10**.

C. Assumptions common to the farm and facility models

Briefly, we assume the following:

- All workers live in the same type of housing (i.e., either individual or employer-provided shared housing).
- All workers work a regular 40-hour, 5-day work week (8-hour shifts) and 2-day weekend, with the model simulation starting on a random day of the week, except for a small number of floating workers (e.g., quality assurance technician, mechanic) in the facility model, for whom the work shift(s) on a given day is/are randomly selected.
- There are many more contacts within the hierarchical structure than outside it (e.g., more contacts between workers on the same crew/production line than between workers on different crews/production lines, more contacts between foremen and supervisors than between other workers and supervisors, etc.), but that contacts are possible between any two agents who are both present on the same shift.
- All contact between workers occurs either (a) while traveling to, at, or traveling home from work, or (b) in shared, employer-provided housing, i.e., that workers in individual housing do not socialize with each other outside of work.
- There is homogeneous mixing within employer-provided housing.

Worker contacts on the way to and from work follow the same basic patterns as worker contacts at work (e.g., we tacitly assume that shared transportation is substantially more likely to group together workers who are on the same crew than workers who are on different crews).

D. Absolute contact rates

We calculate relative contact rates from user set parameters as summarized in **Tables S11 and S12**. From this, we calculate the average total at-work contact rate for all workers on a work day, and then scale this by 5/7 to get an average total (unscaled) at-work contact rate all workers for all days of the week. We then multiply this unscaled “contact rate” by the sum, over all three potentially coworker-infecting infectious stages (IA, IP, and IM), of the product of the average duration per infection of that stage (taking into account the probability of not entering it at all, given complete susceptibility, no interventions, and the age distribution used to randomly set employee ages) and the per-contact transmissibility for that stage. This gives us an unscaled at-work $R_0$ under homogeneous mixing; we then multiply all unscaled contact rates by the ratio of the user-set desired at-work $R_0$ to this calculated unscaled $R_0$. (The assumption of homogeneous mixing will arguably distort the calculated value slightly for the farm model, but to a very limited extent compared to some of our other assumptions, and precisely defining what we mean by $R_0$ for an epidemic on a finite network of relatively modest size is tricky anyway. For the facility model, all employees have the same total contact rate per day, and so this is even less of an issue.)

Likewise, for scenarios with shared housing, we calculate an average daily at-home (unscaled) contact rate with other employees, taking account of the fact that each employee (straightforwardly, for employees that are not all-shift floaters; on average for any that are (applicable to the facility model only)) has 9 "at home, awake" shifts per 7 days. We then use and scale this calculated value in a precisely analogous fashion to how we use and scale the corresponding at-work value.

**Text S5: Interventions cost**

A. Common assumptions applicable to multiple types of interventions

*Time compensation:* We assume that time that employees must dedicate to intervention-specific tasks (training, performing and receiving temperature screenings, performing viral self-tests and waiting for results, and receiving primary vaccinations and/or boosters in response to an employer-initiated program to increase primary vaccination and/or boosting rates) occurs outside of normal working hours (whether greatly so or immediately prior to work). We assume that this time is compensated at employees' regular wages.

*Mask and face shield cost and durability:* We also assume the following costs and durabilities of materials: A KN-95 mask (or near equivalent) costs $1 and may be used by one worker, for one shift. A face shield costs $3 and may be used by one worker, for 30 shifts.

B. Temperature screening

Employers who conduct routine temperature screenings may choose to hire a third-party contractor to perform those screenings, use on-site medical staff, or provide non-specialist employees with the necessary training. In this analysis, we assume that employers use the last of these options. Temperature screening cost therefore includes (1) the initial setup cost including training and equipment, (2) ongoing compensation for the employees conducting screening, and (3) personal protection equipment (PPE) for these employees. Setup cost is a one-time cost that depends on the number of screeners conducting the screening, hourly wage, and unit cost of the equipment. We simplify matters by assuming only minimal essential equipment: one noncontact thermometer per screener, one KN95 per screener per shift, and one face shield per screener per 30 shifts. We assume that one trained screener is required per 100 employees (or fraction thereof), and that training takes 1 hour. The time required for screening before each shift (and hence, the amount of compensation) is calculated based on the number of employees available for that shift, under the assumption that a single measurement takes 3 seconds on average.

C. Virus testing

Virus testing is assumed to be conducted immediately prior to a scheduled work shift, with commercially available rapid testing kits. The mechanics of virus testing are discussed in **Text S3C**. We assume that the cost of one kit is $10 (which is in line with typical costs). In our analysis, we assume employers pay for 100% of the cost. We assume that the waiting time for using the kit is 15 minutes, for which the employee is compensated with their regular hourly wage (as provided by the user).

D. Vaccination and boosting

Current FDA-authorized COVID-19 vaccines and booster shots are distributed for free by states and local communities. Therefore, the vaccination and booster doses themselves do not incur direct costs to the employers. In our analysis, we assume that employees are paid for 0.75 hours (45 minutes) of time taken outside of working hours to receive each dose.

E. Biosafety intervention

The intensities of biosafety strategies are defined in terms of what those interventions achieve, as measured in the reduction in the $R_0$. We use specific examples of "low", "medium", and "high" intensity interventions that are chosen, in part, to illustrate how higher-intensity interventions may be built by "stacking" multiple lower-intensity strategies, and how this can affect net cost.

- For low-intensity intervention, we use KN95 masks, one per employee per shift.
- For medium-intensity intervention, we add face shields, one per employee per 30 shifts, on top of the low-intensity intervention.
- For a high-intensity intervention, we combine this medium-intensity intervention with the use of (a) portable air cleaner(s). CDC and the American Society of Heating, Refrigerating and Air-Conditioning Engineers (ASHRA) recommend using a filter with a Minimum Efficiency Reporting Value (MERV) of 13. In most cases, it should not require new building ventilation systems but simply upgrading filters. The ventilation systems and energy costs differ for each facility. In this analysis, we estimate the cost of using (a) portable high-efficiency particulate air (HEPA) fan/filtration system(s) as an alternative or auxiliary tool for improving ventilation. Choosing (a) suitable portable unit(s) depends on the area in which it will be installed. Some commercial HEPA air purifiers are listed below to provide a general picture of the cost and area (sq. ft. of floorspace) they cover. We assume portable air cleaners have an average cost of $1,000 per 1,000 sq. ft. and a life span of 3 years (1,095 days).
    - EJ120 Air Purifier cleans up to 1,250 sq ft. - $899.00 (Medical Grade True HEPA filter (MERV17) [*62*]
    - Airpura P600 cleans up to 2000 sq ft. - $1,199.98 [*63*]
    - Enviroklenz UV-C Air Purifier cleans up to 1,000 sq. ft. - $799.00 [*64*]
    - aair Medical Pro cleans up to 1,080 $ft^2$ - $1,299.00 [*65*]
    - Blade Air HEPA-Carbon Air Purifier cleans up to 1,400 $ft^2$ – 1,363.00 [*66*]

**Text S6: Model validation**

In this document, the identities of the produce farms and food processing facilities where outbreaks used for model validation had occurred are concealed to protect the companies' and employees' privacy.

A. Farm operation model

For produce farm operations, FInd CoV Control was validated using two outbreaks on produce farms.

- In the first outbreak on a fruit orchard farm, with shared housing, detection began with an initial 6/77 individuals testing positive (with 2 ultimately requiring hospitalization), then 45 more testing positive over a 2-week period (total 51 employees positive). In 401 model simulations, an initial 2 employees requiring hospitalization simultaneously occurred in 71% of simulations and resulted in a median 34 total illnesses (range 2-70; interquartile range 21-47.5) over the next 14 days.
- In the second outbreak on a strawberry greenhouse farm, with a mix of shared and individual housing, detection began with 2 symptomatic workers out of 340. Subsequent testing over the following week resulted in detection of approximately 170 total cases. In 101 model simulations, an

initial 2 employees testing positive simultaneously occurred in 100% of simulations and resulted in a median 190 total illnesses (range 46-290; interquartile range 157-219) over the week.

B. Processing facility model

For processing facilities, FInd CoV Control was validated using three outbreaks, one in each of dairy, pork and produce processing facilities.

- In the first outbreak, in a dairy operation with individual housing, available reports indicated 80 people (out of 275-300 workers) detected as infected within 1 week, ending April 26, 2020, when the outbreak was nearly over. For this facility, the process of validation involved evaluation of the plausibility of the outbreak timing (i.e., when should the outbreak have started based on its size during the week ending April 26, 2020). In 1001 model simulations, the median result has the outbreak starting (with 1 exposed individual) on March 23, 2020. The interquartile range is 'never' to March 30, 2020 (where 'never' means that over 25% of runs either failed to generate any single week in which 80 infections could plausibly be detected through mass testing, or took over 6 months to do so, which would put the start of the outbreak at or before late October 2019, well before the presumed date of COVID-19 introduction into the USA in late January or early February of 2020 that is generally accepted by the scientific community).
- The second outbreak was in a pork processing facility with individual housing and 2,400 workers. In this facility, workers sometimes self-tested and then isolated on symptom onset. This outbreak was monitored based on cumulative self-isolations, with the dates recorded at 5, 20, 100, and 200 cumulative self-isolated workers (including any who had completed self-isolation and returned to work). Validation was achieved with 15% self-isolation (and adjustment of $R_0$ =3.2 based on a mathematical modeling study [*67*] where this outbreak has been documented as "Pork Plant B"). In 201 model simulations, the median interval between the days with 5 and 100 cumulative self-isolations was predicted at 9 days, interquartile range 8.7 to 9.7 days, $2.5^{th}$-$97.5^{th}$ percentiles of 7.3 to 10.7 days (compared with the interval 2 to 10 days inferred from [*67*]). The median time between the day with 100 and 200 cumulative self-isolations was predicted at 4.7 days, interquartile range 4 to 5.3 days, $2.5^{th}$-$97.5^{th}$ percentiles of 3 to 6 days (compared with the interval 5 to 14 days inferred from [*67*]). The predictions thus had reasonable overlap with the ranges used for model fitting in [*67*] that were in turn based on the data reported in the same study.
- The third outbreak was in a produce processing facility with individual housing and 829 workers. In this facility, the first case in the facility was detected on May 1, 2020. Then, in testing of all employees on June 13, 2020 (i.e., 43 days after May 1), 236 employees were found positive. In 1001 model simulations, on day 44 (i.e., 43 days after May 1), the model predicted a median of 206 infected workers, interquartile range 178-234, 2.5th - 97.5th percentiles 129-283, consistent with the observed 236 positive cases.

### Text S7: Insights about farm, large facility and stochastic die-off

Results for the farm model are qualitatively comparable to those for a facility with similar parameters (shown in **Figures 2-4)**. Outcome estimates for a larger facility with 1,003 employees (**Figs. S2, S3** and **S4**) are generally similar, with the main differences being later-peaking outbreaks (due to greater incidence growth required to saturate the larger population) and an associated modest increase in the effectiveness of some interventions. There is also a modest reduction in noisiness, which leads to a reduction in the probability of experiencing labor shortages in individual runs. Differences associated with shared versus individual housing are covered in the "Scenario analysis" section (and **Text S9**).

In the absence of repeated reintroduction from the broader community, the effect of early stochastic die-off is illustrated in **Figure 2C**, visible as two modal regions (groups) with respect to the predicted outbreak size: (i) small or non-existent outbreaks and (ii) large outbreaks (**Figure 2D**). At baseline, over the 90 days since the introduction of an index case, group (i) has 0-12 (median 0, mean 0.60) total infections, not including the index case, and 0-4 symptomatic infections (median 0, mean 0.16), with no infected individuals left by the end of simulation, while group (ii) has 51-97 total infections (median 82, mean 81.9), not including the index case, and 17-49 (median 34, mean 33.1) symptomatic infections. There are also 5 runs (out of 1000) that fell between these two groups, with 21-42 total infections (median 39, mean 35.8) and 5-14 symptomatic infections (median 11, mean 10.6).

### Text S8: Additional results from counterfactual comparison of interventions

Model predictions are characterized by both a large degree of variance in outcomes within an intervention (or within the baseline), and a substantial variance in the counterfactual effects of an intervention. Even an intervention with very poor mean and median performance, such as temperature screening (median reduction in the number of symptomatic infections = 1; mean = 3.1), can be extremely effective in individual runs (maximum reduction of 41, close to the maximum across all interventions of 48). Similarly, even an intervention with a moderate mean and median performance, such as viral testing at a moderate intensity (30% of workers scheduled for each shift) (median reduction in number of symptomatic infections of 29; mean of 27.0), can prove ineffective in individual runs (31 runs with no reduction in number of symptomatic infections, and 1 with an increase of 1).

As noted in the main text, some interventions that are modestly beneficial on average, such as "the moderate effectiveness (-40% $R_0$) biosafety/physical distancing" intervention can even be moderately counterfactually counterproductive in individual runs (68 of 1,000 model runs predicted an *increase* in number of symptomatic infections of 1-20). This may seem counterintuitive, given the care we have taken to make these counterfactual comparisons as direct as possible, and the fact that none of the interventions we consider have *direct* negative effects in our model. This is, in essence, explained by chance, that a moderately bad event may sometimes have consequences that block a worse event from happening. So, for example, a particular employee might, at baseline, become infected on Day 5, develop an asymptomatic infection, recover without transmitting to anyone else, and be completely immune to further infections for the following 61 days. A biosafety intervention might cause them *not* to be infected on Day 5, and as a result to be infected on Day 15 instead (by a contact that would have occurred regardless, but that could not have infected them if they were already infected or recently recovered), and to thereafter (whether due to an extra 10 days of immune waning, or simply to worse luck on Day 20 than they would have had on Day 10) develop a *symptomatic* infection, and to transmit to numerous other employees in the following days. In this case, the biosafety intervention will have delayed (but not, in the end, prevented) that one employee's infection, and indirectly caused multiple additional infections, even though its direct effect (preventing that one employee's infection on Day 5) was strictly positive.

We note that the large outbreaks infect a fraction of the population that is only weakly dependent on the exact value of $R_{eff}$. Consequently, a difference can be seen between (a) some interventions (principally vaccination and boosting) whose distribution of outcomes appears to differ from baseline primarily through a modest quantitative reduction in the location of this higher modal region (**Figure**

**2F**), with little change in the fraction of probability mass that is in each of the modal regions, and (b) other interventions (most clearly, the highly effective (-80% $R_0$) physical distancing/biosafety intervention and the moderately and highly effective virus testing interventions) that appear to have a pronounced increase in the fraction of runs resulting in few or no symptomatic infections (**Figure 2F**).

#### Text S9: Scenario analysis

The factors in scenario analysis are shown in **Table S15**. We observed in our primary scenario analysis that symptomatic infections and unavailability were both higher in our "Individual" housing setting (where there is community transmission, but not dormitory transmission) than in our "Shared" housing setting (where there is dormitory transmission, but not community transmission). The simplest explanation for this would be that both dormitory transmission and community transmission increase symptomatic infections and unavailability, but that the effects of community transmission are (at the parameter values we examined) stronger. To confirm that this was the case, we created a version of the model in which the presence or absence of community transmission and the presence or absence of dormitory transmission could be set independently, and then repeated the same analyses that generated **Figure 5**.

The results of these modified analyses are shown in **Fig. S9**. As expected, we see for all outcomes, wherever either dormitory or community transmission is present as a node at all, its presence is associated with a higher value of the outcome in question than its absence. For both number of symptomatic infections and number of worker shifts unavailable, wherever dormitory transmission is present as a node, it is lower in the tree (less significant) than community transmission. Both of these results accord with the simple explanation discussed above. As in the primary analysis, only intervention parameters appear in the tree for intervention expenses (**Fig. S9C**). The one slightly surprising result, given the generally stronger effects of community transmission compared to dormitory transmission, is that this pattern is reversed when it comes to production losses (**Fig. S9D**). This results from the fact that production losses only occur when many employees are sick *at the same time*. Therefore, even though community transmission (which provides additional opportunities for a large outbreak to occur, but has little effect on outbreak size given that a large outbreak occurs) has a stronger effect on the total number of worker shifts missed, dormitory transmission (which has a modest effect on the probability of a large outbreak occurring, but a somewhat stronger one on the size of a large outbreak, given that one occurs) has a stronger effect on the propensity of numerous of those missed shifts to occur simultaneously.

To illustrate the sensitivity of intervention effects to the degree of community transmission, we present, in **Figs. S10**, **S11**, and **S12**, results analogous to those in **Figures 2**, **3**, and **4** (respectively), for a scenario identical to the one that generated **Figures 2**, **3**, and **4**, except that community transmission is "Intermediate," instead of "None." Unsurprisingly, results are worse overall. What is more interesting is the pattern of relative effectiveness among interventions – when confronted with repeated reintroductions from the broader community, the effectiveness of testing interventions suffers particularly badly. There are two causes for this: First, testing interventions' effects are especially attributable, even compared to other interventions, to preventing large outbreaks from occurring at all, and so are particularly heavily impacted by having to deal with multiple introductions instead of just one, increasing the probability that *some* introduction will still manage to generate a large outbreak. Second vaccination and boosting interventions, as we have modeled them, are slower-acting than other interventions, including testing interventions. This means that they will have a greater impact on outbreaks that occur (generally from reintroductions) late in the simulation than those that occur early (generally due to the one agent who starts the simulation in the Exposed state.

**Text S10: Sensitivity analysis**

A. Approach.

Parameter values tested in sensitivity analysis are shown in **Table S16**. Plots showing sensitivities for all parameters in **Table S16** are shown in **Figs. S5 to S8**.

As summarized in **Table S2** in the main text, we have several pairs of sensitivity parameters that define Gamma distributions for various infection stage durations, using the notation Gamma(shape, scale). As can be seen in **Table S16**, however, we did not use the shape and scale parameters directly as our sensitivity parameters, as this would mean that increasing *either* of these parameters, while holding all other parameters fixed, would increase the mean of the distribution in question, thereby potentially unnecessarily causing effects that are due solely to changes in this mean to appear as sensitivities to two separate parameters. Instead, we use the mean ($\mu_{\text{stage}}$) and the shape ($k_{\text{stage}}$) of the distribution as our two sensitivity parameters; hence, when we increase or decrease the shape, while holding all other parameters fixed, we correspondingly increase or decrease (respectively) the scale, in order to keep the mean fixed.

For example, at default values of our sensitivity parameters, the Duration of Presymptomatic Infection, for a given agent *i* ($D_{\text{IP, i}}$), is randomly set, following a Gamma(1.058, 2.174) distribution (**Table S2**). Mechanically, this is actually a Gamma($k_{\text{IP}}$, $\frac{\mu_{IP}}{k_{IP}}$) distribution (where $k_{\text{IP}}$ = 1.058 and $\mu_{\text{IP}}$ = 1.058 * 2.174 ≈ 2.300 (**Table S16**). Thus, when we reduce $k_{\text{IP}}$ to 0.5 times its default value for the purpose of sensitivity testing, $D_{\text{IP, i}}$ now follows a Gamma(0.529, 4.348) distribution.

B. Sensitivity of the symptomatic infections outcome.

For symptomatic infections (**Fig. S5**, first column of **Figure 6**), we see the largest effect for two parameters of mildly symptomatic infection (the mean duration ($\mu_{\text{IM}}$) and relative per-contact probability of transmission ($\beta_{\text{IM}}$) for this stage), followed by a parameter ($\varphi$) that pertains to how much protection from developing symptomatic disease Recovered and Hybrid immunity provide to infected individuals. The direction of all of these effects is negative: when the parameter is increased, the number of symptomatic infections decreases. The negative effect of $\varphi$ on the number of symptomatic infections is straightforward: increasing protection of (some) infected individuals from developing symptomatic infections will, all else being equal, decrease the number of symptomatic infections. This effect will be further strengthened, in most cases, by decreased secondary transmissions. The negative effect of $\mu_{\text{IM}}$ and $\beta_{\text{IM}}$ is less direct, and comes from two sources: (i) We calculate effective contact rates in order to obtain a certain (homogeneous) $R_0$. Thus, increasing either the duration or the relative per-contact transmissibility of mild infection will, all else being equal, increase the fraction of $R_0$ that comes from transmissions during mild infection, and decrease the corresponding fractions for asymptomatic and presymptomatic infection (while maintaining the same ratio between the latter two). This, in turn, will increase the fraction of transmissions, in the absence of any immunity, that come from individuals on the symptomatic path. Because our default settings include immunity that reduces the probability of an infected individual taking the symptomatic path, this change will result in that immunity producing a larger reduction in $R_{eff}$ than it would otherwise. This is therefore in some respects a modeling artifact, as it derives from how we convert some of our sensitivity parameters into derived parameters that are used directly in our simulations. But it may also have some practical significance, in terms of the inherent sensitivity of this or other models or analyses to assumptions or inferences about the distribution of transmission potential between different types and stages of infections, when $R_0$ is estimated using methods that do not give information about that distribution. (ii) Increasing the fraction of $R_0$ that is attributable to transmissions from mild infection also increases both the fraction of transmissions that are from *currently* symptomatic individuals and the fraction that is attributable to transmissions that occur late in infections. The effects described in (i) and (ii) mean that the increased fraction of transmissions from currently symptomatic individuals (at a given $R_0$) should increase the effectiveness of any testing intervention that depends on the presence of symptoms and the increased fraction from late in infection should increase the effectiveness of just about any testing intervention.

C. Sensitivity of economics outcomes and $R_{eff}$

For worker-shifts missed (**Fig. S6**, second column of **Figure 6**), we see the strongest effects from relative frequency of severe infection, given any symptomatic infection ($\psi$), followed by the mean duration of severe infection ($\mu_{IS}$). These both have pronounced effects on the number of worker-shifts missed, but not on other outcomes, as we would expect (given our assumption that severely-infected individuals must be hospitalized, and therefore cannot contribute to further workplace transmission). Effects of all of these parameters on $R_{eff}$ (**Fig. S8,** third column of **Figure 6**) and total cost (**Fig. S7**, forth column of **Figure 6**) were generally smaller than effects on the other two outcomes. The effects on $R_{eff}$ are largely mediated by a greater fraction of transmission potential (as measured by contributions to a constant $R_0$) coming from symptomatic infection, resulting in a lower $R_{eff.}$ in the presence of (partial) immunity (as noted above), and by factors that increase the average level of immunity at the start of the simulation. $R_{eff}$ is also affected by a higher mean duration ($\mu_{IM}$) or relative infectiousness ($\beta_{IM}$) of mildly symptomatic infection, when temperature screening or viral testing makes an infected individual more likely to become isolated during their mildly symptomatic infection. Effects on worker-shifts missed (second column of **Figure 6**) exhibit a striking pattern of interactions between sensitivity variables and viral testing: For $\mu_{IM}$ and $\beta_{IM}$, the *greatest* sensitivity to changes is seen in the presence of viral testing; for $\psi$ and $\mu_{IS}$, the *lowest* sensitivity is seen in the presence of viral testing. The explanation for this is that changes to the value of the latter two variables largely affect the number worker-shifts missed due to hospitalization, and such changes therefore have a smaller effect when unavailability is more heavily driven by isolation. For $\varphi$, sensitivities are more similar across interventions. The only notable exception is a modestly higher effect under temperature screening, which only detects and isolates symptomatics.

**Text S11: Derivations of defaults for initial fractions**

To derive the default for the initial fraction of agents who have recovered from natural infection within the past year, we multiplied the number of observed COVID-19 cases in the US between February 15, 2021 and February 15, 2022 [*34*] by the fraction of COVID-19 cases observed in 18 to 64 year olds [*35*] and an estimated 4:1 ratio of total infections to observed cases [*35*], and divided by the number of 18 to 64 year olds in the US [*32*]:

$$f_R = \frac{50{,}229{,}225 * 0.69984 * 4}{0.616 * 331{,}449{,}281} \approx 0.69$$

To derive the defaults for the initial fractions of agents who have received a full two-dose primary series, the initial fraction of agents who have received a full two-dose primary series within the past five months, and the initial fraction of agents eligible for a booster dose who have received one, and the initial fraction of agents eligible for a booster dose who have received one within the past five months, we performed analogous calculation, but using vaccination data [*33*] instead of case data:

$$f_{V2} = \frac{145369049}{204172757} \approx 0.71$$

$$f_{V2,recent} = \frac{17615044}{204172757} \approx 0.09$$

$$f_B = \frac{59119317}{127754005} \approx 0.46$$

$$f_{B,recent} = \frac{57964964}{127754005} \approx 0.45$$

**Text S12: Derivation of coefficients for logistic decay formulas**

Although the coefficients we use are not explicitly presented by Bobrovitz *et al.* [*53*], that paper does present the level of protection after various intervals, and so we performed logistic regressions, and confirmed that the predicted values matched all of those given to within rounding error. As they gave intervals in months, and our calculations are performed in terms of days, we divided the time-dependent coefficients generated by the regressions by 30.5 in order to obtain the coefficients listed in **Table S16**.

**Text S13: Model running**

For each run, the model is first initialized. Then, at each time step, the following processes occur:

- Agents eligible for deisolation (**Text S3C**) are deisolated.
- If any testing is being performed (**Text S3C**), agents who are scheduled to work and (potentially) available are tested.
  - If the testing probability per shift is 1, then all (potentially) available agents are tested.
  - If the testing probability per shift is < 1, then the number of tests to be performed is determined, and these are performed on the (potentially) available agents in order from least to most recently tested, randomizing ties.
  - If any agents test positive, they are isolated, and their isolation time is set to the present time.
- Agents to be vaccinated are randomly selected (meaning that they are not infected and either they have just become eligible for a booster and are boosting on time, or they are eligible to receive some form of vaccine, and there is a vaccination-promoting intervention), and their immunity event times, immunity trajectories, and vaccination statuses are updated accordingly.
- Transmission (potentially) occurs, with probabilities as described in **Text S3A**.
- Infected agents who are eligible to leave their current state of infection (i.e., the sum of their time of entering that state and their precalculated duration for that state is less than the time at which the current time step ends) do so, randomly selecting which new infection state to enter, if necessary, as described above.
  - This step is repeated as necessary, i.e., an agent may in principle progress twice in a single time step if the duration of one of their infection states is sufficiently small.
  - If an agent recovers, their last recovery and last immunity event times ($t_{R,i}$ and $t_{last,i}$) are set to the present time, and their immunity trajectory is updated.

Outcomes are recorded for use in subsequent analysis.

### Supplementary figures

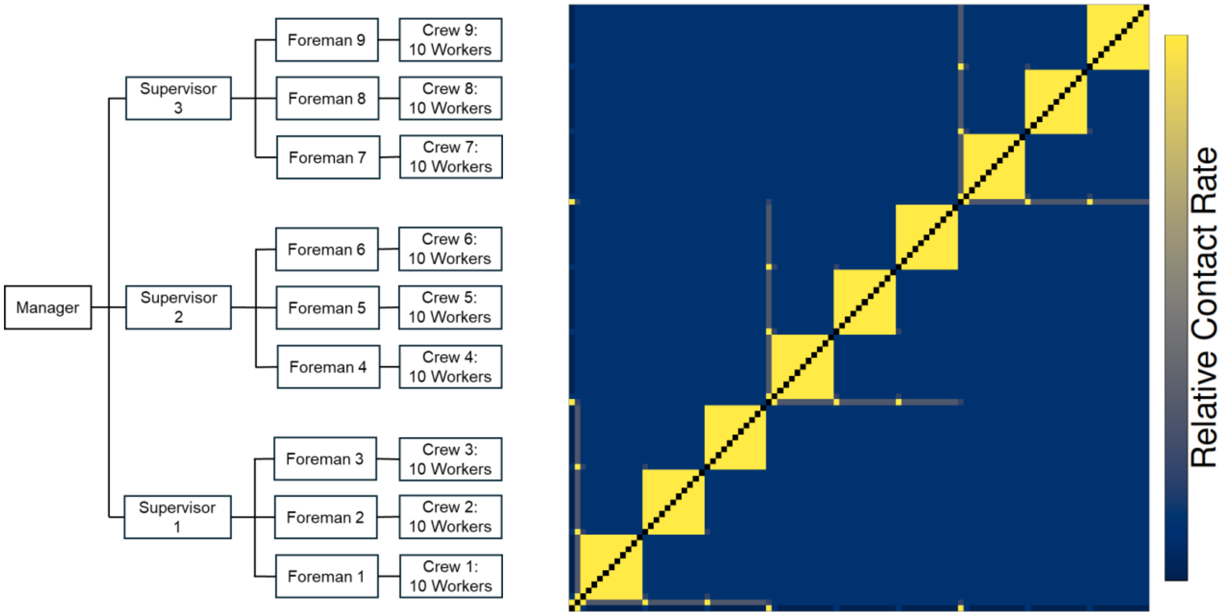

**Fig. S1. Schematic representation of the agent number and hierarchy (left) and agent contact network and relative contact rates (right) in the work environment module for a produce farm.** The relative contact rates are calculated from user set parameters in **Tables 2** and **S11**. These relative contact rates are multiplied by a constant factor (i.e., scaled) to obtain the user set $R_0$(s) when calculating the effective contact rates. Because of scaling, the values of relative contact rates for individual employee pairs in the contact matrix should only be interpreted relative to each other. For illustration, the contact network shown here is for a farm model with individual housing and otherwise default parameters; the relative contact rates between pairs of farm employees range from 0.01 (for an "ordinary" employee (not a foreman or supervisor) and the on-site manager of the operation) to 1 (for a pair of employees who are members of the same crew and who are therefore expected to make frequent contacts at work).

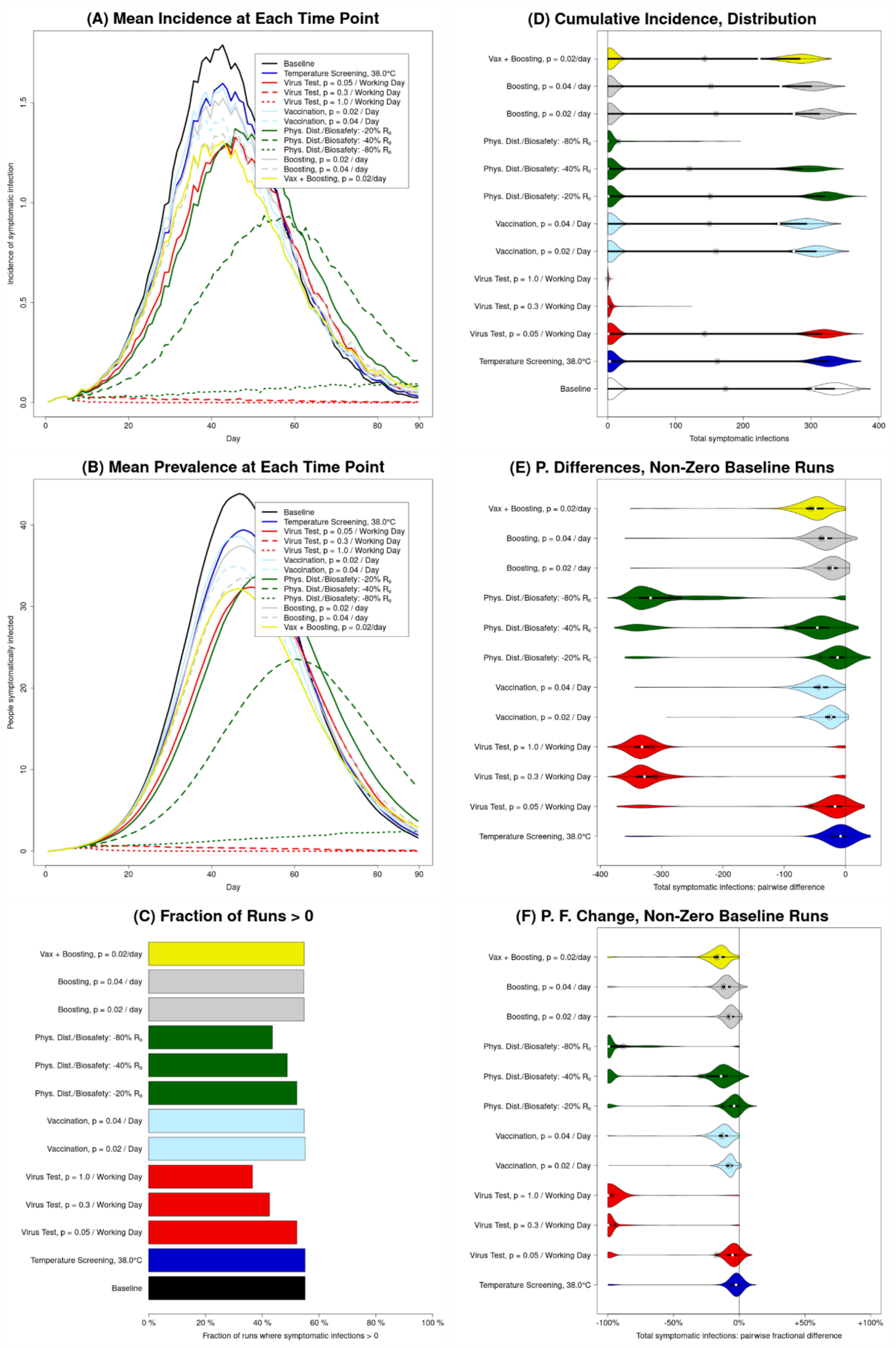
(A) Mean Incidence at Each Time Point
Incidence of symptomatic infection
Day
Baseline
Temperature Screening, 38.0°C
Virus Test, p = 0.05 / Working Day
Virus Test, p = 0.3 / Working Day
Virus Test, p = 1.0 / Working Day
Vaccination, p = 0.02 / Day
Vaccination, p = 0.04 / Day
Phys. Dist./Biosafety: -20% R0
Phys. Dist./Biosafety: -40% R0
Phys. Dist./Biosafety: -80% R0
Boosting, p = 0.02 / day
Boosting, p = 0.04 / day
Vax + Boosting, p = 0.02/day
(B) Mean Prevalence at Each Time Point
People symptomatically infected
Day
(C) Fraction of Runs > 0
Fraction of runs where symptomatic infections > 0
(D) Cumulative Incidence, Distribution
Total symptomatic infections
(E) P. Differences, Non-Zero Baseline Runs
Total symptomatic infections: pairwise difference
(F) P. F. Change, Non-Zero Baseline Runs
Total symptomatic infections: pairwise fractional difference

**Fig. S2. Illustration of public health outcomes for a large facility (1,003 employees) for baseline (no intervention), for each of the interventions in absolute terms as well as relative to the baseline**

Results for number of symptomatic infections in a processing facility with 1,003 employees over the 90 days of the simulation run are shown (results for total infections are similar, apart from scale). (A and B) The mean across all runs of the incidence (A) and prevalence (B) of symptomatic infection, at each time point; these illustrate the dynamics over time, but also conceal the high level of variation between runs. (C) The fraction of runs for which the total number of symptomatic infections is greater than zero. (D) Violin plots representing the distribution, between runs, of the *total* number of symptomatic infections; these violin plots illustrate the bimodal nature of most distributions. (E) Violin plots representing the distribution of *counterfactual effects* of the various interventions, i.e., the distribution of *pairwise differences* between *corresponding* runs with and without that intervention (the number at that intervention ($N_I$) minus the number at baseline ($N_B$); ($N_I - N_B$)), for runs that *do* have one or more symptomatic infections at baseline. (F) Violin plots representing the distribution of pairwise *fractional* differences (i.e., $(N_I - N_B)/N_B$), for runs with a non-zero number of symptomatic infections at baseline.

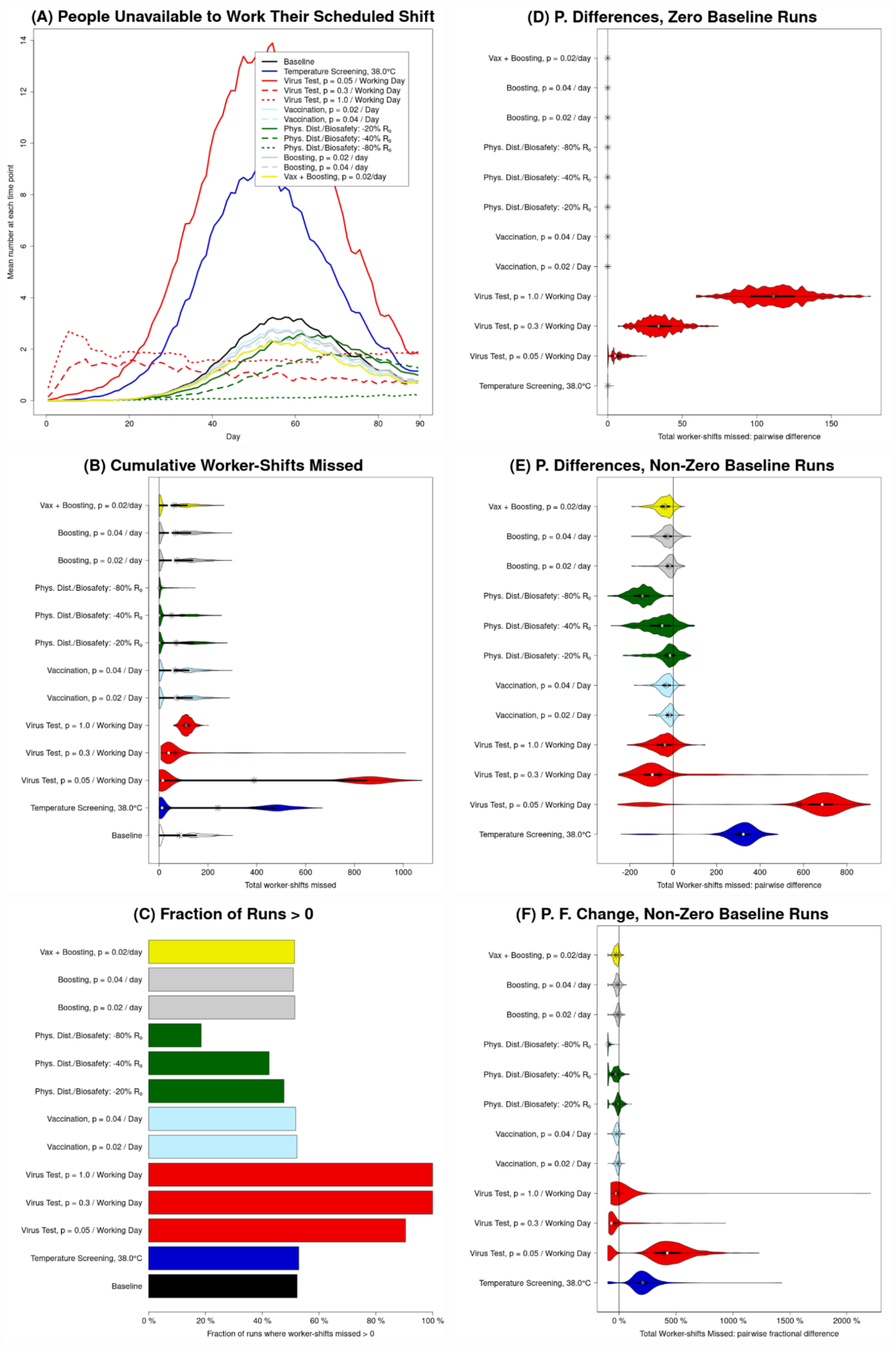
(A) People Unavailable to Work Their Scheduled Shift
Mean number at each time point
Day
Baseline
Temperature Screening, 38.0°C
Virus Test, p = 0.05 / Working Day
Virus Test, p = 0.3 / Working Day
Virus Test, p = 1.0 / Working Day
Vaccination, p = 0.02 / Day
Vaccination, p = 0.04 / Day
Phys. Dist./Biosafety: -20% R0
Phys. Dist./Biosafety: -40% R0
Phys. Dist./Biosafety: -80% R0
Boosting, p = 0.02 / day
Boosting, p = 0.04 / day
Vax + Boosting, p = 0.02/day
(B) Cumulative Worker-Shifts Missed
Total worker-shifts missed
(C) Fraction of Runs > 0
Fraction of runs where worker-shifts missed > 0
(D) P. Differences, Zero Baseline Runs
Total worker-shifts missed: pairwise difference
(E) P. Differences, Non-Zero Baseline Runs
Total Worker-shifts missed: pairwise difference
(F) P. F. Change, Non-Zero Baseline Runs
Total Worker-shifts Missed: pairwise fractional difference

**Fig. S3. Illustration of unavailability for a large facility (1,003 employees) for baseline (no intervention), for each of the interventions in absolute terms as well as relative to the baseline**

Unavailability (i.e., worker-shifts missed) depends not only on how many employees are infected, and how many of those are symptomatic, but also on how likely an infected employee (whether symptomatic or asymptomatic) is to be removed from the workforce (due to hospitalization, or to detection and isolation). All results are for 90-day long simulation runs. (A) The mean across all runs of the number of employees unavailable to work their scheduled production shift, for each day of the simulation; this illustrates the dynamics over time, but also conceals the substantial level of variation between runs. (B) Violin plots representing the distribution, between runs, of the sum of the number of workers unavailable to work their scheduled production shift, over all such shifts; this violin plot illustrates the varying shapes of these distributions. (C) The fraction of runs for which the total number of worker-shifts missed is greater than zero. (D and E) Violin plots representing the distribution of *counterfactual effects* of the various interventions, i.e., the distribution of *pairwise differences* between *corresponding* runs with and without that intervention (the number at that intervention ($N_I$) minus the number at baseline ($N_B$); ($N_I - N_B$)), for runs with zero (panel D) and non-zero (panel E) worker-shifts missed at baseline. (F) Violin plots representing the distribution of pairwise *fractional* differences (i.e., $(N_I - N_B)/N_B$), for runs with non-zero worker-shifts missed at baseline.

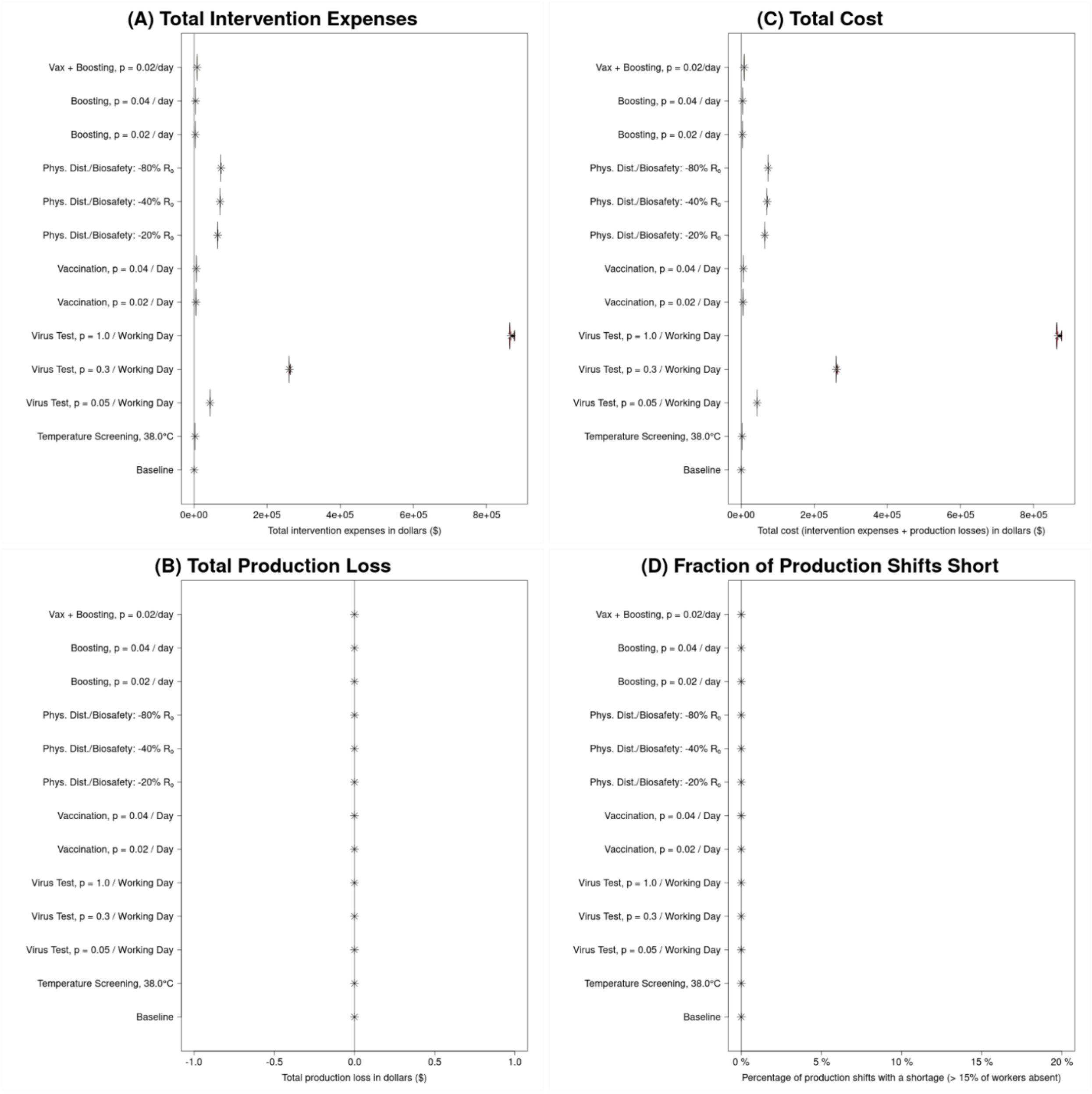

**Fig. S4. Illustration of costs for a large facility (1,003 employees) for baseline (no intervention) and each of the interventions**

All panels consist of violin plots (although in some cases, these may be sufficiently horizontally compressed that this is not obvious) representing the distribution (across runs within an intervention) of an outcome. All results are for 90-day long simulation runs. (A) Distribution of direct intervention expenses (supplies purchased and/or additional wages paid for tasks performed outside of an individual's normal scheduled working hours); these are generally relatively constant for an intervention, and are always US$0 by definition for the baseline. (B) Distribution of production losses due to worker absences; as a result of how we model unavailability (occurring only on days when >15% of workers miss their shift) this is almost always US$0 in the absence of a testing intervention. In this larger facility, and with otherwise default parameters, we do not see production losses in the presence of low-to-moderate levels of testing, unlike in the smaller facility examined in the main text. This is essentially because the average level of unavailability at a given time is less than 15% even in the smaller facility, but stochastic variation is easily capable of pushing it over that line in a non-trivial number of runs. In the larger facility, the ability of stochastic variation to cross that (correspondingly higher) threshold is reduced. (C) Distribution of total costs (in US$), which we define to be the sum of intervention expenses and production losses. (D) Fraction of production shifts (within a single run) that are "short", i.e., more than 15% of workers absent ("0%" means that in a particular run none of the shifts were "short").

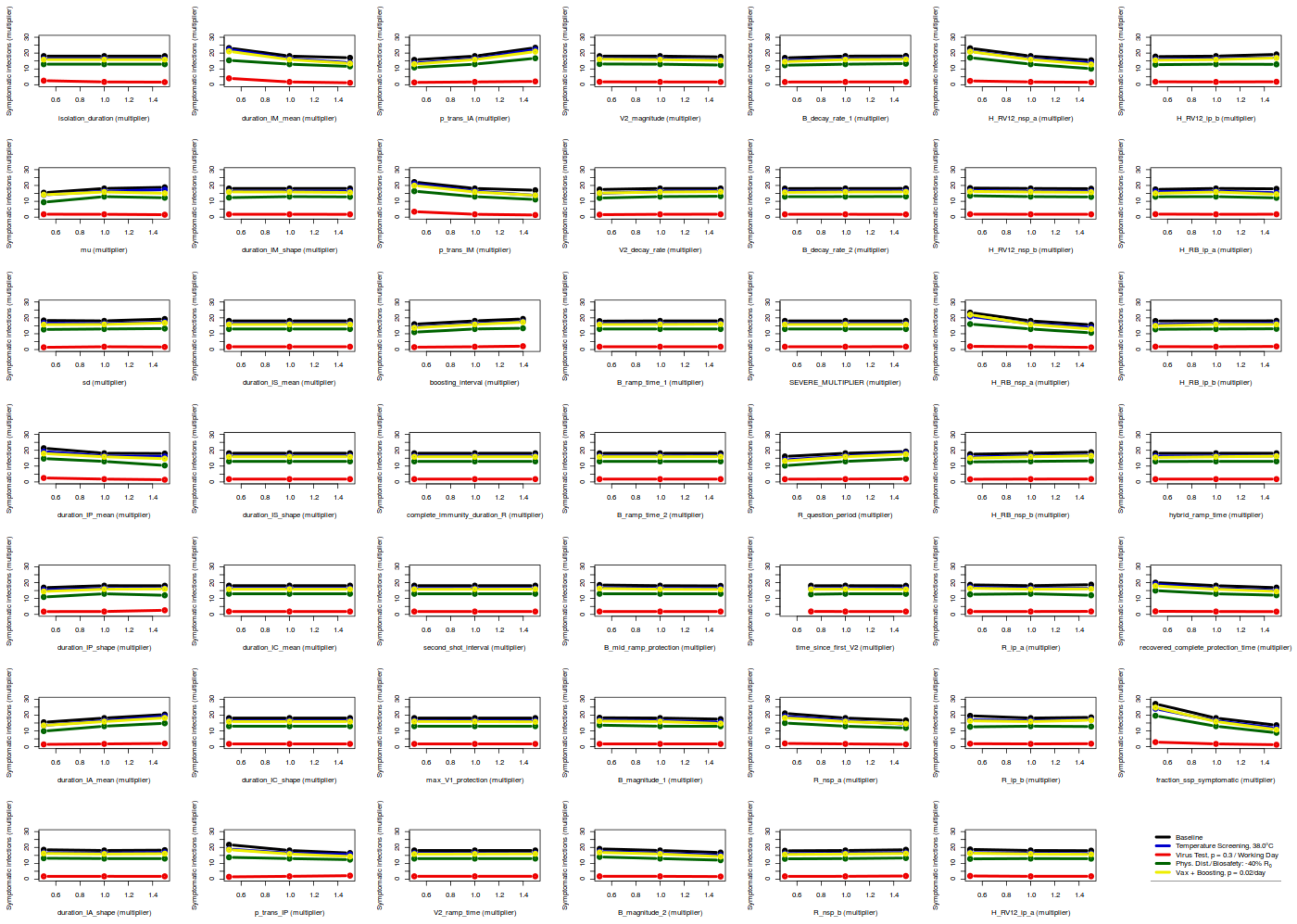

**Fig. S5. Plot of sensitivities of the number of symptomatic infections with respect to all sensitivity parameters**

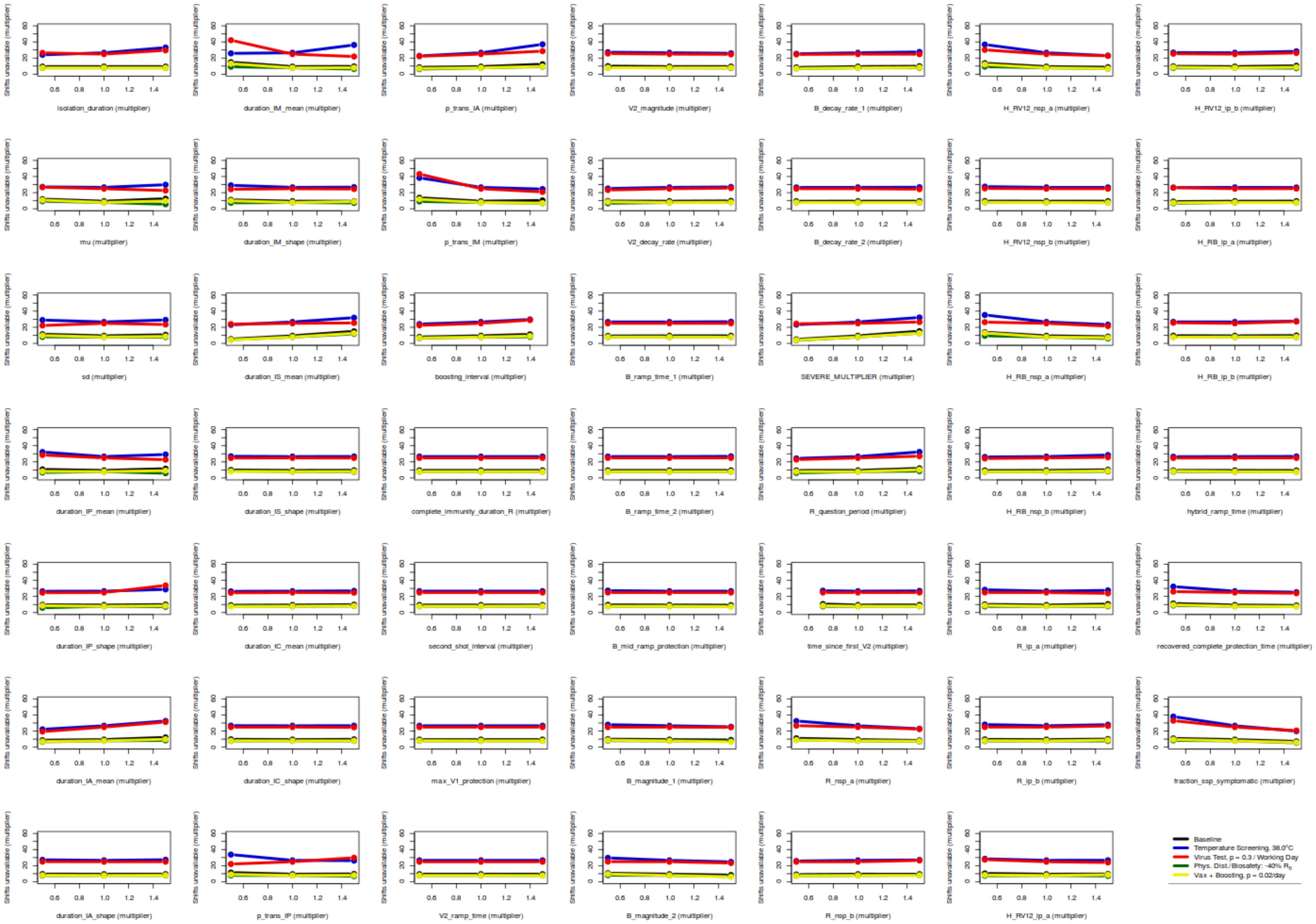

**Fig. S6. Plot of sensitivities of the number of worker-shifts unavailable with respect to all sensitivity parameters**

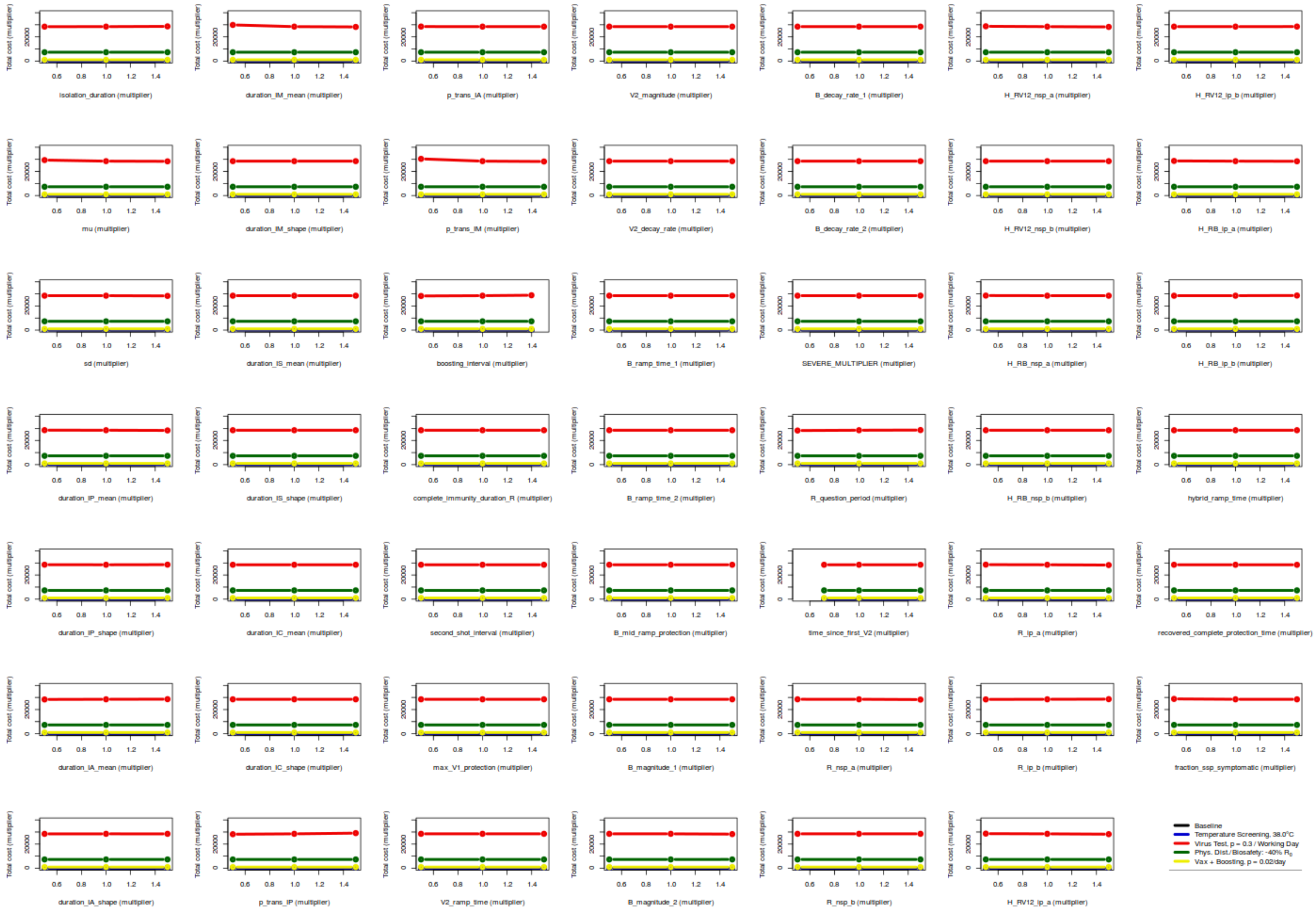

**Fig. S7. Plot of sensitivities of the total cost with respect to all sensitivity parameters**

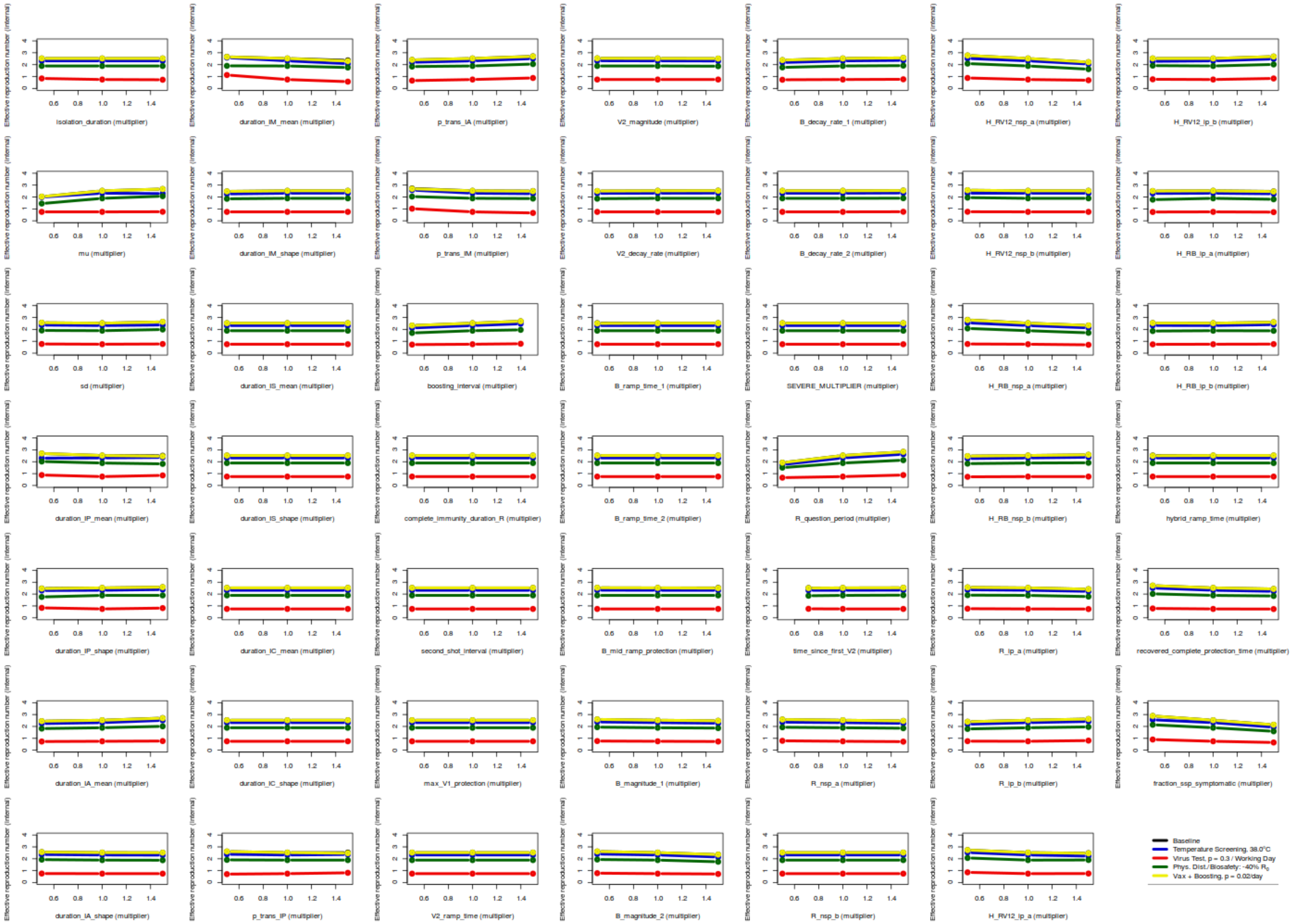

**Fig. S8. Plot of sensitivities of the effective reproduction number with respect to all sensitivity parameters**

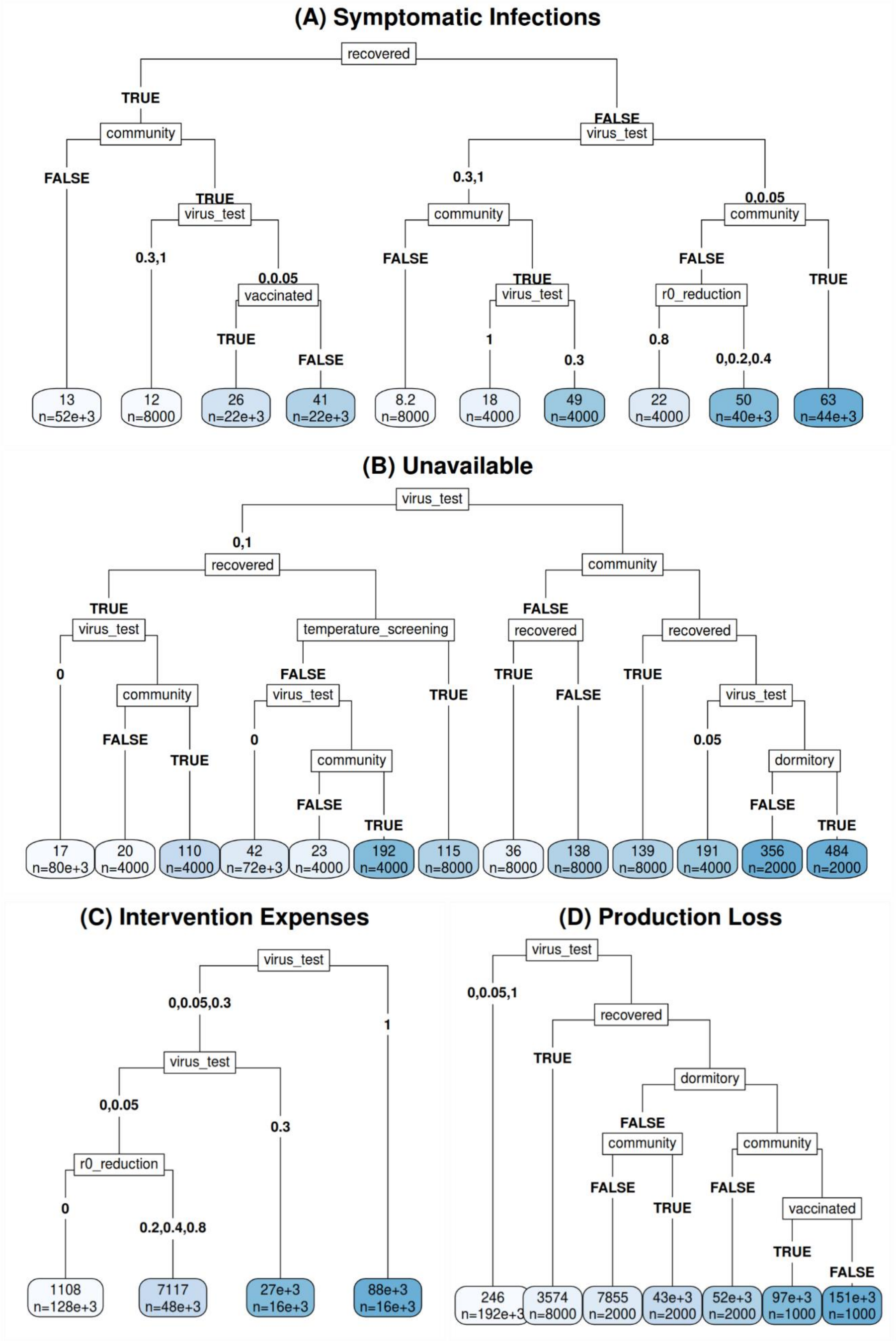
(A) Symptomatic Infections
recovered
TRUE
FALSE
community
virus_test
FALSE
TRUE
0.3,1
0,0.05
virus_test
community
community
0.3,1
0,0.05
FALSE
TRUE
FALSE
TRUE
vaccinated
virus_test
r0_reduction
TRUE
FALSE
1
0.3
0.8
0,0.2,0.4
13
n=52e+3
12
n=8000
26
n=22e+3
41
n=22e+3
8.2
n=8000
18
n=4000
49
n=4000
22
n=4000
50
n=40e+3
63
n=44e+3
(B) Unavailable
virus_test
0,1
recovered
community
TRUE
FALSE
virus_test
temperature_screening
recovered
recovered
0
community
FALSE
TRUE
TRUE
FALSE
TRUE
FALSE
TRUE
virus_test
virus_test
0
0.05
community
dormitory
FALSE
TRUE
FALSE
TRUE
17
n=80e+3
20
n=4000
110
n=4000
42
n=72e+3
23
n=4000
192
n=4000
115
n=8000
36
n=8000
138
n=8000
139
n=8000
191
n=4000
356
n=2000
484
n=2000
(C) Intervention Expenses
virus_test
0,0.05,0.3
1
virus_test
0,0.05
0.3
r0_reduction
0
0.2,0.4,0.8
1108
n=128e+3
7117
n=48e+3
27e+3
n=16e+3
88e+3
n=16e+3
(D) Production Loss
virus_test
0,0.05,1
recovered
TRUE
dormitory
FALSE
community
community
FALSE
TRUE
FALSE
vaccinated
TRUE
FALSE
246
n=192e+3
3574
n=8000
7855
n=2000
43e+3
n=2000
52e+3
n=2000
97e+3
n=1000
151e+3
n=1000

**Fig. S9. Regression Trees for a variation of the model in which the presence ("True") or absence ("False") of community transmission and dormitory transmission are allowed to vary independently**

For all panels, the labels on the branches descending from a node represent values of the parameter listed in the node itself. Where there are only two values for a parameter, the value is sometimes only listed explicitly on the left branch, to save space; the right branch simply has the value of that parameter that the left branch does not. In all cases, branches are ordered by making the left branch the one with the *lower* average value for the outcome represented in that panel. (This does not, however, result in *all* leaves being ordered from lowest to highest, because the branches are not allowed to cross.) The value at each leaf indicates the mean value of the outcome across relevant scenarios x interventions x runs over the 90 days of the simulation run, and *n* indicates the number of runs represented by the leaf (out of 104,000 runs). (A) Total number (Cumulative Incidence) of Symptomatic Infections, (B) Total number of Worker-Shifts Unavailable, (C) Cumulative intervention expenses (in US$), (D) Cumulative production losses (in US$).

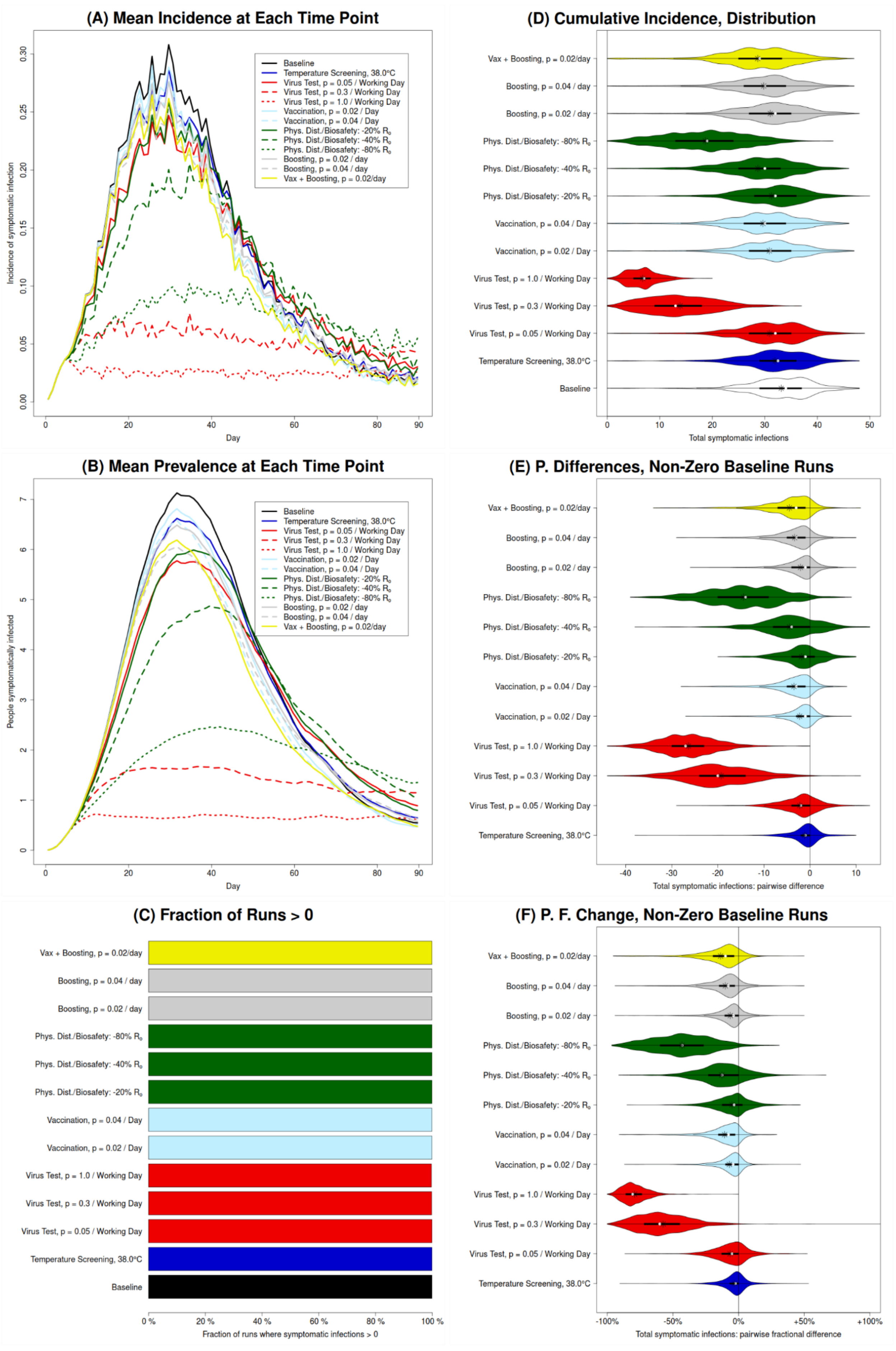
(A) Mean Incidence at Each Time Point
Baseline
Temperature Screening, 38.0°C
Virus Test, p = 0.05 / Working Day
Virus Test, p = 0.3 / Working Day
Virus Test, p = 1.0 / Working Day
Vaccination, p = 0.02 / Day
Vaccination, p = 0.04 / Day
Phys. Dist./Biosafety: -20% R0
Phys. Dist./Biosafety: -40% R0
Phys. Dist./Biosafety: -80% R0
Boosting, p = 0.02 / day
Boosting, p = 0.04 / day
Vax + Boosting, p = 0.02/day
Incidence of symptomatic infection
0.00
0.05
0.10
0.15
0.20
0.25
0.30
Day
0
20
40
60
80
90
(D) Cumulative Incidence, Distribution
Vax + Boosting, p = 0.02/day
Boosting, p = 0.04 / day
Boosting, p = 0.02 / day
Phys. Dist./Biosafety: -80% R0
Phys. Dist./Biosafety: -40% R0
Phys. Dist./Biosafety: -20% R0
Vaccination, p = 0.04 / Day
Vaccination, p = 0.02 / Day
Virus Test, p = 1.0 / Working Day
Virus Test, p = 0.3 / Working Day
Virus Test, p = 0.05 / Working Day
Temperature Screening, 38.0°C
Baseline
0
10
20
30
40
50
Total symptomatic infections
(B) Mean Prevalence at Each Time Point
Baseline
Temperature Screening, 38.0°C
Virus Test, p = 0.05 / Working Day
Virus Test, p = 0.3 / Working Day
Virus Test, p = 1.0 / Working Day
Vaccination, p = 0.02 / Day
Vaccination, p = 0.04 / Day
Phys. Dist./Biosafety: -20% R0
Phys. Dist./Biosafety: -40% R0
Phys. Dist./Biosafety: -80% R0
Boosting, p = 0.02 / day
Boosting, p = 0.04 / day
Vax + Boosting, p = 0.02/day
People symptomatically infected
0
1
2
3
4
5
6
7
Day
0
20
40
60
80
90
(E) P. Differences, Non-Zero Baseline Runs
Vax + Boosting, p = 0.02/day
Boosting, p = 0.04 / day
Boosting, p = 0.02 / day
Phys. Dist./Biosafety: -80% R0
Phys. Dist./Biosafety: -40% R0
Phys. Dist./Biosafety: -20% R0
Vaccination, p = 0.04 / Day
Vaccination, p = 0.02 / Day
Virus Test, p = 1.0 / Working Day
Virus Test, p = 0.3 / Working Day
Virus Test, p = 0.05 / Working Day
Temperature Screening, 38.0°C
-40
-30
-20
-10
0
10
Total symptomatic infections: pairwise difference
(C) Fraction of Runs > 0
Vax + Boosting, p = 0.02/day
Boosting, p = 0.04 / day
Boosting, p = 0.02 / day
Phys. Dist./Biosafety: -80% R0
Phys. Dist./Biosafety: -40% R0
Phys. Dist./Biosafety: -20% R0
Vaccination, p = 0.04 / Day
Vaccination, p = 0.02 / Day
Virus Test, p = 1.0 / Working Day
Virus Test, p = 0.3 / Working Day
Virus Test, p = 0.05 / Working Day
Temperature Screening, 38.0°C
Baseline
0 %
20 %
40 %
60 %
80 %
100 %
Fraction of runs where symptomatic infections > 0
(F) P. F. Change, Non-Zero Baseline Runs
Vax + Boosting, p = 0.02/day
Boosting, p = 0.04 / day
Boosting, p = 0.02 / day
Phys. Dist./Biosafety: -80% R0
Phys. Dist./Biosafety: -40% R0
Phys. Dist./Biosafety: -20% R0
Vaccination, p = 0.04 / Day
Vaccination, p = 0.02 / Day
Virus Test, p = 1.0 / Working Day
Virus Test, p = 0.3 / Working Day
Virus Test, p = 0.05 / Working Day
Temperature Screening, 38.0°C
-100%
-50%
0%
+50%
+100%
Total symptomatic infections: pairwise fractional difference

**Fig. S10. Illustration of public health outcomes for baseline (no intervention) and each of the interventions in absolute terms as well as relative to the baseline, for a scenario with both community and dormitory transmission**

Results for number of symptomatic infections in a processing facility with 103 employees over the 90 days of the simulation run are shown (results for total infections are similar, apart from scale). (A and B) The mean across all runs of the incidence (A) and prevalence (B) of symptomatic infection, at each time point; these illustrate the dynamics over time, but also conceal the high level of variation between runs. (C) The fraction of runs for which the total number of symptomatic infections is greater than zero. (D) Violin plots representing the distribution, between runs, of the *total* number of symptomatic infections; these violin plots illustrate the bimodal nature of most distributions. (E) Violin plots representing the distribution of *counterfactual effects* of the various interventions, i.e., the distribution of *pairwise differences* between *corresponding* runs with and without that intervention (the number at that intervention ($N_I$) minus the number at baseline ($N_B$); ($N_I - N_B$)), for runs that *do* have one or more symptomatic infections at baseline. (F) Violin plots representing the distribution of pairwise *fractional* differences (i.e., $(N_I - N_B)/N_B$), for runs with a non-zero number of symptomatic infections at baseline. For one of the interventions, there is a single positive outlier that is cut off by the axis limits to avoid excessively compressing the depiction of the other 11,999 points.

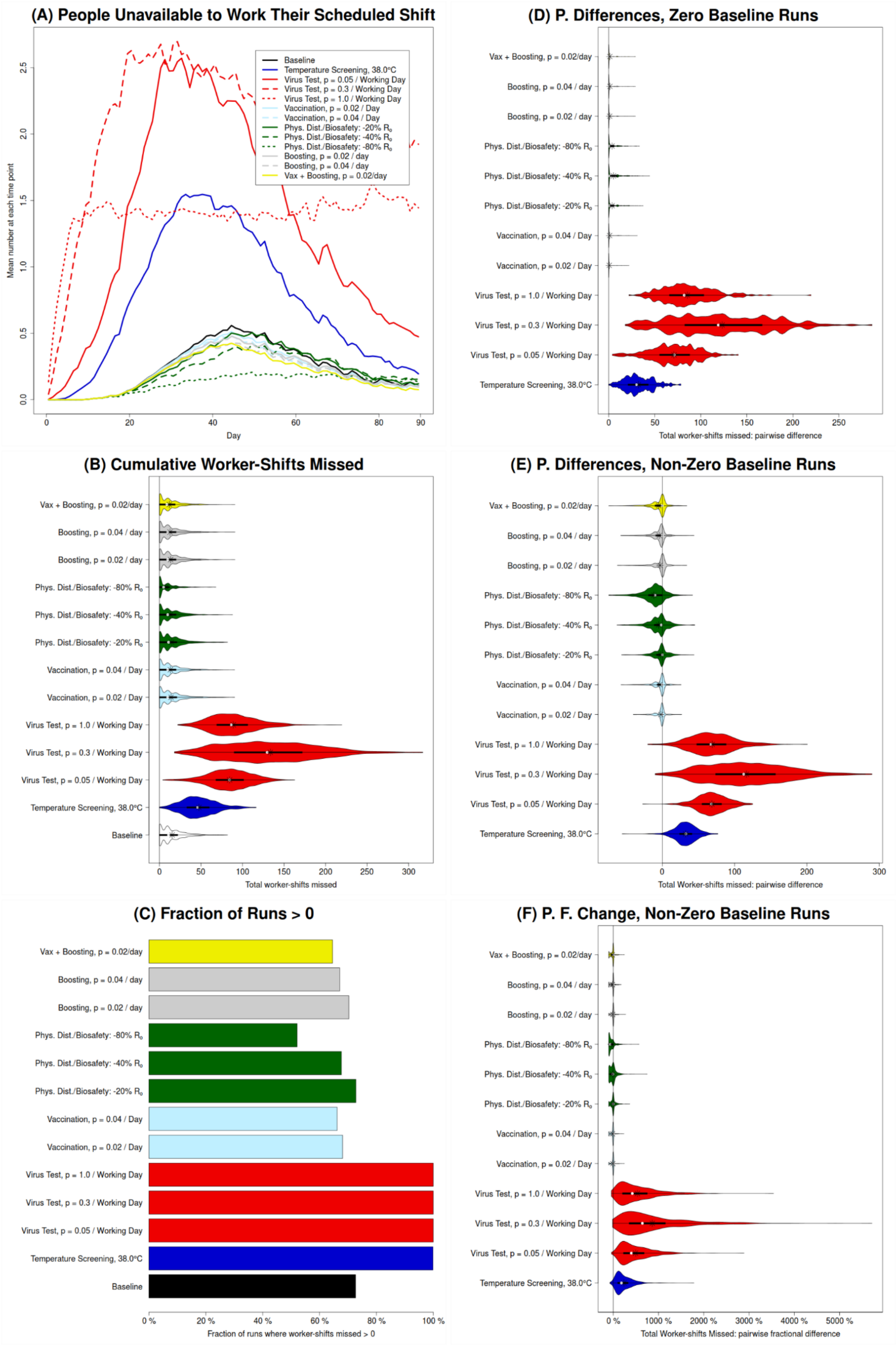

(A) People Unavailable to Work Their Scheduled Shift
Baseline
Temperature Screening, 38.0°C
Virus Test, p = 0.05 / Working Day
Virus Test, p = 0.3 / Working Day
Virus Test, p = 1.0 / Working Day
Vaccination, p = 0.02 / Day
Vaccination, p = 0.04 / Day
Phys. Dist./Biosafety: -20% $R_0$
Phys. Dist./Biosafety: -40% $R_0$
Phys. Dist./Biosafety: -80% $R_0$
Boosting, p = 0.02 / day
Boosting, p = 0.04 / day
Vax + Boosting, p = 0.02/day
Mean number at each time point
Day
(D) P. Differences, Zero Baseline Runs
Total worker-shifts missed: pairwise difference
(B) Cumulative Worker-Shifts Missed
Total worker-shifts missed
(E) P. Differences, Non-Zero Baseline Runs
Total Worker-shifts missed: pairwise difference
(C) Fraction of Runs > 0
Fraction of runs where worker-shifts missed > 0
(F) P. F. Change, Non-Zero Baseline Runs
Total Worker-shifts Missed: pairwise fractional difference

**Fig. S11. Illustration of unavailability for baseline (no intervention) and each of the interventions in absolute terms as well as relative to the baseline, for a scenario with both community and dormitory transmission**

Unavailability (i.e., worker-shifts missed) depends not only on how many employees are infected, and how many of those are symptomatic, but also on how likely an infected employee (whether symptomatic or asymptomatic) is to be removed from the workforce (due to hospitalization, or to detection and isolation). All results are for 90-day long simulation runs. (A) The mean across all runs of the number of employees unavailable to work their scheduled production shift, for each day of the simulation; this illustrates the dynamics over time, but also conceals the substantial level of variation between runs. (B) Violin plots representing the distribution, between runs, of the sum of the number of workers unavailable to work their scheduled production shift, over all such shifts; this violin plot illustrates the varying shapes of these distributions. (C) The fraction of runs for which the total number of worker-shifts missed is greater than zero. (D and E) Violin plots representing the distribution of *counterfactual effects* of the various interventions, i.e., the distribution of *pairwise differences* between *corresponding* runs with and without that intervention (the number at that intervention ($N_I$) minus the number at baseline ($N_B$); ($N_I - N_B$)), for runs with zero (panel D) and non-zero (panel E) worker-shifts missed at baseline. (F) Violin plots representing the distribution of pairwise *fractional* differences (i.e., $(N_I - N_B)/N_B$), for runs with non-zero worker-shifts missed at baseline.

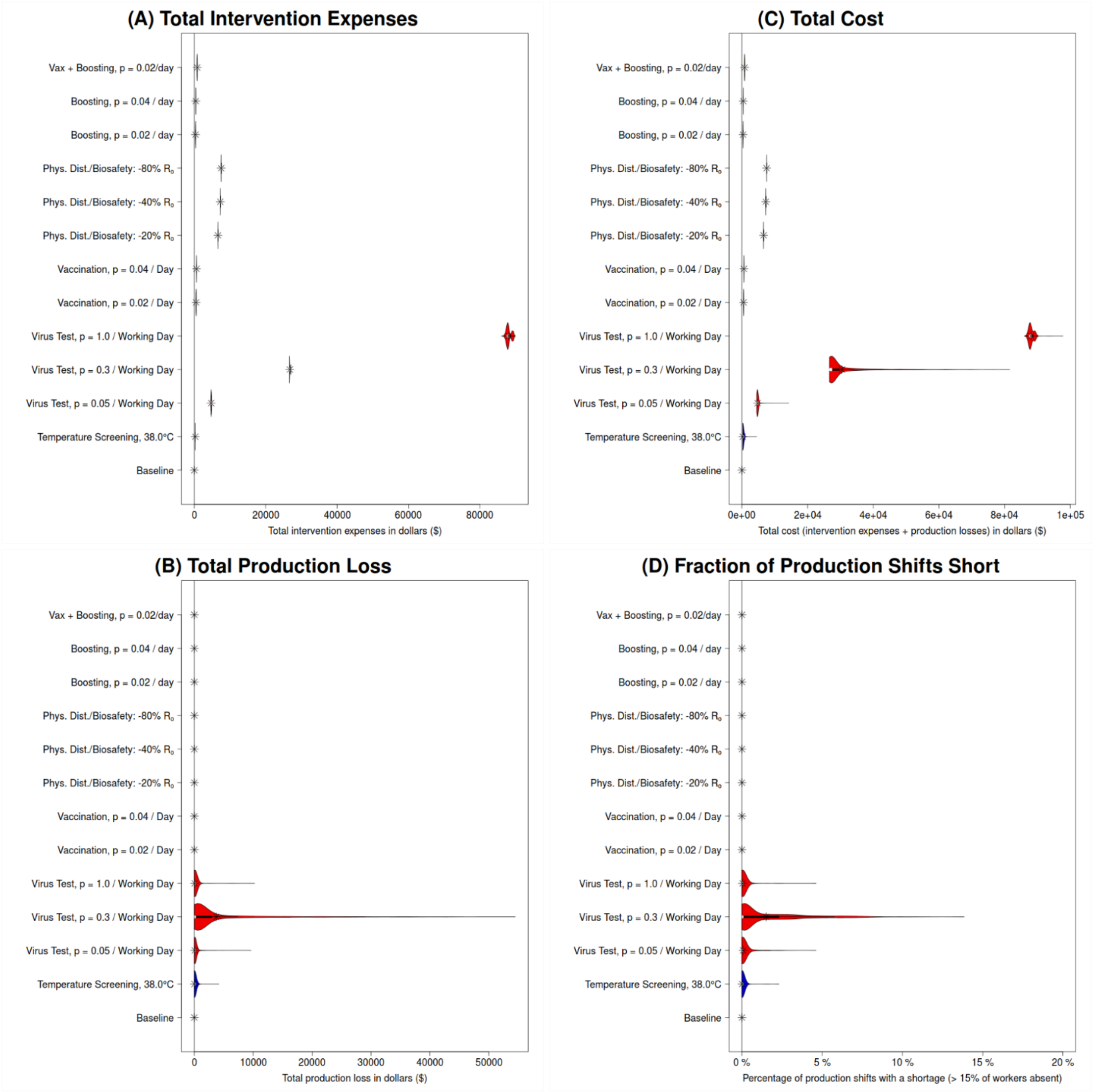

**Fig. S12. Illustration of costs for baseline (no intervention) and each of the interventions, for a scenario with both community and dormitory transmission**

All panels consist of violin plots (although in some cases, these may be sufficiently horizontally compressed that this is not obvious) representing the distribution (across runs within an intervention) of an outcome. All results are for 90-day long simulation runs. (A) Distribution of direct intervention expenses (supplies purchased and/or additional wages paid for tasks performed outside of an individual's normal scheduled working hours); these are generally relatively constant for an intervention, and are always US$0 by definition for the baseline. (B) Distribution of production losses due to worker absences; as a result of how we model unavailability (occurring only on days when >15% of workers miss their shift) this is almost always US$0 in the absence of a testing intervention. Here, we can see that low to moderate levels of routine viral testing may be insufficient to interrupt transmission, but sufficient to remove significant numbers of employees through detection and isolation, and thus causing significant production losses. (C) Distribution of total costs (in US$), which we define to be the sum of intervention expenses and production losses. (D) Fraction of production shifts (within a single run) that are "short", i.e., more than 15% of workers absent ("0%" means that in a particular run none of the shifts were "short").

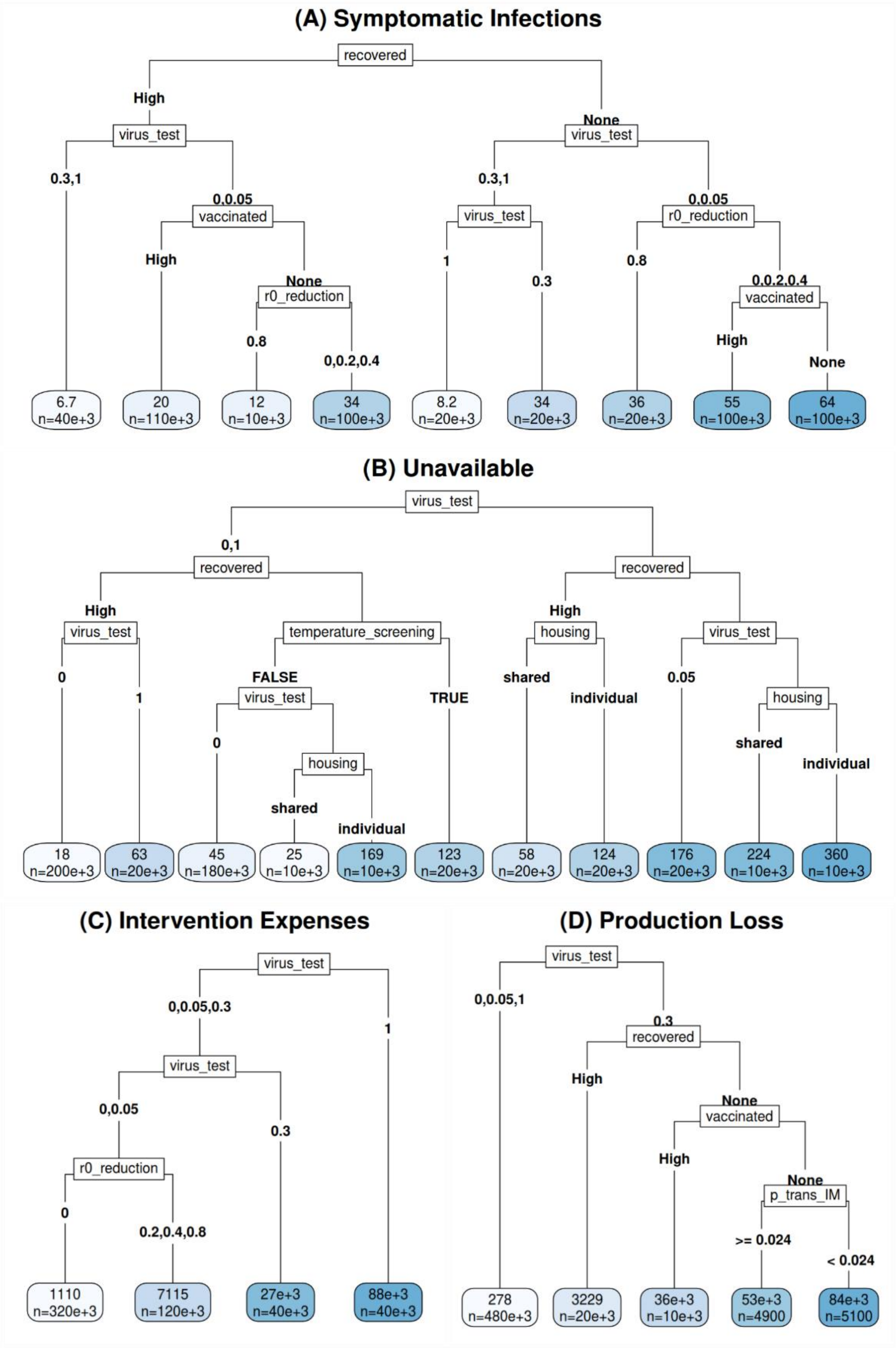

**Fig. S13. Regression Trees for multivariable analysis of setting and intervention parameters and the five most sensitive uncertain parameters (from Figure 6).**

For all panels, the labels on the branches descending from a node represent values of the parameter listed in the node itself. In all cases, branches are ordered by making the left branch the one with the *lower* average value for the outcome represented in that panel. Each leaf represents the collection of all runs (out of 100 for each combination of one of 8 setting scenarios, one of 50 sets of sensitivity parameters for that scenario, and one of 13 interventions, for a total of 520,000 runs) that meet the criteria specified by the nodes and branches ancestral to that leaf. The value in each leaf represents the mean value of that panel's outcome over all runs in that collection, measured over the 90 days of the simulation run, and *n* indicates the number of runs in that collection. (A) Total number (Cumulative Incidence) of Symptomatic Infections, (B) Total number of Worker-Shifts Unavailable, (C) Cumulative intervention expenses (in US$), (D) Cumulative production losses (in US$).

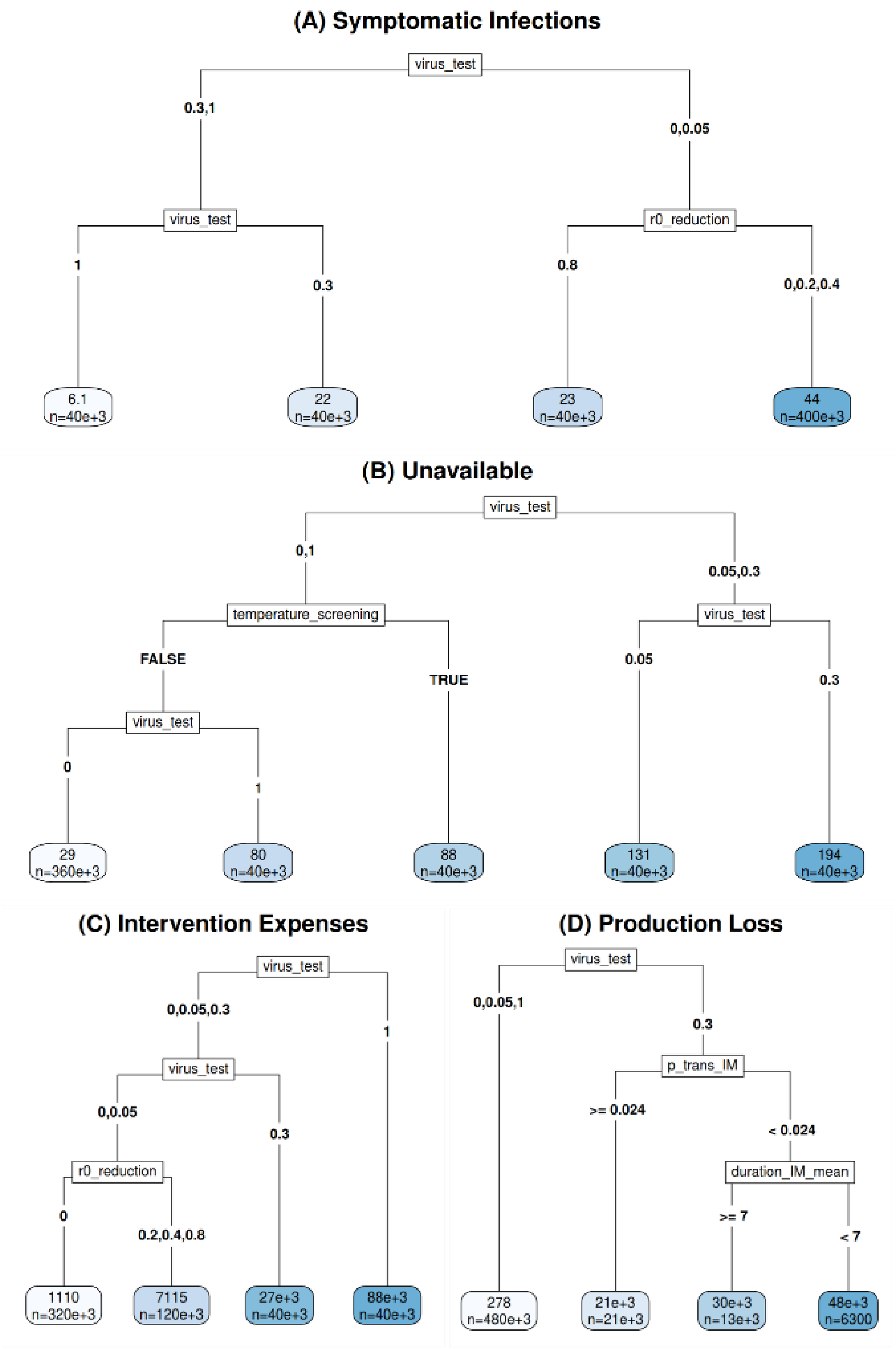

**Fig. S14. Regression Trees for multivariable analysis of intervention parameters and the five most sensitive uncertain parameters (from Figure 6).**

For all panels, the labels on the branches descending from a node represent values of the parameter listed in the node itself. Although the three setting scenario parameters were varied between different runs as in **Fig. S13**, they were excluded from use constructing recursion trees.

In all cases, branches are ordered by making the left branch the one with the *lower* average value for the outcome represented in that panel. Each leaf represents the collection of all runs (out of 100 for each combination of one of 8 scenarios, one of 50 sets of sensitivity parameters for that scenario, and one of 13 interventions, for a total of 520,000 runs) that meet the criteria specified by the nodes and branches ancestral to that leaf. The value in each leaf represents the mean value of that panel's outcome over all runs in that collection, measured over the 90 days of the simulation run, and *n* indicates the number of runs in that collection. (A) Total number (Cumulative Incidence) of Symptomatic Infections, (B) Total number of Worker-Shifts Unavailable, (C) Cumulative intervention expenses (in US$), (D) Cumulative production losses (in US$).

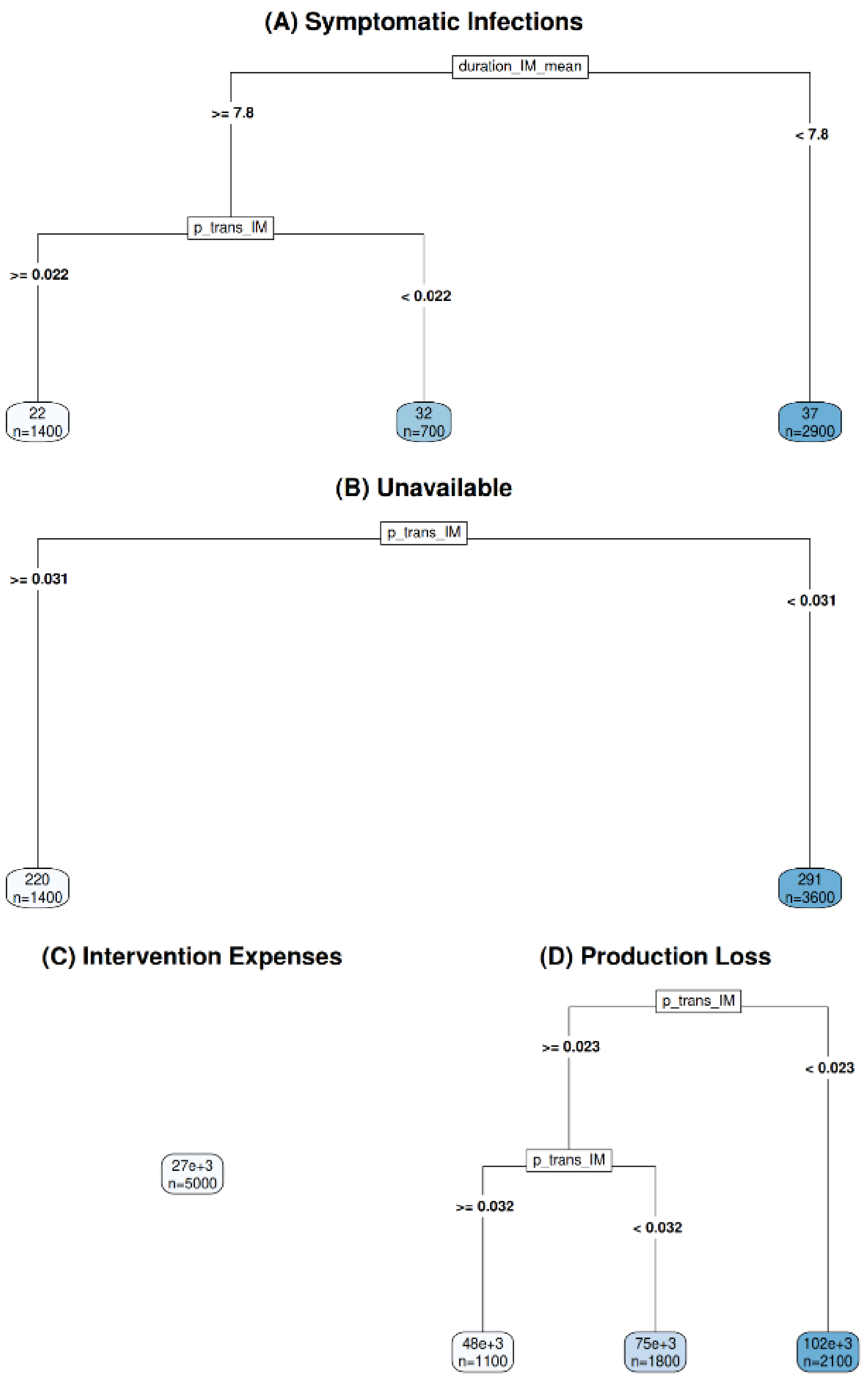

**Fig. S15. Regression Trees for multivariable analysis of the five most sensitive uncertain parameters (from Figure 6) in a particular setting-intervention scenario for a facility with no vaccinated and recovered employees and with shared housing and moderate viral testing (*p* = 0.3/work day).**

For all panels, the labels on the branches descending from a node represent values of the parameter listed in the node itself. In all cases, branches are ordered by making the left branch the one with the *lower* average value for the outcome represented in that panel. Each leaf represents the collection of all runs (out of 100 for each of 50 sets of sensitivity parameters, for a total of 5000 runs). The value in each leaf represents the mean value of that panel's outcome over all runs in that collection, measured over the 90 days of the simulation run, and *n* indicates the number of runs in that collection. (A) Total number (Cumulative Incidence) of Symptomatic Infections, (B) Total number of Worker-Shifts Unavailable, (C) Cumulative intervention expenses (in US$), (D) Cumulative production losses (in US$).

### Supplementary tables

#### Table S1. Numbers of agents with various immune states and histories derived from user-set parameters

Additional details, including the equations for the calculation of $N$ from user-set parameters, can be found in **Text S2** and **Table S4**. In the rightmost column, equations are presented for how past event times ($t_{R,i}$, $t_{V2,i}$, and/or $t_{B,i}$, as applicable and relevant) and boosting status or "intention" ($B_{on\ time,i}$) are set for individuals encompassed in each of the described counts. As some of these counts are nested within other counts (e.g, $N_{B,\ recent}$ within $N_B$ within $N_{V2,\ older}$), these attribute distributions are specified at the highest level for each they are consistent. For individuals who have received the second dose of their two-dose primary vaccination series at least $T_{V2\rightarrow B}$ ago, $B_{on\ time,i}$ indicates whether that individual *received* a booster dose "on time" when they became eligible to receive one (i.e., $T_{V2\rightarrow B}$ after the second dose of their primary series). For individuals who have completed their primary series less than $T_{V2\rightarrow B}$ ago, or who have not completed their primary series at all, it indicates whether they *will receive* a booster dose "on time," if and when they become eligible to receive one.

| **Symbol** | **Description** | **Value** | **Associated equation(s), for an agent *i* who is part of this count** |
|---|---|---|---|
| $N$ | Total number of agents (employee) | Farm: ≥4<br>Facility: ≥7 | |
| $N_R(0)$ | Number of agents who have recovered from natural infection within the last year before the simulation start | round($N$ * $f_R$) | $t_{R,i}$ ~ Uniform($-T_{RQ}$, 0) |
| $N_{V2}(0)$ | Number of agents who have completed a course of primary vaccination at simulation start | round($N$ * $f_{V2}$) | |
| $N_{V2,\ recent}(0)$ | Number of agents who have completed a course of primary vaccination *less than* $T_{V2\rightarrow B}$ = 152 days (5 months) prior to the start of simulation | round($N$ * $f_{V2,\ recent}$) | $t_{V2,i}$ ~ Uniform($-T_{V2\rightarrow B}$, 0)<br>$B_{on\ time,i}$ ~ Bernoulli($f_B$) |
| $N_{V2,\ older}(0)$ | Number of agents who have completed a course of primary vaccination *more than* $T_{V2\rightarrow B}$ prior to the start of simulation | $N_{V2}(0)$ - $N_{V2,\ recent}(0)$ | |
| $N_B(0)$ | Number of agents who have received a booster dose | $f$B * $N_{V2,\ older}$ | $B_{on\ time,i}$ = 1 |
| $N_{B,\ recent}(0)$ | Number of agents who have received a booster dose *less than* $T_{V2\rightarrow B}$ prior to the start of simulation | $f_{B,\ recent}$ * $N_{V2,\ older}$ | $t_{V2,i}$ ~ Uniform(-2 * $T_{V2\rightarrow B}$, $-T_{V2\rightarrow B}$)<br>$t_{B,i}$ = $t_{V2,i}$ + $T_{V2\rightarrow B}$ |
| $N_{B,older}(0)$ | Number of agents who have received a booster dose *more than* $T_{V2\rightarrow B}$ prior to the start of simulation | $N_B(0)$ - $N_{B,\ recent}(0)$ | $t_{V2,i}$ ~ Uniform($-T_{1st\ V2\ ->\ 0}$, -(2 * $T_{V2\rightarrow B}$ + 1))<br>$t_{B,i}$ = $t_{V2,i}$ + $T_{V2\rightarrow B}$ |
| $N_{V2,older,no\ booster}$ | Number of agents who have completed a course of primary vaccination *more than* $T_{V2\rightarrow B}$ prior to the start of simulation but have *not* received a booster (despite presumably being eligible) | $N_{V2,older}$ - $N_B(0)$ | $t_{V2,i}$ ~ Uniform($-T_{1st\ V2\ ->\ 0}$, $-T_{V2\rightarrow B}$)<br>$B_{on\ time,i}$ = 0 |
| $N_{no\ V2}$ | Number of agents who have not completed a | $N$ - $N_{V2}$ | $B_{on\ time,i}$ ~ Bernoulli($f_B$) |

| | |
|---|---|
| | primary course of vaccination |

**Table S2. Model parameters**

These parameters are also relevant to sensitivity analysis. Gamma distributions are notated Gamma(shape, scale); when the mean is varied in sensitivity analysis, this is done by varying the scale parameter, while holding the shape parameter fixed. For more details, see **Text S10**. In the case of $\beta_{IM}$, it is the ratios between $\beta_{IM}$ and the corresponding parameters for asymptomatic and presymptomatic infection ($\beta_{IA}$ and $\beta_{IP}$) that are taken from Moghadas et al. 2020 [*51*]; the absolute magnitudes are rendered irrelevant by how we set contact rates. The derivation of coefficients for logistic decay formulas (including the conversion from rates per month to rates per day) is in **Text S12**. The selected value for parameter $\psi$ is towards the high end of a range of estimates, as a precautionary measure given significant uncertainty [*58*].

| **Symbol** | **Definition** | **Formula/value** | **Reference** |
|---|---|---|---|
| $D_{IP,\ i}$ | Duration of Presymptomatic Infection | Gamma(1.058, 2.174) | [*51*] |
| $D_{E+IP,\ i}$ | Incubation Period (time from infection to symptoms; model structure implies that this must be >= latent period) | max(Lognormal(1.65, 0.0192), $D_{IP,\ i}$) | [*51*] |
| $D_{E,\ i}$ | Duration of Exposed stage (Time from infection to infectiousness, i.e., latent period) | $D_{E+IP,\ i}$ - $D_{IP,\ i}$ | |
| $D_{IA,\ i}$ | Duration of Asymptomatic Infection | Gamma(5, 1) | [*51*] |
| $D_{IM,\ i}$ | Duration of Mildly symptomatic Infection | Gamma(16, 0.5) | [*57*] |
| $D_{IS,\ i}$ | Duration of Severely symptomatic Infection | Gamma(34.0278, 0.4114) | [*57*] |
| $D_{IC,\ i}$ | Duration of Critically symptomatic Infection | Gamma(34.0278, 0.4114) | [*57*] |
| $\mu_{IM}$ | Mean duration (shape * scale) of mildly symptomatic infection. | 8 | [*57*] |
| $\beta_{IM}$ | Relative per-contact transmissibility during Mild infection (unitless) | 0.0253 | [*51*] |
| $\varphi$ | Parameter controlling the relative magnitude of protection from symptomatic disease given infection, and protection from severe disease given symptoms, from natural and hybrid immunity (details in **Text S3B**) | 0.5 | Assumption |
| $a_{R,\ IS}$ | Constant coefficient in logistic decay formula for overall Protection from Severe Infection granted by natural immunity | 1.70512 | [*53*] |
| $b_{R,\ IS}$ | Time-dependent coefficient (1/days) in logistic decay formula for overall Protection from Severe Infection granted by natural immunity | -0.05211/30.5 = -0.00170852459 | [*53*] |
| $a_{R,\ E}$ | Constant coefficient in logistic decay formula for Protection from Any Infection granted by natural immunity | 1.2100 | [*53*] |
| $b_{R,\ E}$ | Time-dependent coefficient (1/days) in logistic decay formula for Protection from Any Infection granted by natural immunity | -0.1937/30.5 = -0.00635081967 | [*53*] |
| $\psi$ | Relative frequency of severe infection, in the absence of any immunity, for the (original) Omicron strain relative to 2020 strains | 1.2 | [*58*] |
| $T_{ramp,\ RH}$ | Interval from beginning of ramp-up of natural or hybrid immunity (if applicable) to achieving maximum protection | 1 month (30.5 days) | [*53*] |
| $T_{total,\ R}$ | Interval of complete protection following natural recovery | 2 months (61 days) | [*53*] |
| $T_{V2 \rightarrow B}$ | Minimum interval between completion of primary series and booster dose | 5 months (152 days) | [*54*] |
| $T_{1st\ V2\ -> 0}$ | Number of days since the first individuals in the US received the secondary dose of their primary vaccine series, at the start of simulation | 1 year, 61 days (426 days) | [*54*] |
| $T_{RQ}$ | Period for which the user is asked to supply the fraction of workers who have recovered from natural infection in the past [duration] | 1 year (365 days) | |

**Table S3. Age categories and associated probabilities for stages of infection**

Notations used: IP=presymptomatic; IM=Mildly symptomatic; IA=asymptomatic; IS=Severely symptomatic infection; IC=Critically symptomatic infection; D=Death. A vertical bar indicates conditional probability, e.g., P(IP | E) should be read as "probability of IP given E," and represents the probability of a fully susceptible individual progression to the infection status IP (and subsequently, IM), given that they have reached the infection status E. Hence, all probabilities are conditional on having reached the indicated point in the symptomatic path (e.g., the probability of an 18 year old agent dying, given only that they get infected, is not 0.25, but 0.55 * 0.0036* 0.05 * 0.25 = 2.475* $10^{-5}$. Outcome probabilities are taken from [*57*] (citing [*21*, *68*, and *69*]), modified by multiplying all P(IS | IP) entries by $\psi$ = 1.2, as a precautionary measure given significant uncertainty about the severity of Omicron relative to 2020 strains [*58*].

| **Age category (years)** | **Probability of an agent being in this age category** | **P(IP \| E)** | **P(IS \| IP)** | **P(IC \| IS)** | **P(D \| IC)** |
|---|---|---|---|---|---|
| 10-19 | 0.04 | 0.55 | 0.0036 | 0.05 | 0.25 |
| 20-29 | 0.26 | 0.60 | 0.012 | 0.05 | 0.28 |
| 30-39 | 0.26 | 0.65 | 0.036 | 0.05 | 0.31 |
| 40-49 | 0.21 | 0.70 | 0.06 | 0.06 | 0.45 |
| 50-59 | 0.15 | 0.75 | 0.12 | 0.12 | 0.28 |
| 60-69 | 0.07 | 0.80 | 0.204 | 0.27 | 0.21 |
| 70-79 | 0.01 | 0.85 | 0.288 | 0.43 | 0.27 |
| 80+ | 0 | 0.90 | 0.324 | 0.71 | 0.48 |

**Table S4. Immunity events and their effects**

The definitions and effects (apart from those listed here) of the immune trajectories listed in this table are given in **Table S6-S9**. The various $P_{\text{last, X, i}}$ (where *X* is one of *E*, *IP*, or *IS*) are used to create a smooth ramp-up of immunity following a transition into V1, V2, B, HV1, HV2, or HB. These are set based on the corresponding current values (i.e., $P_{last, X, i}$,, for the same *X*), as calculated immediately prior to the updating of Immune Trajectory, $t_{last, i}$, $t_{R, i}$ and the various $P_{X, i}$ themselves. $P_{last, E, i}$ is always used (as a starting point for the ramp-up of $P_{E, i}$ (Protection from Any Infection)), but due to differences in parameterization and functional form, only one of $P_{IP, i}$ and $P_{IS, i}$ is used for a given Immunity Trajectory, as indicated in the final column of this table. As noted in **Table S16**, the default value of $T_{\text{total, R}}$ (used for all purposes other than testing sensitivity of results to $T_{\text{total, R}}$) is 61 days.

| **Event description** | **Immunity Event Time** | **Resulting Vaccination Status** | **Previous Immune Trajectory** | **Resulting Immune Trajectory** | **$t_{\text{last, i}}$ set to** | ***Which*** **$P_{\text{last, X, i}}$ are used** |
|---|---|---|---|---|---|---|
| First dose of primary vaccination | $t_{\text{V1, i}}$ | V1 | S | V1 | $t_{\text{V1, i}}$ | *E, IP* |
| | | | R | HV1 | $t_{\text{V1, i}}$ | *E, IS* |
| Second dose of primary vaccination | $t_{\text{V2, i}}$ | V2 | V1 | V2 | $t_{\text{V2, i}}$ | *E, IP* |
| | | | HV1 | HV2 | $t_{\text{V2, i}}$ | *E, IS* |
| Booster dose | $t_{\text{B, i}}$ | B | V2 | B | $t_{\text{B, i}}$ | *E, IP* |
| | | | HV2 | HB | $t_{\text{B, i}}$ | *E, IS* |
| Recovery from infection | $t_{\text{R, i}}$ | Unchanged | S, R | R | $t_{\text{R, i}}$ | NA |
| | | | V1, HV1 | HV1 | $t_{\text{R, i}}$ | NA |
| | | | V2, HV2 | HV2 | $t_{\text{R, i}}$ | NA |
| | | | B, HB | HB | $t_{\text{R, i}}$ | NA |

### Table S5. Transmissibility by infection stage

| Infection stage | Relative transmissibility per contact | Probability of transmission per contact ($p_i$) |
|---|---|---|
| IP | 1 (def.) | 0.0575 |
| IA | 0.11 | 0.006325 |
| IM | 0.44 | 0.0253 |

### Table S6. Immunity curve symbols, definitions

| Symbol | Description |
|---|---|
| S | Fully Susceptible |
| V1 | Partially Vaccinated, not recovered from natural infection within the past year |
| V2 | Fully (primarily) Vaccinated, not recovered from natural infection within the past year |
| B | Boosted, not recovered from natural infection within the past year |
| R | Recovered, unvaccinated |
| HV1 | Hybrid immunity, Partially Vaccinated |
| HV2 | Hybrid immunity, Fully (primarily) Vaccinated |
| HB | Hybrid immunity, Boosted |
| $D_{X,\,Y}(u)$ (where X is any immune trajectory and Y is one of E, IP, and IS) | A notation adopted, for the sake of convenience, for the "directly parameterized" overall protection from entering infection status Y ($P_{Y,\,i}$), at a time since employee i's last immunity event u that is sufficient for the "ramp-up" period to be over, and (if applicable) a time since employee i's last recovery that is sufficient for the period of total protection to be over. In almost all cases, this will be a decay curve.<br><br>Here "directly parameterized" means that $D_{X,\,Y}(u)$ is directly calculated from u and parameters, rather than being calculated from the value(s) of some other protection variable(s) (and possibly also additional parameters). For example, as indicated in **Tables S8** and **S9**, if employee *i* has an Immune Trajectory of V2, and $u > T_{ramp,\,V2}$, then $P_{IP,\,i}$ is equal to $\min(1, g(u, M_{V2}, r_{V2}))$, and so we call this formula $D_{V2,\,IP}(u)$. But in that same situation, $P_{E,i} = 1 - \sqrt{1 - P_{IP,i}}$, and so we do not call this $D_{V2,\,E}(u)$, but say that $D_{V2,\,E}(u)$ is "NA." |

### Table S7. Immunity curve symbols, additional notation

| Notation | Description | Equation | Used for immunity curves |
|---|---|---|---|
| ramp(u, $t_1$, $y_0$, $y_1$) | The value at time u of a linear ramp from a value of $y_0$ at time $t_0$ to a value of $y_1$ at time $t_1$ | $\frac{t_1 - u}{t_1} y_0 + \frac{u}{t_1} y_1$ | V1, V2, B, $HV1_{ramp}$, $HV2_{ramp}$, $HB_{ramp}$ |
| f(u, a, b) | The value at time u of a logistic decay (if b is negative) or growth (if b is positive) curve with a value at u = 0 of $\frac{e^a}{1+e^a}$ and a change in log odds of b per day. | $\frac{\mathrm{e}^{\mathrm{a+bu}}}{1 + \mathrm{e}^{\mathrm{a+bu}}}$ | $HV1_{ramp}$, $HV2_{ramp}$, $HB_{ramp}$ |
| g(u, M, r) | The value at time u of an exponential decay curve with initial value M and decay rate (per day) of r | $Me^{-ru}$ | V2 |
| h(u, $M_1$, $M_2$, $r_1$, $r_2$) | The value at time u of the sum of two exponentially decaying components with initial values $M_1$ and $Ma_2$ and decay rates (per day) of $r_1$ and $r_2$, respectively | $M_1 e^{-r_1 u} + M_2 e^{-r_2 u}$ | B |

**Table S8. Explanation of "directly-parameterized" immune trajectory components**

Zeros for the S immune trajectory considered indicate that all immune trajectory components are always 0. In all instances, immune trajectory components represent a waning phase, except for overall Protection from Severe Infection of individuals in HV1 and HV2 from severe infection is, which is actually growing (slowly, and from a high baseline), rather than shrinking, due to the positive sign of $b_{HV12, IS}$. NA in this table means that the value of the component in question is derived from the values of other components, as shown in **Table S9**.

| **Immune Trajectory (X)** | $D_{X, IS}(u)$ | $D_{X, IP}(u)$ | $D_{X, E}(u)$ |
|---|---|---|---|
| S | 0 | 0 | 0 |
| V1 | NA | $M_{V1}$ | NA |
| V2 | NA | $\min(1, g(u, M_{V2}, r_{V2}))$ | NA |
| B | NA | $h(u, M_{B, 1}, M_{B, 2}, r_{B, 1}, r_{B, 2})$ | NA |
| R | $f(u, a_{R, IS} + b_{R, IS})$ | NA | $f(u, a_{R, E} + b_{R, E})$ |
| HV1 | $f(u, a_{HV12, IS} + b_{HV12, IS})$ | NA | $f(u, a_{HV12, E} + b_{HV12, E})$ |
| HV2 | $f(u, a_{HV12, IS} + b_{HV12, IS})$ | NA | $f(u, a_{HV12, E} + b_{HV12, E})$ |
| HB | $f(u, a_{HB, IS} + b_{HB, IS})$ | NA | $f(u, a_{HB, E} + b_{HB, E})$ |

### Table S9. Calculation of relevant immune trajectory components

| **Immune trajectory** | **Time since last immunity event** **$u = t - t_{last, i}$** | **Time since last recovery** **$v = t - t_{R, i}$** | $P_{IS, i}$ | $P_{IP, i}$ | $P_{E, i}$ | $P_{IS \mid E, i}$ | $P_{IS \mid IP, i}$ | $P_{IP \mid E, i}$ |
|---|---|---|---|---|---|---|---|---|
| S | NA | NA | 0 | 0 | 0 | 0 | 0 | 0 |
| V1 | $u < T_{V1 \to V2}$ | NA | Unused | ramp(u, $T_{V1 \to V2}$, $P_{last, i}$, $D_{IP}(T_{V1 \to V2})$) | $1-\sqrt{1-P_{IP, i}}$ | Unused | $P_{IP \mid E, i}$ * | $1-\sqrt{1-P_{IP, i}}$ |
| V1 | $u \geq T_{V1 \to V2}$ | NA | Unused | $D_{IP}(u)$ | $1-\sqrt{1-P_{IP, i}}$ | Unused | $P_{IP \mid E, i}$ * | $1-\sqrt{1-P_{IP, i}}$ |
| V2 | $u < T_{ramp, V2}$ | NA | Unused | ramp(u, $T_{ramp, V2}$, $P_{last, i}$, $D_{IP}(T_{ramp})$) | $1-\sqrt{1-P_{IP, i}}$ | Unused | $P_{IP \mid E, i}$ * | $1-\sqrt{1-P_{IP, i}}$ |
| V2 | $u \geq T_{ramp, V2}$ | NA | Unused | $D_{IP}(u)$ | $1-\sqrt{1-P_{IP, i}}$ | Unused | $P_{IP \mid E, i}$ * | $1-\sqrt{1-P_{IP, i}}$ |
| B | $u < T_{ramp, B, 1}$ | NA | Unused | ramp(u, $T_{ramp, B, 1}$, $P_{last, i}$, $M_{mid\text{-}ramp, B}$) | $1-\sqrt{1-P_{IP, i}}$ | Unused | $P_{IP \mid E, i}$ * | $1-\sqrt{1-P_{IP, i}}$ |
| B | $T_{ramp, B, 1} \leq u < T_{ramp, B, 1} + T_{ramp, B, 2}$ | NA | Unused | ramp(u - $T_{ramp, B, 1}$, $T_{ramp, B, 2}$, $M_{mid\text{-}ramp, B}$, $D_{IP}(T_{ramp, B, 1} + T_{ramp, B, 2})$) | $1-\sqrt{1-P_{IP, i}}$ | Unused | $P_{IP \mid E, i}$ * | $1-\sqrt{1-P_{IP, i}}$ |
| B | $u \geq T_{ramp, B, 1} + T_{ramp, B, 2}$ | NA | Unused | $D_{IP}(u)$ | $1-\sqrt{1-P_{IP, i}}$ | Unused | $P_{IP \mid E, i}$ * | $1-\sqrt{1-P_{IP, i}}$ |
| R | $u < T_{total, R}$ | $v < T_{total, R}$ | 1 | 1 | 1 | 1 | 1 | 1 |
| R | $u \geq T_{total, R}$ | $v \geq T_{total, R}$ | $D_{IS}(u)$ | Unused | $D_E(u)$ | $1-\frac{1-P_{IS,i}}{1-P_{E,i}}$ | $1-(1-P_{IS\mid E,i})^{1-\phi}$ | $1-(1-P_{IS\mid E,i})^{\phi}$ |
| HV1 | $u < T_{ramp, RH}$ | $v \geq T_{total, R}$ | ramp(u, $T_{ramp, RH}$, $P_{last, i}$, $D_{IS}(T_{ramp, RH})$) | Unused | ramp(u, $T_{ramp, RH}$, $P_{last, i}$, $D_E(T_{ramp, RH})$) | $1-\frac{1-P_{IS,i}}{1-P_{E,i}}$ | $1-(1-P_{IS\mid E,i})^{1-\phi}$ | $1-(1-P_{IS\mid E,i})^{\phi}$ |
| HV1 | $u \geq T_{ramp, RH}$ | $v \geq T_{total, R}$ | $D_{IS}(u)$ | Unused | $D_E(u)$ | $1-\frac{1-P_{IS,i}}{1-P_{E,i}}$ | $1-(1-P_{IS\mid E,i})^{1-\phi}$ | $1-(1-P_{IS\mid E,i})^{\phi}$ |
| HV1 | [any] | $v < T_{total, R}$ | 1 | 1 | 1 | 1 | 1 | 1 |
| HV2 | $u < T_{ramp, RH}$ | $v \geq T_{total, R}$ | ramp(u, $T_{ramp, RH}$, $P_{last, i}$, $D_{IS}(T_{ramp, RH})$) | Unused | ramp(u, $T_{ramp, RH}$, $P_{last, i}$, $D_E(T_{ramp, RH})$) | $1-\frac{1-P_{IS,i}}{1-P_{E,i}}$ | $1-(1-P_{IS\mid E,i})^{1-\phi}$ | $1-(1-P_{IS\mid E,i})^{\phi}$ |
| HV2 | $u \geq T_{ramp, RH}$ | $v \geq T_{total, R}$ | $D_{IS}(u)$ | Unused | $D_E(u)$ | $1-\frac{1-P_{IS,i}}{1-P_{E,i}}$ | $1-(1-P_{IS\mid E,i})^{1-\phi}$ | $1-(1-P_{IS\mid E,i})^{\phi}$ |
| HV2 | [any] | $v < T_{total, R}$ | 1 | 1 | 1 | 1 | 1 | 1 |
| HB | $u < T_{ramp, RH}$ | $v \geq T_{total, R}$ | ramp(u, $T_{ramp, RH}$, $P_{last, i}$, $D_{IS}(T_{ramp, RH})$) | Unused | ramp(u, $T_{ramp, RH}$, $P_{last, i}$, $D_E(T_{ramp, RH})$) | $1-\frac{1-P_{IS,i}}{1-P_{E,i}}$ | $1-(1-P_{IS\mid E,i})^{1-\phi}$ | $1-(1-P_{IS\mid E,i})^{\phi}$ |
| HB | $u \geq T_{ramp, RH}$ | $v \geq T_{total, R}$ | $D_{IS}(u)$ | Unused | $D_E(u)$ | $1-\frac{1-P_{IS,i}}{1-P_{E,i}}$ | $1-(1-P_{IS\mid E,i})^{1-\phi}$ | $1-(1-P_{IS\mid E,i})^{\phi}$ |
| HB | [any] | $v < T_{total, R}$ | 1 | 1 | 1 | 1 | 1 | 1 |

*By assumption, in the absence of sufficient data.

**Table S10. Work and sleep schedules by day, shift, and agent "type"**

Note that fractions (e.g., "1/2 work, 1/2 awake non-work") indicate the (conceptual) probability that any given agent of that type is doing the indicated thing on a given shift.

| | **Shift 1 worker** (Farm worker, Production Shift 1 worker) | **Shift 2 worker** (Production Shift 2 worker) | **Shift 3 worker** (Cleaning Shift worker) | **Between-shift "floating" worker with 1 Production shift/day** | **Between-shift "floating" worker with 2 Production shifts/day** |
|---|---|---|---|---|---|
| Work day, shift 1 | Work | Sleep | Awake non-work | 1/2 work, 1/2 awake non-work | 1/3 work, 1/3 awake non-work, 1/3 sleep |
| Work day, shift 2 | Awake non-work | Work | Sleep | 1/2 awake non-work, 1/2 sleep | 1/3 work, 1/3 awake non-work, 1/3 sleep |
| Work day, shift 3 | Sleep | Awake non-work | Work | 1/2 work, 1/2 sleep | 1/3 work, 1/3 awake non-work, 1/3 sleep |
| Non-working day, shift 1 | Awake non-work | Sleep | Awake non-work | Awake non-work | 2/3 awake non-work, 1/3 sleep |
| Non-working day, shift 2 | Awake non-work | Awake non-work | Sleep | 1/2 awake non-work, 1/2 sleep | 2/3 awake non-work, 1/3 sleep |
| Non-working day, shift 3 | Sleep | Awake non-work | Awake non-work | 1/2 awake non-work, 1/2 sleep | 2/3 awake non-work, 1/3 sleep |

**Table S11. Relative contact rates for the farm and facility model**

Relative frequencies were assumed based on feedback from the Industry Advisory Council for the study. Note: where a pair of employees may meet on more than one shift (possible only if both are all-shifts floaters), the relative frequency listed (o_hh) is the daily total; the average per-shift frequency is $\frac{1}{n_{sh}+1}$ times this.

| Variable name | Closeness in hierarchy | Roles (lower-ranked or more localized first) | Relative frequency |
|---|---|---|---|
| **Farm model** | | | |
| c_ww | Same crew | Worker-worker | 1 (reference) |
| t_ww | Same team, different crews | Worker-worker | 0.1 |
| o_ww | Same facility, different teams | Worker-worker | 0.1 |
| t_ff | Same team, different crews (by definition) | Foreman-foreman | 0.2 |
| o_ff | Different teams | Foreman-foreman | 0.1 |
| c_wf | Same crew | Worker-foreman | 1 |
| t_wf | Same team, different crews | Worker- | 0.1 |
| o_wf | Different teams | Worker- | 0.1 |
| o_ss | Different teams (by definition) | Supervisor-supervisor | 0.2 |
| t_ws | Same team | Worker-supervisor | 0.3 |
| o_ws | Different teams | Worker-supervisor | 0.1 |
| t_fs | Same team | Foreman-supervisor | 1 |
| o_fs | Different teams | Foreman-supervisor | 0.1 |
| o_wm | Same facility | Worker-manager | 0.01 |
| o_fm | Same facility | Foreman-manager | 0.1 |
| o_sm | Same facility | Supervisor-manager | 1 |
| **Facility model** | | | |
| c_ww | Same production line | Line worker–line worker | 1 (reference) |
| t_ww | Same shift, different production line | Line worker–line worker | 0.1 |
| o_ww | Same facility, different shifts | Line worker–line worker | 0 |
| o_ss | Different shifts (by definition) | Production shift supervisor–production-shift supervisor | 0 |
| t_ws | Same shift | Line worker– production-shift supervisor | $\chi_s$ |
| o_ws | Different shifts | Line worker– production-shift supervisor | 0 |
| t_gg | Same shift | Within-shift floating worker–within-shift floating worker | $\chi_s$ |
| o_gg | Different shifts | Within-shift floating worker–within-shift floating worker | 0 |
| t_wg | Same shift | Line worker–within-shift floating worker | $\chi_s$ |
| o_wg | Different shifts | Line worker–within-shift floating worker | 0 |
| t_gs | Same shift | Within-shift floating worker–-supervisor | $\chi_s$ |
| o_gs | Different shifts | Within-shift floating worker–supervisor | 0 |
| t_cc | Same shift (by definition) | Cleaning shift worker–cleaning shift worker | $\frac{\Sigma_t}{n_{cs}-1}$ |
| o_wh | Same facility | Line worker–between-shift floating worker | $\chi_h$ |
| o_sh | Same facility | Production shift supervisor –between-shift floating worker | $\chi_h$ |
| o_gh | Same facility | Within-shift floating worker–between-shift floating worker | $\chi_h$ |
| o_hh | Same facility | Between-shift floating worker–between-shift floating worker | $(n_{sh} + 1)\chi_h$ |

**Table S12. Intermediate values used in calculating relative contact rates for the facility model in Table S11**

As in **Table S11**, all rates and sums of rates are relative to a reference value of $c_{ww} = 1$, and must be scaled as described in **Text S4D** to obtain absolute contact rates.

| Variable name | Definition | Value | Notes |
|---|---|---|---|
| $\Sigma_{ww}$ | Total contact rate between one line worker and all other line workers | $c\_ww * (n_{w,l} - 1) + t\_ww * (n_l - 1) * n_{w,l}$ | |
| $N_{w,t}$ | Number of line workers on a shift | $n_{w,l} * n_l$ | |
| $\chi_s$ | Contact rate between each supervisor or within-shit floating worker and each other individual on the same shift | $\frac{\Sigma_{ww}}{N_{w,t} - 1}$ | Results in the same total contact of contacts with other individuals on the same shift for line workers, supervisors, and within-shift floaters. The -1 accounts for the fact that, because contacts are symmetric, these contacts increase the total contact rate of line workers as well as supervisors and within-shift floating workers. |
| $\Sigma_{sh}$ | Total number of employees on a production shift | $(n_{w,l} * n_l + n_{f,sh} + 1)$ | Including supervisor |
| $\Sigma_t$ | Total contact rate between one production shift worker (i.e., line worker, within-shift floating worker, or shift supervisor) and all other production shift workers (not including all-shift floaters) | $\chi_s * \Sigma_{sh}$ | This form of the equation is most natural if thinking of the supervisor as the one production shift worker in question, but the number is equal if anyone else is used. Use in calculating c_cc and $\chi_h$ results in all workers having the total daily contact rate). |
| $\Sigma_{all}$ | For an all-shift floater, the number of combinations of another employee (including other all-shift floaters) and a shift such that it is possible for the all-shift floater to make contact with that employee on that shift | $n_{sh} * \Sigma_{sh} + n_{cs} + (n_{sh} + 1) * (n_{f,all} - 1)$ | |
| $\chi_h$ | Average per-shift contact rate between an all-shift floater and any other employee who *may* be present on the same shift | $\frac{\Sigma_t}{\Sigma_{all} - n_{all}}$ | |

### Table S13. Calculation of numbers of employees with specified characteristics, including total number of employees (N)

| Variable name | Definition | Value | Notes |
|---|---|---|---|
| **Produce Model** | | | |
| $N_{w, c}$ | Number of employees per crew, including foreman | $n_{w, c} + 1$ | +1 for the foreman |
| $N_{w, t}$ | Number of employees per supervisor, including supervisor | $N_{w, c} * n_c + 1$ | +1 for the supervisor |
| N | Total number of employees, including manager | $N_{w, t} * n_s + 1$ | +1 for the manager |
| **Facility Model** | | | |
| $N_{w,t}$ | Number of line workers on a shift | $n_{w, l} * n_l$ | No +1 because we do not have line foremen in the facility model |
| $\Sigma_{sh}$ | Total number of employees on a production shift | $(n_{w, l} * n_l + n_{f, sh} + 1)$ | +1 for the shift supervisor |
| N | Total number of employees | $n_{sh} * \Sigma_{sh} + n_{cs} + n_{f, all}$ | No +1 because we define $n_{f, all}$ as including the manager |

### Table S14. Summary of agent attributes, with additional information compared to Table 1

$P_{last, IP, i}$ and $P_{last, IS, i}$ are explained in **Table S4**. *'Ramp-up' refers to an initial increase in immunity following a vaccination event.

| Symbol | Description | Directly affect(s) | Set at simulation start | Updated during run | Reference/Notes |
|---|---|---|---|---|---|
| $A_i$ | Age | Transition probabilities | Randomly, based on age-distribution of US agricultural workforce | No | [*50*], details in **Table S3** |
| $p_{stage, i}$ | Transition probabilities at full susceptibility | Asymptomatic vs. Symptomatic infection; recovery from each stage on the symptomatic path vs. further progression/ death | Based on age | No | [*57*], *citing* [*21, 68, and 69*] Partial immunity is also incorporated into the "decision" between transitions, but is recorded as a separate variable |
| $D_{stage, i}$ | Infection stage durations | Timing of recovery/ progression/ death | Randomly | No | [*51, 57*]; distributions in **Table S2.** Precalculated for each run for computational convenience |
| $t_{event\_type, i}$ ($t_{V1, i}$, $t_{V2, i}$, $t_{B, i}$, and $t_{R, i}$) | Immunity event times | Protection against Any Infection ($P_{E, i}$) and either overall Protection against Symptomatic Infection ($P_{IP, i}$) or overall Protection against Severe Infection ($P_{IS, i}$) | Randomly, based in part on user-supplied fraction of employees with the relevant event within the applicable past interval | Updated when immunity events occur | Only records the most recent time for a given class of immunity event. Undefined for events that have never happened yet (set to + infinity, as a computational convenience). Note that while these events types share names (V1, V2, B, and R) with immune trajectories, not everyone with a finite value for one of these events entered that immune trajectory at the time, because they may have entered a hybrid trajectory instead. |
| $t_{infection\ status, i}$ ($t_{E, i}$, $t_{IA, i}$, $t_{IP, i}$, $t_{IM, i}$, $t_{IS, i}$, and $t_{IC, i}$) | Infection and infection progression times | Timing of recovery/ progression/ death | Randomly, for individuals who start the simulation infected | Updated upon infection and progression from one stage of infection to another | Only records the most recent time of entry into a given infection status. Undefined for events that have never happened yet (set to + infinity, as a computational convenience). There is no $t_{NI, i}$ as such, but if there were, it would be equal to $t_{R, i}$. |
| $C_i$ | Immunity trajectories | Protection against Any Infection ($P_{E, i}$) and either overall Protection against Symptomatic Infection ($P_{IP, i}$) or overall Protection against Severe Infection ($P_{IS, i}$) | Calculated from nature and order of (finite) event times | Recalculated when immunity events occur | |
| $t_{last, i}$ | Time of last immunity event | Immunity components (for certain immunity trajectories) | Maximum (finite) immunity event time | Updated whenever an immunity event occurs | Relevant because of immunity ramp-up and waning. Considered to be undefined if immunity event times are infinite (i.e., the individual is fully Susceptible (S)). |
| $P_{last, E, i}$ | "Previous" (at the time of the last immunity event) level of Protection against Any Infection ($P_{E, i}$) | Current level of Protection against Any Infection, during ramp-up phases only | 0 (or undefined) for fully susceptible agents and agents in immunity trajectories without ramp-up. Otherwise calculated in the same fashion as $P_{E, i}$, but at the time of the last immunity event | Updated (to current $P_{E, i}$ infinitesimally prior to the event) whenever an immunity event occurs | |

| | | | | | |
|---|---|---|---|---|---|
| $P_{\text{last, IP, i}}$ | "Previous" (at the time of the last immunity event) level of overall Protection against Symptomatic Infection ($P_{\text{IP, i}}$;) | Current overall Protection against Symptomatic Infection, during ramp-up phases only | 0 (or undefined) for fully susceptible agents. Otherwise calculated in the same fashion as $P_{\text{IP, i}}$, but at the time of the last immunity event | Updated (to current $P_{\text{IP, i}}$ infinitesimally prior to the event) whenever an immunity event occurs | May be left undefined when entering certain immunity trajectories for which components are calculated in a different fashion |
| $P_{\text{last, IS, i}}$ | "Previous" (at the time of the last immunity event) level of overall Protection against Severe Infection ($P_{\text{IS, i}}$) | Current overall Protection against Severe Infection, during ramp-up phases only | 0 (or undefined) for fully susceptible agents. Otherwise calculated in the same fashion as $P_{\text{IS, i}}$, but at the time of the last immunity event | Updated (to current $P_{\text{IS, i}}$ infinitesimally prior to the event) whenever an immunity event occurs | May be left undefined when entering certain immunity trajectories for which components are calculated in a different fashion |
| $P_{\text{E, i}}$ | Protection against Any Infection | Relative susceptibility to infection (i.e., to transitioning from Not Infected to Exposed) | Calculated at t = 0 from immunity trajectories, time of last immunity event, and (only during ramp-up) $P_{\text{last, E, i}}$ | Recalculated at the start of each shift from immunity trajectories, time of last immunity event, and (only during ramp-up) $P_{\text{last, E, i}}$ | |
| $P_{\text{IP \| E, i}}$ | Protection against Symptomatic Infection given Any Infection | Relative probability of transitioning from Exposed to Presymptomatically Infected (rather than to Asymptomatically Infected) | Calculated at t = 0 from immunity trajectories, time of last immunity event, and (only during ramp-up) either $P_{\text{last, IP, I}}$ or $P_{\text{last, IS, i}}$ | Recalculated at the start of each shift from immunity trajectories, time of last immunity event, and (only during ramp-up) either $P_{\text{last, IP, I}}$ or $P_{\text{last, IS, i}}$ | |
| $P_{\text{IS \| IP, i}}$ | Protection against Severe Infection given Symptomatic Infection | Relative probability of transitioning from Mildly Infected to Severely Infected (rather than recovering) | Calculated at t = 0 from immunity trajectories, time of last immunity event, and (only during ramp-up) either $P_{\text{last, IP, I}}$ or $P_{\text{last, IS, i}}$ | Recalculated at the start of each shift from immunity trajectories, time of last immunity event, and (only during ramp-up) either $P_{\text{last, IP, I}}$ or $P_{\text{last, IS, i}}$ | |
| $B_{\text{on time, i}}$ | Boosting on time | Whether the agent has received/will receive a booster shot $T_{V2 \rightarrow B}$ = 5 months after the second shot of their primary series (if any) | Randomly set at simulation start based on user-supplied information about the percent of their employees eligible for a booster who have actually received one | No | The fraction (unobserved) of currently ineligible agents for whom this is True is set equal to the corresponding fraction (observed) of eligible agents |
| $V_{\text{i}}$ | Vaccination status | Eligibility for future shots | Randomly set at simulation start based on user-supplied information | Updated upon receiving an additional shot (whether primary series or boosting) | Does not affect immunity directly (although vaccination updates both vaccination status and immunity trajectories) |
| $I_{\text{i}}$ | Infection status | Transmissibility; hospitalization; progression, death, and recovery | User-set number of randomly-selected agents infected at baseline; all others Not Infected | Updated upon infection, progression, recovery, and death | |
| $t_{\text{tested, i}}$ | Most recent time tested, if any | Priority for future testing | Uniformly negative infinity, representing no employees having been tested recently enough to matter | Updated upon testing | |
| $t_{\text{Q, i}}$ | Time isolated | Eligibility for deisolation | Uniformly negative infinity, representing no employees having been isolated recently enough to matter | Updated upon isolation, and upon development of symptoms while already isolated due to a (presymptomatic) positive test result | |
| $Q_i$ | Isolation status | Eligibility for deisolation, presence or absence at work and, if applicable, shared housing (and hence, potential to transmit) | Uniformly false | Updated upon isolation and deisolation | |

### Table S15. Factors tested in scenario analysis

| Factor/intervention | Setting or values | Note |
|---|---|---|
| **Scenario parameters** | | |
| Setting | Farm | |
| | Facility | |
| Housing | Individual | Non-zero force of infection due to community transmission during shifts that are neither working nor sleeping |
| | Shared | Non-zero contact rate between employees during shifts that are neither working nor sleeping (leading to dormitory transmission) |
| Vaccinated | High (default) | Based on US national levels in early 2022 [*32*,*33*) (Derivation in **Text S11**) |
| | None | |
| Recovered | High (default) | Based on US national levels in early 2022 [*32, 34, 35*) (Derivation in **Text S11**) |
| | Nothing | |
| **Intervention parameters** | | Only one set to an intervening value at a time in the analyses presented in this paper, except that vaccination and boosting are allowed to both be 0.02. |
| Temperature screening | True | Everyone is assumed to be screened every day that are scheduled to work and are not already unavailable. |
| | False | |
| Viral testing (probability per work day) | 0 | No testing |
| | 0.05 | 5% of scheduled workers who are not already unavailable tested each shift, amounting to testing every worker approximately once every 4 weeks |
| | 0.3 | 30% of scheduled workers who are not already unavailable tested each shift, amounting to testing every worker about 1.5 times per week |
| | 1 | Every scheduled worker who is not already unavailable tested each shift |
| $R_0$ reduction | -20% | Represented by the use of KN95 masks, one per employee per shift |
| | -40% | Represented by masking and face shield use (one/employee/30 days), without ventilation improvements |
| | -80% | Represented by a combination of masking, face shield use, and ventilation improvements (e.g., portable air cleaner) |
| Vaccination (probability per day) | 0.02 | 2% probability of each currently unvaccinated worker receiving their first dose each day that they are not currently infected or isolated (and their second, $T_{V1\rightarrow V2}$ = 21 days later) |
| | 0.04 | 4% probability of each currently unvaccinated worker receiving their first dose each day that they are not currently infected or isolated (and their second, $T_{V1\rightarrow V2}$ = 21 days later) |
| Boosting (probability per day) | 0.02 | 2% probability of each worker who has received their dose at least $T_{V2\rightarrow B}$ = 5 months ago but is currently unboosted receiving their booster dose each day that they are not currently infected or isolated |
| | 0.04 | 4% probability of each worker who has received their dose at least $T_{V2\rightarrow B}$ = 5 months ago but is currently unboosted receiving their booster dose each day that they are not currently infected or isolated |
| Vaccination + boosting (probability per day) | 0.02 | A combination of the vaccination at 2% probability per day and boosting at 2% probability per day interventions |

### Table S16. Sensitivity Parameters

| Parameter (variable) name | Symbol | Definition | Unit | Value | Notes |
|---|---|---|---|---|---|
| isolation_duration | | Length of isolation following a positive viral test or temperature screening; reset following a subsequent onset of symptoms | days | 5 | |
| mu | $\mu_{E+IP}$ | Mean duration (with exceptions, see: D_{E+IP, i}) of incubation period | days | 5.2 | |
| sd | $\sigma_{E+IP}$ | Standard deviation of duration (with exceptions, see: D_{E+IP, i}) of incubation period | days | 0.1 | |
| duration_IP_mean | $\mu_{IP}$ | Mean duration (shape * scale) of presymptomatic infection | days | 1.058*2.174 | This and the other mean parameters are presented as products, to clarify derivation from [*51*] |
| duration_IP_shape | $k_{IP}$ | Shape parameter for duration of presymptomatic infection | Unitless | 1.058 | |
| duration_IA_mean | $\mu_{IA}$ | Mean duration (shape * scale) of asymptomatic infection | days | 1*5 | |
| duration_IA_shape | $k_{IA}$ | Shape parameter for duration of asymptomatic infection | Unitless | 5 | |
| duration_IM_mean | $\mu_{IM}$ | Mean duration (shape * scale) of mildly symptomatic infection | days | 0.5*16 | |
| duration_IM_shape | $k_{IM}$ | Shape parameter for duration of mildly symptomatic infection | Unitless | 16 | |
| duration_IS_mean | $\mu_{IS}$ | Mean duration (shape * scale) of severe infection | days | 0.4114*34.0278 | |
| duration_IS_shape | $k_{IS}$ | Shape parameter for duration of severe infection | Unitless | 34.0278 | |
| duration_IC_mean | $\mu_{IC}$ | Mean duration (shape * scale) of critical infection | days | 0.4114*34.0278 | |
| duration_IC_shape | $k_{IC}$ | Shape parameter for duration of critical infection | Unitless | 34.0278 | |
| p_trans_IP | $\beta_{IP}$ | Relative per-contact transmissibility during presymptomatic infection | Unitless | 0.0575 | |
| p_trans_IA | $\beta_{IA}$ | Relative per-contact transmissibility during asymptomatic infection | Unitless | 0.0575*0.11 | |
| p_trans_IM | $\beta_{IM}$ | Relative per-contact transmissibility during mildly symptomatic infection | Unitless | 0.0575*0.44 | |
| boosting_interval | $T_{V2 \rightarrow B}$ | Minimum interval between completion of primary series and booster dose | days | 152 | |
| second_shot_interval | $T_{V1 \rightarrow V2}$ | Interval between first and second shots of two-dose primary series | days | 21 | |
| max_V1_protection | $M_{V1}$ | Maximum net protection from symptomatic infection from a single dose of a two-dose primary series, without hybrid immunity | Unitless | 0.36 | |
| V2_ramp_time | $T_{ramp, V2}$ | Interval from receiving second dose of two-dose primary series to achieving maximum protection, for an individual without hybrid immunity | days | 14 | |
| V2_magnitude | $M_{V2}$ | Magnitude of exponentially decaying overall protection from symptomatic infection from complete two-dose primary series, for an individual without hybrid immunity | | 0.9115739 | I.e., this is what protection would be immediately upon receiving the shot if $T_{ramp, V2}$ were 0. The actual maximum protection achieved is $M_{V2}$ * exp(-$r_{V2}$ * $T_{ramp, V2}$), which is somewhat lower. |
| V2_decay_rate | $r_{V2}$ | Rate of exponential decay of net protection from symptomatic infection obtained from a complete two-dose primary series, for an individual without hybrid immunity | 1/days | 0.08904459/7 | The retention of "/7" reflects the derivation from week-granularity data |
| B_ramp_time_1 | $t_{ramp, B, 1}$ | Interval from receiving booster dose to achieving net protection from symptomatic infection given by B_mid_ramp_protection, for an individual without hybrid immunity | days | 7 | |

| | | | | | |
|---|---|---|---|---|---|
| B_ramp_time_2 | $t_{ramp, B, 2}$ | Interval from achieving net protection from symptomatic infection given by B_mid_ramp_protection, following a booster dose, to achieving maximum protection, for an individual without hybrid immunity | days | 7 | |
| B_mid_ramp_protection | $M_{mid-ramp,B}$ | Net protection from symptomatic infection achieved B_ramp_time_1 days after receiving a booster dose, for an individual without hybrid immunity | Unitless | 0.62 | |
| B_magnitude_1 | $M_{B, 1}$ | Magnitude of ***rapidly*** (exponentially-)decaying component of net protection from symptomatic infection given by receiving a booster dose, for an individual without hybrid immunity | Unitless | 0.471669758 | I.e., this is what the rapidly-decaying component of net protection would be immediately upon receiving the shot if $T_{ramp, B, 1} = T_{ramp, B, 2} = 0$. Since the actual maximum value achieved of the protection from this component is $M_{B, 1} * \exp(-r_{B, 1} * (T_{ramp, B, 1} + T_{ramp, B, 2}))$, it's lower. |
| B_magnitude_2 | $M_{B, 2}$ | Magnitude of ***slowly*** (exponentially-)decaying component of net protection from symptomatic infection given by receiving a booster dose, for an individual without hybrid immunity | Unitless | 0.32660087 | I.e., this is what the slowly-decaying component of net protection would be immediately upon receiving the shot if $T_{ramp, B, 1} = T_{ramp, B, 2} = 0$. Since the actual maximum value achieved of the protection from this component is $M_{B, 2} * \exp(-r_{B, 2} * (T_{ramp, B, 1} + T_{ramp, B, 2}))$, it's lower. |
| B_decay_rate_1 | $r_{B, 1}$ | Rate of exponential decay of ***rapidly***-decaying component of protection from symptomatic infection given by receiving a booster dose, for an individual without hybrid immunity | 1/days | 0.083161719/7 | |
| B_decay_rate_2 | $r_{B, 2}$ | Rate of exponential decay of ***slowly***-decaying component of protection from symptomatic infection given by receiving a booster dose, for an individual without hybrid immunity | 1/days | 0.008970573/7 | |
| SEVERE_MULTIPLIER | $\psi$ | Multiplier on age-specific probabilities (originally inferred for 2020 strains) of severe disease (requiring hospitalization), given symptomatic disease and 0 immunity | Unitless | 1.2 | |
| R_question_period | $T_{RQ}$ | Period for which the user is asked to supply the fraction of workers who have recovered from natural infection in the past [duration] | days | 365 | Or, looked at another way, for the purpose of sensitivity testing, the period of time over which the given number of recovered individuals (with or without hybrid immunity) at simulation start have made their recoveries. |
| time_since_first_V2 | $T_{1st\ V2 \to 0}$ | Number of days since the first individuals in the US received the secondary dose of their primary vaccine series, at the start of simulation | days | 365+61 | |
| R_nsp_a | $a_{R, IS}$ | Constant coefficient in logistic decay formula for net protection from severe infection granted by natural immunity, if not also vaccinated | Unitless | 1.70512 | Hence, log odds of the protection that would be granted immediately upon recovery, if not for the fact that protection is complete for two months following recovery |
| R_nsp_b | $b_{R, IS}$ | Time-dependent coefficient in logistic decay formula for net protection from severe infection granted by natural immunity, if not also vaccinated | 1/days | -0.001708525 | Note that here and for hybrid immunity, we give the coefficient itself (which is negative), unlike for V2 and B, where we give the decay rate (which is positive) |
| H_RV12_nsp_a | $a_{HV12, IS}$ | Constant coefficient in logistic "decay" formula for net protection from severe infection granted by hybrid immunity with either 1 or 2 doses of primary vaccination | Unitless | 3.0473642 | This is calculated from [*53*] for 2 doses, but is assumed to apply equally to 1 and 2 doses in the absence of better data. |

| | | | | | |
|---|---|---|---|---|---|
| H_RV12_nsp_b | $b_{HV12, IS}$ | Time-dependent coefficient in logistic “decay” formula for net protection from severe infection granted by hybrid immunity with either 1 or 2 doses of primary vaccination | 1/days | 0.04724741/30.5 | This is calculated from [*53*] for 2 doses, but is assumed to apply equally to 1 and 2 doses in the absence of better data.<br>Note that this coefficient is positive, meaning that it's actually a slow logistic _growth_ of immunity over time, reflecting data from [*53*]. The “30.5” reflects data originally given in months. |
| H_RB_nsp_a | $a_{HB, IS}$ | Constant coefficient in logistic decay formula for net protection from severe infection granted by hybrid immunity with a booster dose | Unitless | 4.0685452 | |
| H_RB_nsp_b | $b_{HB, IS}$ | Time-dependent coefficient in logistic decay formula for net protection from severe infection granted by hybrid immunity with a booster dose | 1/days | -0.005758993 | |
| R_ip_a | $a_{R,E}$ | Constant coefficient in logistic decay formula for net protection from any infection granted by natural immunity, if not also vaccinated | Unitless | 1.21 | |
| R_ip_b | $b_{R,E}$ | Time-dependent coefficient in logistic decay formula for net protection from any infection granted by natural immunity, if not also vaccinated | 1/days | -0.00635082 | |
| H_RV12_ip_a | $a_{HV12,E}$ | Constant coefficient in logistic decay formula for net protection from any infection granted by hybrid immunity with either 1 or 2 doses of primary vaccination | Unitless | 1.176188 | This is calculated from [*53*] for 2 doses, but is assumed to apply equally to 1 and 2 doses in the absence of better data |
| H_RV12_ip_b | $b_{HV12,E}$ | Time-dependent coefficient in logistic decay formula for net protection from any infection granted by hybrid immunity with either 1 or 2 doses of primary vaccination | 1/days | -0.00412059 | This is calculated from [*53*] for 2 doses, but is assumed to apply equally to 1 and 2 doses in the absence of better data. |
| H_RB_ip_a | $a_{HB,E}$ | Constant coefficient in logistic decay formula for net protection from any infection granted by hybrid immunity with a booster dose | Unitless | 1.7006945 | |
| H_RB_ip_b | $b_{HB,E}$ | Time-dependent coefficient in logistic decay formula for net protection from any infection granted by hybrid immunity with a booster dose | 1/days | -0.010059308 | |
| hybrid_ramp_time | $T_{ramp, RH}$ | Interval from beginning ramp-up of natural or hybrid immunity (if applicable) to achieving maximum protection | days | 30.5 | 1 month |
| recovered_complete_protection_time | $T_{total, R}$ | Interval of complete protection following natural recovery | days | 61 | 2 months |
| fraction_ssp_symptomatic | $\phi$ | Measure of how much of the protection from severe disease, conditional on any infection, that is attributable to natural or hybrid immunity is due to protection from symptomatic disease, conditional on any infection | | 0.5 | |

**References**

(numbering reflects numbers in the main text)

21. Ferguson, N. M. et al. Report 9 - Impact of non-pharmaceutical interventions (NPIs) to reduce COVID-19 mortality and healthcare demand. https://www.imperial.ac.uk/mrc-global-infectious-disease-analysis/disease-areas/covid-19/report-9-impact-of-npis-on-covid-19/ (2020).
32. The U. S. Census Bureau. United States. https://data.census.gov/cedsci/profile?q=United%20States&g=0100000US (2022).
33. Centers for Disease Control and Prevention, (CDC). COVID-19 vaccination demographics in the United States, National. https://data.cdc.gov/Vaccinations/COVID-19-Vaccination-Demographics-in-the-United-St/km4m-vcsb/about_data (2022).
34. Centers for Disease Control and Prevention, (CDC). COVID data tracker. https://covid.cdc.gov/covid-data-tracker (2022).
35. Centers for Disease Control and Prevention, (CDC). Estimated COVID-19 burden. https://www.cdc.gov/coronavirus/2019-ncov/cases-updates/burden.html (2022).
51. Moghadas, S. M. et al. The implications of silent transmission for the control of COVID-19 outbreaks. *Proc. Natl. Acad. Sci.* **117**, 17513–17515 (2020).
52. U.K. Health Security Agency. COVID-19 vaccine surveillance report week 4. https://assets.publishing.service.gov.uk/government/uploads/system/uploads/attachment_data/file/1050721/Vaccine-surveillance-report-week-4.pdf (2022).
53. Bobrovitz, N. et al. Protective effectiveness of previous SARS-CoV-2 infection and hybrid immunity against the omicron variant and severe disease: A systematic review and meta-regression. *Lancet Infect. Dis.* **23**, 556–567 (2023).
54. Pfizer. Vaccine information fact sheet for recipients and caregivers about Comirnaty (COVID-19 vaccine, mRNA), the Pfizer-Biontech COVID-19 vaccine, and the Pfizer-Biontech COVID-19 vaccine, bivalent (original and omicron BA.4/BA.5) to prevent coronavirus disease 2019 (COVID-19) for individuals 12 years of age and older. https://labeling.pfizer.com/ShowLabeling.aspx?id=14472&format=pdf (2022).
55. R Core Team. R: A language and environment for statistical computing. R Foundation for Statistical Computing, Vienna, Austria. https://www.R-project.org/ (2020).
56. Ahmad, S. Economic theory, applications and issues - estimating input-mix efficiency in a parametric framework: application to state-level agricultural data for the United States. ISSN: 1444-8890 (2017).
57. Kerr, C. C. et al. Covasim: An agent-based model of COVID-19 dynamics and interventions. *PLoS Comput. Biol.* **17**, e1009149 (2021).
58. Arabi, M. et al. Severity of the Omicron SARS-CoV-2 variant compared with the previous lineages: A systematic review. *J. Cell Mol. Med.* **27**, 1443–1464 (2023).
59. Bielecki, M., Crameri, G. A. G., Schlagenhauf, P., Buehrer, T. W. & Deuel, J. W. Body temperature screening to identify SARS-CoV-2 infected young adult travelers is ineffective. *Travel Medicine and Infectious Disease* **37**, 101832 (2020).
60. Okoye, G. A. et al. Diagnostic accuracy of a rapid diagnostic test for the early detection of COVID-19. *J. Clin. Virol.* **147**, 105023 (2022).
61. Center for Disease Prevention, (CDC). Ending isolation and precautions for people with COVID-19: interim guidance. www.cdc.gov/coronavirus/2019-ncov/hcp/duration-isolation.html (2022).
